# Within-country age-based prioritisation, global allocation, and public health impact of a vaccine against SARS-CoV-2: a mathematical modelling analysis

**DOI:** 10.1101/2021.03.19.21253960

**Authors:** Alexandra B Hogan, Peter Winskill, Oliver J Watson, Patrick G T Walker, Charles Whittaker, Marc Baguelin, Nicholas F Brazeau, Giovanni D Charles, Katy A M Gaythorpe, Arran Hamlet, Edward Knock, Daniel J Laydon, John A Lees, Alessandra Løchen, Robert Verity, Lilith K Whittles, Farzana Muhib, Katharina Hauck, Neil M Ferguson, Azra C Ghani

## Abstract

The worldwide endeavour to develop safe and effective COVID-19 vaccines has been extraordinary, and vaccination is now underway in many countries. However, the doses available in 2021 are likely to be limited. We extended a mathematical model of SARS-CoV-2 transmission across different country settings to evaluate the public health impact of potential vaccines using WHO-developed target product profiles. We identified optimal vaccine allocation strategies within- and between-countries to maximise averted deaths under constraints on dose supply. We found that the health impact of SARS-CoV-2 vaccination depends on the cumulative population-level infection incidence when vaccination begins, the duration of natural immunity, the trajectory of the epidemic prior to vaccination, and the level of healthcare available to effectively treat those with disease. Within a country we find that for a limited supply (doses for <20% of the population) the optimal strategy is to target the elderly. However, with a larger supply, if vaccination can occur while other interventions are maintained, the optimal strategy switches to targeting key transmitters to indirectly protect the vulnerable. As supply increases, vaccines that reduce or block infection have a greater impact than those that prevent disease alone due to the indirect protection provided to high-risk groups. Given a 2 billion global dose supply in 2021, we find that a strategy in which doses are allocated to countries proportional to population size is close to optimal in averting deaths and aligns with the ethical principles agreed in pandemic preparedness planning.

**Highlights:**

- The global dose supply of COVID-19 vaccines will be constrained in 2021
- Within a country, prioritising doses to protect those at highest mortality risk is efficient
- For a 2 billion dose supply in 2021, allocating to countries according to population size is efficient and equitable

## Introduction

COVID-19 has caused an unprecedented global public health and economic challenge which will continue to disrupt lives and livelihoods until a preventative intervention, such as a vaccine, becomes widely available. Demand for a vaccine is unparalleled and extraordinary public and private sector efforts have been undertaken to identify vaccine candidates, to conduct clinical trials on compressed timelines and to scale up manufacturing ahead of regulatory approval. To date, 13 vaccines have been approved or authorised for use by individual or groups of countries, and many more vaccine candidates are in development^1–6^.

Even as some countries begin to introduce approved vaccines, the demand for doses is likely to exceed supply through 2021 due to constraints in manufacturing. Despite the development of international initiatives such as COVAX to share the risks of the research and development process, and to ensure equitable access^7^, political and economic incentives for countries to prioritise national interest remain high – as has already been demonstrated by countries stockpiling treatment supplies and other pharmaceuticals^8,9^ as well as countries signing advanced purchase agreements for individual vaccines. Four allocation principles, reflected in the current World Health Organization (WHO) global allocation framework^10^, have been identified by bioethicists to guide allocation of scarce resources: (A) that the health benefits of the resource are maximised; (B) that priority is given to those who would be worst-off in the absence of the resource; (C) that individuals in equal circumstances should be treated equally; and (D) that societal benefit is maximised^11,12^. A key component in achieving principles A and B will be targeting the vaccine to those at highest risk of death. Given the strong age-gradient of risk associated with COVID-19 infection^13^, allocation is likely to be age-targeted. However, to meet principle B, vaccine dose allocation will need to be balanced against both life-expectancy (in order to minimise life-years lost) and the additional variation in the risk of death resulting from, for example, inequitable access to healthcare across the globe. In addition, aligned with principle D, it is likely that any vaccine allocation would also prioritise essential workers such as those providing the frontline health response^14^. Adhering to these principles to derive a fair and optimal allocation strategy, given limited vaccine stocks is far from straightforward.

We extend a model of SARS-CoV-2 transmission to explore the public health impact of different vaccine characteristics, epidemic stages, and population-targeting strategies. We apply the model to countries with different levels of income to understand the impact of demographics, societal mixing and health system constraints in overall vaccine benefit. We explore the implications of these characteristics for within-country allocation and global allocation and quantify the maximum public health benefit of different allocation strategies under a range of likely supply constraints, over the time periods 2021 only and 2021–2022.

## Methods

### Mathematical Model

We extended a previously developed age-structured deterministic SEIR-type compartmental model of SARS-CoV-2 transmission^15^ to include vaccination. The model explicitly incorporates the clinical pathway for those requiring hospitalisation, allowing estimates of the need for oxygen support and/or intensive care unit (ICU) support. Transmission depends on age-based contact matrices and a constant transmission rate per contact (in sensitivity analysis we explore reduced transmission from children). Other risk groups or settings (such as healthcare workers or care homes) are not included. The model was extended to capture loss of naturally acquired immunity by including an additional flow from the recovered state to the susceptible state.

Once vaccination is introduced into a population, we assume that all eligible individuals (depending on the targeting or prioritisation that is applied) are vaccinated at a constant rate, up to a maximum level of coverage. Vaccination prioritisation strategies are modelled by specifying an age disaggregated matrix of coverage targets where rows are ordered prioritisation steps (see Supplementary Information, Section 1.2). Individuals who are susceptible, in the latent period, or recovered (immune) can be vaccinated. The model structure assumes that people who are currently infected do not receive the vaccine, and while this simplification may miss asymptomatic individuals, such people represent a small fraction of the total population at any given time. Vaccinated individuals initially enter a temporary state to capture the delay between receiving the vaccine and being protected before moving into a vaccine-protected state. We assume that protection is partial, with efficacy parameters detailed in Table 1. Given that the duration of protection for the approved and candidate vaccines is not yet known, we assume that vaccine-induced immunity can either be long-term, one year, or six months, and we model decay in vaccine-induced immunity by moving individuals from the vaccinated and protected state to a further state in which they are again susceptible to infection (unless they acquire immunity through natural infection). Protection against infection only is modelled by reducing the risk of acquiring infection in those who are vaccinated by a constant factor, capturing the fact that vaccines will not give complete protection against infection. Protection against severe disease only is captured by reducing the proportion of cases that are hospitalised. Combined protection (the default setting) assumes protection against infection, with an additional reduction in the risk of severe disease in vaccinated individuals who experience a breakthrough infection. Additional details of the model are given in the Supplementary Information (Figure S1; Tables S1 and S2).

**Table 1:**
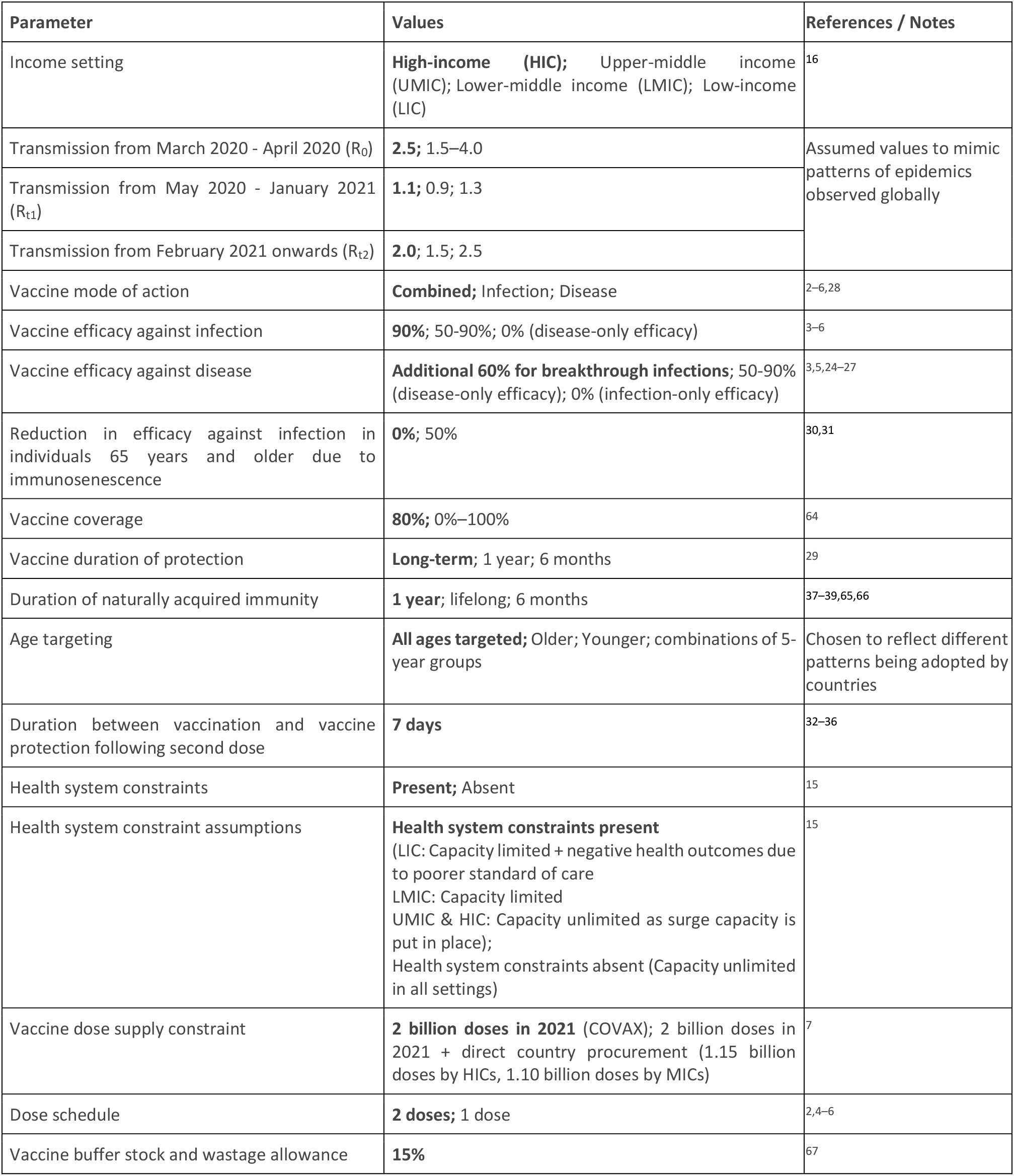
Summary of Scenarios Explored. The values in bold represent the default parameters, unless otherwise stated.

| Parameter | Values | References / Notes |
| --- | --- | --- |
| Income setting | **High-income (HIC)**; Upper-middle income (UMIC); Lower-middle income (LMIC); Low-income (LIC) | <sup>16</sup> |
| Transmission from March 2020 - April 2020 ($R_0$) | **2.5**; 1.5–4.0 | Assumed values to mimic patterns of epidemics observed globally |
| Transmission from May 2020 - January 2021 ($R_{t1}$) | **1.1**; 0.9; 1.3 | |
| Transmission from February 2021 onwards ($R_{t2}$) | **2.0**; 1.5; 2.5 | |
| Vaccine mode of action | **Combined**; Infection; Disease | <sup>2–6,28</sup> |
| Vaccine efficacy against infection | **90%**; 50–90%; 0% (disease-only efficacy) | <sup>3–6</sup> |
| Vaccine efficacy against disease | **Additional 60% for breakthrough infections**; 50–90% (disease-only efficacy); 0% (infection-only efficacy) | <sup>3,5,24–27</sup> |
| Reduction in efficacy against infection in individuals 65 years and older due to immunosenescence | **0%**; 50% | <sup>30,31</sup> |
| Vaccine coverage | **80%**; 0%–100% | <sup>64</sup> |
| Vaccine duration of protection | **Long-term**; 1 year; 6 months | <sup>29</sup> |
| Duration of naturally acquired immunity | **1 year**; lifelong; 6 months | <sup>37–39,65,66</sup> |
| Age targeting | **All ages targeted**; Older; Younger; combinations of 5-year groups | Chosen to reflect different patterns being adopted by countries |
| Duration between vaccination and vaccine protection following second dose | **7 days** | <sup>32–36</sup> |
| Health system constraints | **Present**; Absent | <sup>15</sup> |
| Health system constraint assumptions | **Health system constraints present**<br>(LIC: Capacity limited + negative health outcomes due to poorer standard of care<br>LMIC: Capacity limited<br>UMIC & HIC: Capacity unlimited as surge capacity is put in place);<br>Health system constraints absent (Capacity unlimited in all settings) | <sup>15</sup> |
| Vaccine dose supply constraint | **2 billion doses in 2021** (COVAX); 2 billion doses in 2021 + direct country procurement (1.15 billion doses by HICs, 1.10 billion doses by MICs) | <sup>7</sup> |
| Dose schedule | **2 doses**; 1 dose | <sup>2,4–6</sup> |
| Vaccine buffer stock and wastage allowance | **15%** | <sup>67</sup> |

The open-source model code is available as an R package at https://github.com/mrc-ide/nimue. To aid utility of the modelling framework by policymakers, government agencies and other stakeholders, we additionally developed a user-friendly interface (Box 1). In the interface the model is fitted to country epidemics (see Section 1.4 in Supplementary Information) with these fits updated weekly to reflect rapidly changing epidemic situations. Updates to this interface will incorporate new information that emerges as the pandemic unfolds.

### Parameterisation

We stratify the global population into four groups, based on the current World Bank^16^ classification of high-income countries (HIC), upper-middle-income countries (UMIC), lower-middle-income countries (LMIC) and low-income countries (LIC). Countries are assumed to remain in their current group over the projection horizon. For each income setting, the age distribution of the population and age-based contact patterns are modified to capture epidemiological characteristics of countries in the different income strata. We use the current age distribution for the country with the median GDP in each of the four groups (HIC: Malta; UMIC: Grenada; LMIC: Nicaragua; and LIC: Madagascar)^17^, and age-based contact patterns representative of these settings based on the availability of contact data studies^15^. Natural history parameters for SARS-CoV-2 infection are based on a review of published estimates and large clinical studies^18,19^ with age-stratified probabilities of requiring hospitalisation and intensive care obtained from a national study of French hospital admissions^20^ and the age-stratified infection fatality ratio (IFR) obtained from a recent meta-analysis^21^ (Table S2). The model has been previously validated by using Bayesian methods to fit to country-specific data on COVID-19 deaths (Figure S2)^22,23^. Modelled levels of transmission are determined by the time-varying reproduction number, R_t_. Transmission scenarios are detailed below.

Phase III clinical trials have generally reported endpoints of symptomatic or severe COVID-19 illness. Interim efficacy estimates against symptomatic COVID-19 have been published for several candidate vaccines, including Pfizer/BioNTech (95% efficacy), Moderna (94.1%), Oxford/AstraZeneca (63.1% for two standard doses), and the Gamaleya Center (Sputnik V, 91.6%)^3–6^. Trial results to date indicate higher protection against severe COVID-19 disease, and this is supported by evidence being generated by vaccine implementation in the UK and Israel^3,5,24–27^. The extent to which these candidate vaccines will prevent infection is not fully known, however the phase III Oxford/AstraZeneca study reported efficacy against asymptomatic COVID-19 of 27.3% (95% CI −17.2–54.9) in a preliminary analysis^2^, and evidence from the UK indicates a four-fold reduction in asymptomatic COVID-19 in vaccinated versus unvaccinated healthcare workers, following a single dose of the Pfizer/BioNTech vaccine^28^. Given these results, for our baseline simulations we assume vaccine efficacy against infection of 90%, with an additional 60% efficacy against severe disease for vaccinated individuals who experience breakthrough infection^24^. We further consider 70% efficacy against infection as a sensitivity analysis, which also aligns with the preferred efficacy level in the WHO Target Product Profiles^29^. We also consider scenarios for a vaccine that reduces risk of infection only, with no additional protection against severe disease, and a vaccine that reduces the risk of severe disease only, without reducing the risk of infection. In simulations exploring the potential for lower vaccine efficacy in older age groups, we also consider a scenario where vaccine efficacy in those aged 65 years and older is 50% of that in younger individuals, based on observations from influenza vaccination^30,31^. We assume an average period between vaccination and protection of 7 days, consistent with observations of increasing antibody and T-cell responses in the phase II data^32–36^. We assume no partial protection in the period between vaccination and protection. Antibody reversion from natural infection^37^ and re-infection^38^ has now been reported, and a UK-based study of hospital staff found that prior infection was associated with an 83% lower risk of infection, with a median duration of follow-up of five months^39^. Our baseline runs therefore assume that natural immunity following infection persists for an average of one year, and we additionally explore both six-month and long-term durations in sensitivity analyses, consistent with values identified in the WHO Target Product Profiles^29^. Key parameters are summarised in Table 1, Table S1, and Table S3.

### Scenarios and Counterfactual

To evaluate the public health impact of any vaccine, we must consider the epidemic trajectory in the absence of vaccination. However, because of the vastly different experiences between countries in their exposure and response to SARS-CoV-2, considering such counterfactuals is challenging, particularly since testing rates differ markedly and so it can be difficult to accurately assess the current epidemic stages of different countries. Furthermore, whilst vaccination has commenced in some countries, scale-up will occur over a longer period. In the interim, countries will likely continue to implement some level of non-pharmaceutical interventions (NPIs). We do not explicitly model individual NPIs, but instead incorporate the assumed impact of combined NPIs in the estimate of the reproduction number R_t_.

To understand the public health value of the vaccine and consider allocation strategies that do not penalise countries for lacking the capacity to implement NPIs, in our baseline scenario we make the simplifying assumption that the vaccine is introduced while NPIs are in place. This avoids the situation in which our modelled results suggest the vaccine has no impact because the vaccine introduction “misses” the counterfactual epidemic in the year being analysed. NPIs are then lifted (i.e. R_t_ increases) after the target vaccine coverage is achieved, allowing vaccine impact in a fully vaccinated population to be compared to the counterfactual scenarios. Under this simplification, we consider a scenario where the first dose of a vaccine is introduced from the beginning of 2021 over 30 days, and that a second vaccine dose occurs 21 days after the first, allowing target coverage to be achieved by the end of February 2021 and with no further vaccination taking place subsequently (although in practice if vaccine-induced immunity wanes, then repeat vaccination is likely to occur). We then compare scenarios from March 2021 onwards and quantify vaccine impact over the time horizons of 2021 only and 2021–2022. In sensitivity analyses we also consider a more realistic coverage scale-up throughout the year.

Our baseline scenarios assume that an initial peak in transmission occurred in the first half of 2020, which is reduced by NPIs, resulting in lower levels of transmission through the remainder of the year (Figure S3). Assuming a mean duration of immunity of one year and for levels of transmission where R_0_ = 2.5 and R_t1_ = 1.1, the simulation results in 11% of the population becoming immune by the end of 2020 in a HIC setting (plausible given seroprevalence surveys to date^23,24^), and results in 11%, 12%, and 16% immune in UMIC, LMIC and LIC settings respectively (Figure 1; Figure S5). We explore additional scenarios with long-term natural immunity, which results in a higher proportion immune at vaccine introduction (Figure S5), and also explore different plausible values of R_0_ and R_t1_, which result in the proportion of the population immune at vaccine introduction varying between 4% and 24% (Figure S6).

**Figure 1:**
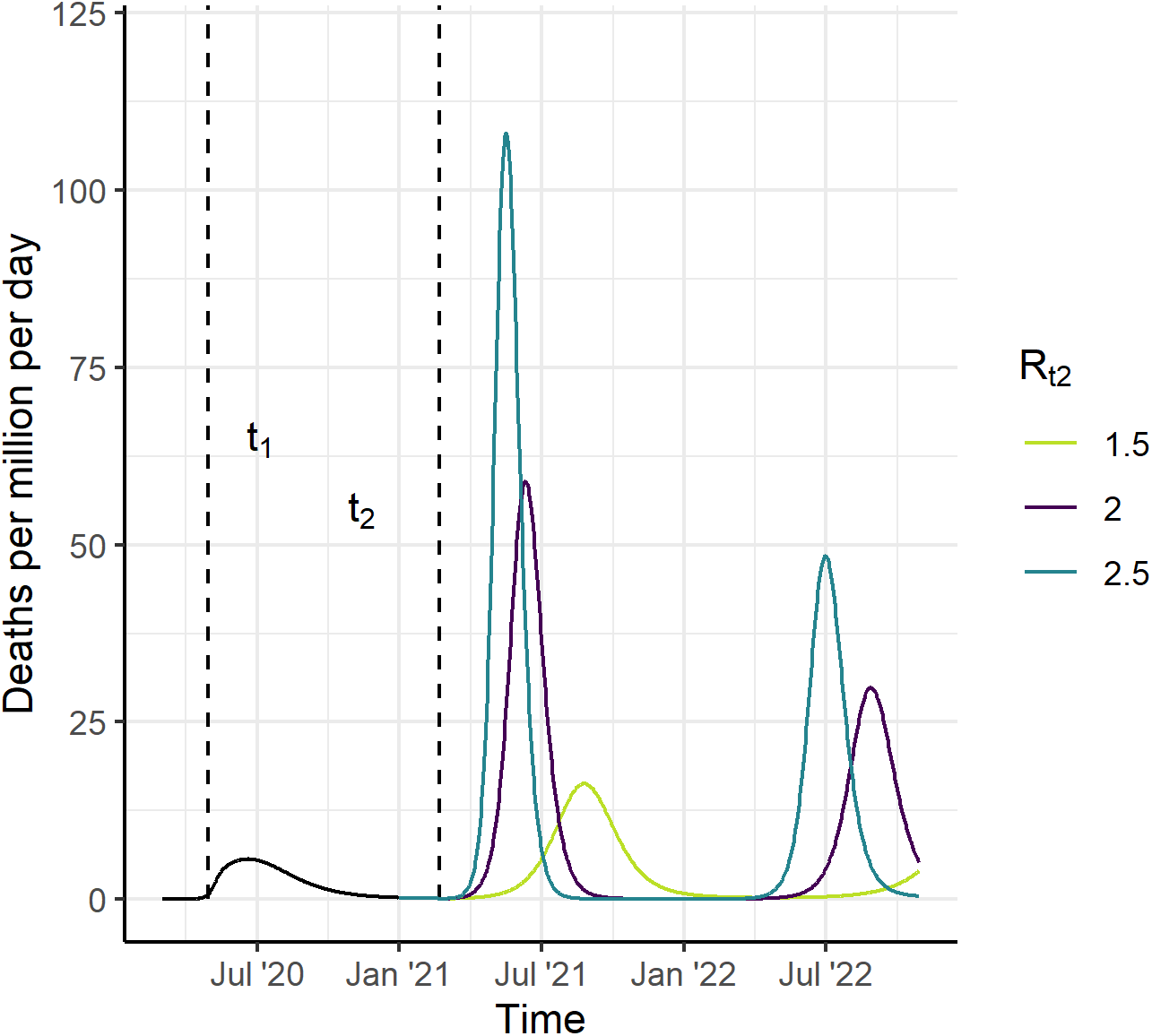
Scenarios for the Course of the Epidemic from 2020–2022, for a High-Income Country Setting, in the Absence of a Vaccine (counterfactual scenarios). We assume that R_0_=2.5 up to time t_1_ (May 2020) and that R_t1_ drops to 1.1 between time t_1_ and t_2_ (February 2021). Assuming an average duration of naturally acquired immunity of one year, this results in 11% in the recovered (immune) state at vaccine introduction. From time t_2_ onwards, we consider three counterfactual scenarios, R_t2_=1.5, 2 and 2.5 shown in light green, purple, and turquoise, respectively. Vaccine impact is compared to these counterfactual scenarios.

For each income setting, we consider two possibilities for health system capacity. The first is that all health systems are unconstrained – and hence that a constant (age-dependent) proportion of infections are hospitalised and receive appropriate care, regardless of the size of the epidemic. This results in population-level IFRs that are highest in HIC settings and lowest in LIC settings given the different demographics of these populations^15^. Second, we more realistically assume that health systems will be constrained to varying degrees. Here we follow the assumptions and parameters in Walker et al.^15^ in which LIC and LMIC settings have limited hospital capacity (estimated using World Bank data) and that once exceeded, those requiring hospitalisation but who do not receive it experience worse outcomes. In contrast, in UMIC and HIC settings, while existing hospital capacity may also be exceeded, we assume that surge capacity is implemented to fill this gap. In LIC settings we additionally assume that outcomes for hospitalised cases are worse than in other settings. This results in slightly higher population-level IFRs in LIC and LMIC compared to UMIC and HIC settings^15^. Our default assumption is that health system constraints are in place.

It is unlikely that relaxing NPIs would result in a return to the high level of transmission (R_0_ between values of 2 and 4) seen before controls were introduced, due to ongoing interventions (e.g. the use of facemasks, working from home and test/trace/isolate)^40^. We therefore explore three scenarios for R_t2_ from March 2021 onwards: R_t2_=1.5, 2, or 2.5. These values were chosen to represent a range of epidemic trajectories in the absence of vaccination, with the higher R_t2_ resulting in a rapid epidemic and the lower R_t2_ values resulting in flatter but longer epidemics. We calculate the deaths averted and life-years gained as the differences in deaths and life-years lost between the vaccinated and counterfactual scenarios. The life-years lost is estimated by summing the remaining life expectancy for each death in each age group, where the reference life expectancy is fixed at 86.6 years^41^, consistent with the approach advocated for ethical vaccine allocation^42^.

### Vaccine Allocation

We first consider optimal vaccine allocation within a country under different supply constraints. To do this, we divide the population into 5-year age groups and generate all possible age group combinations, where the selected age groups would then be allocated the vaccine. Since it is unlikely that multiple non-sequential 5-year age groups would be selected in any vaccine programme (for example, vaccinating only the 20–24-, 40–44- and 60–64-year-olds is not programmatically likely), we retain the age group combinations in which up to two contiguous “bands” of age groups could be selected. This would allow, for example, the elderly and children to be selected. The full set of age group combinations is described in the Supplementary Information (Section 1.6; Figure S3). For each combination we calculate the deaths averted over the time horizon, assuming 80% coverage (or uptake) of the vaccine within each 5-year age group and assuming default vaccine efficacy, duration, and coverage parameters, and with a 2020 epidemic that results in 11% of the HIC population being immune when the vaccine is introduced (Table 1). The vaccine supply is calculated as the percentage of the total population who is vaccinated. We then select the most efficient allocation frontier from this set of simulations (see Supplementary Information, Figure S11). We compare this optimal strategy to two age-targeted approaches. In the first approach we sequentially allocate from the oldest age group downwards (80+ years, 75+, 70+ etc.), for both uniform vaccine efficacy across age groups and where efficacy in the 65+ group is reduced by 50%. In the second approach we target the working-age population, beginning with the highest risk group (60–64 years) and working downwards by age, and then sequentially add in the younger and older age groups either side of the working age population until the entire population is covered. We repeat this analysis for each income setting.

Second, we explore the impact of different global vaccine dose allocation strategies, assuming that the dose supply is constrained in 2021, with the dose constraint guided by the two billion doses that will be made available by the end of 2021 through the COVAX facility^43^. We use the same simulations as above to calculate the deaths averted in 2021 assuming default vaccine efficacy, duration, and coverage across the four income settings and for the set of 5-year age group combinations (Supplementary Information Section 1.6). We then calculate the total population-level impact in terms of deaths averted in the year 2021 for that income setting and age group, and use these results to quantify the total global impact and number of vaccine doses required for every combination of age-targeting strategies across the four income settings.

We then identify six plausible global vaccine allocation strategies, and simulate the health impact for each strategy, assuming a two-dose schedule, a 2 billion dose constraint, and 15% buffer stock and wastage (Table 1):

- Strategy A: Countries are allocated doses relative to population size.
- Strategy B: Countries are allocated doses relative to population size, with individuals 65 years and older targeted first.
- Strategy C: Countries are allocated doses relative to size of population 65 years and older, with that age group targeted first.
- Strategy D: High-income countries can access doses first.
- Strategy E: Low- and lower-middle-income countries can access doses first.
- Strategy F: Countries are allocated the first 2 billion doses relative to population size, with additional doses available to high- and middle-income (UMIC and LMIC) countries (1.15 billion and 1.10 billion respectively, again distributed according to population).

For these six strategies, we do not stratify the population within an income setting into individual countries, therefore by extension we assume that doses can be distributed across any number of countries within an income setting, even if the dose supply means that coverage within an individual country is low.

We additionally use the ompr package in R^44^, which formulates and solves mixed linear integer optimisation problems, to identify the optimal global vaccine allocation strategy. To do this, we stratify the global population into the population sizes of the 85 largest countries (comprising 95% of the global population), assigning each country the characteristic age distribution, contact matrix, and health system capacity for its income setting to reduce computational requirements. Under this optimisation algorithm, the available doses can be allocated both between and within countries, thereby allowing different combinations of 5-year age groups to be selected within each country.

For both the fixed and optimal global vaccine allocation strategies, we calculate the total deaths averted per million population, and the deaths averted per 100 fully vaccinated persons, in order to compare the efficiency of the identified strategies.

### Sensitivity Analysis

We identify the optimal within-country and global between-country vaccine allocations, and calculate the impact of the six fixed global strategies (A–F), while varying our default parameter assumptions: vaccine efficacy reduced from 90% to 70%; efficacy in the 65 years and older population reduced by 50%; mode of vaccine action as disease-reducing only; greater NPIs following the introduction of a vaccine (implemented as R_t2_ reduced from 2.0 to 1.5); reduced level of NPIs following the introduction of a vaccine (or higher transmission, implemented as R_t2_ increased from 2.0 to 2.5); health system constraints assumed to be absent; infectiousness in children younger than 10 years reduced by 50%; and life-years gained rather than deaths averted used as the global optimisation outcome measure.

## Results

Vaccines that are efficacious against infection can have health impact in two ways: by reducing the burden of disease through direct protection of those vaccinated, and by reducing infection, and therefore onward transmission, thus providing indirect protection to the entire population. For our assumed R_0_ of 2.5, the theoretical coverage required to achieve herd immunity for a 100% efficacious vaccine is 60%, in a population that mixes randomly. For a more transmissible strain (such as the recently identified new UK variant^45,46^) with an R_0_ of 4 the coverage required for herd immunity increases to 75%. Coverage must be higher if vaccine efficacy is below 100% (Figure 2A). For example, for a vaccine with 90% efficacy the coverage would need to be 67% for R_0_=2.5 and 83% for R_0_=4. However, as demonstrated in our simulations, temporary herd immunity can be reached at lower coverage levels since ongoing NPIs generate a lower effective reproduction number (illustrated here with R_0_=2). With wider vaccine availability from 2022 onwards it is likely that NPIs would gradually be lifted and hence higher coverage would be required to sustain herd immunity.

**Figure 2:**
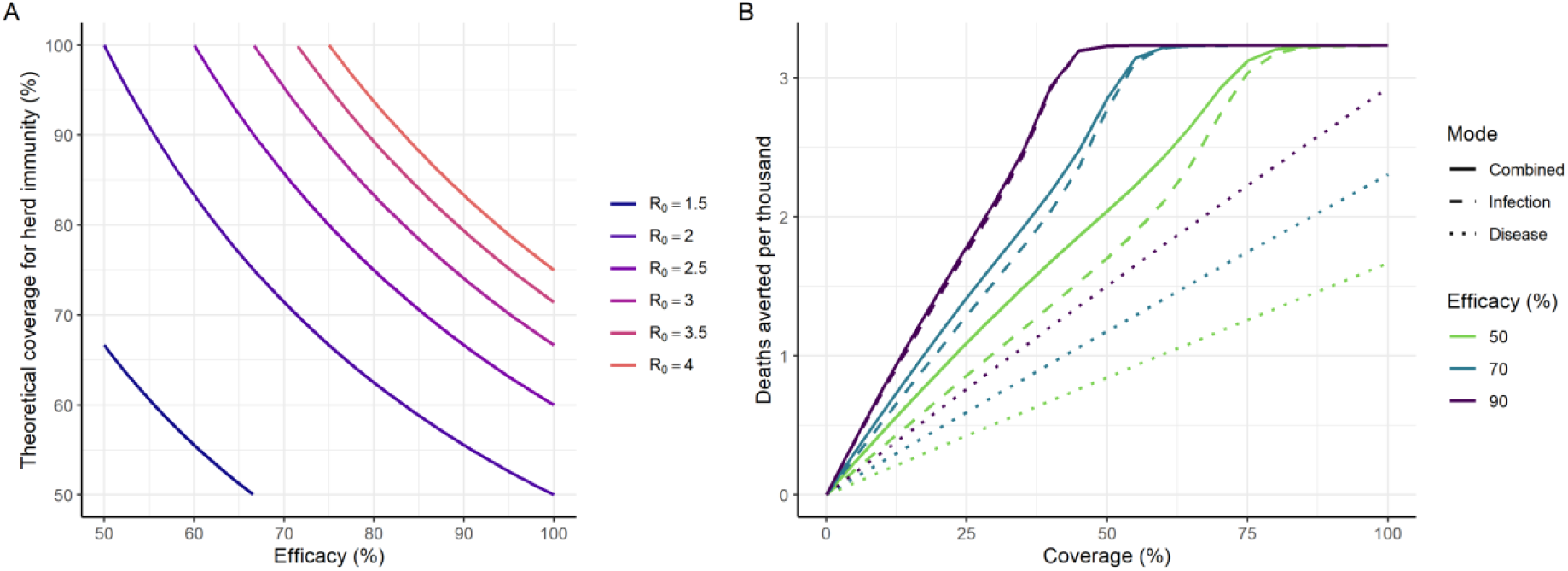
Vaccine Efficacy and Herd Immunity. (A) The relationship between efficacy of a vaccine that reduces the risk of infection, and the theoretical coverage required for herd immunity, for a range of levels of transmission shown as the level of the reproduction number R_t_. The theoretical coverage assumes random mixing of the population. (B) Projected total deaths averted per thousand population in 2021 under the default assumptions shown in Table 1 (with R_0_ = 2.5, R_t1_ = 1.1, and R_t2_ = 2.0). The colours show different magnitudes of vaccine efficacy. Solid lines represent impact for a vaccine that is efficacious against infection, with additional efficacy against severe disease. Dashed lines represent a vaccine that is efficacious against infection only, and dotted lines represent a vaccine that only prevents severe disease (and hence death) but does not reduce infection or onwards transmission (Table S3). Impact is shown for a HIC setting and all age groups are vaccinated uniformly; additional plots for other income settings and health system constraint assumptions are in Figure S7.

A vaccine that prevents infection has a greater impact than one of equivalent efficacy that prevents severe disease only due to the indirect effect of the former on transmission (Figure 2B). With an infection-reducing vaccine the projected deaths averted increases sharply with coverage until herd immunity is approached, at which point the curve plateaus. In contrast, a vaccine that only provides direct protection against disease has a linear relationship between coverage and health impact (deaths averted per thousand population). For example, for R_0_=2.5, R_t1_=1.1, R_t2_=2.0, vaccine efficacy of 90% and coverage of 67%, only approximately 60% of the deaths averted by an infection-reducing vaccine would be averted by a disease-reducing vaccine; this finding is consistent across income settings (Figure S7). The additional value of indirect protection remains important for all potential vaccine efficacies and levels of coverage, as seen in the different slopes of the solid and dashed lines in Figure 2B. Similarly, the duration of protection will affect overall impact (Figure S8) although this could be overcome through repeat vaccination.

The timing of vaccine introduction relative to the epidemic stage in each country will determine the additional health benefit that the vaccine generates. The health impact of the vaccine depends on the proportion of the population that have naturally acquired immunity at the time of vaccine introduction and the duration of that immunity. There is a clear decrease in public health impact with the extent of pre-existing immunity in the population, under the assumption that ongoing NPIs in 2021 do not differ based on this pre-existing immunity (Figure 3A–B). For example, we predict 6.0 deaths averted per 1,000 population up to the end of 2022 if only 4% of the population have pre-existing immunity compared to 3.9 if 14% of the population have pre-existing immunity. In addition, if vaccination is timely, its impact is greater in countries where other NPIs are not feasible, i.e. where R_t2_ is higher (Figure 3C–D). While our main analysis assumed that the target population is vaccinated before NPIs are lifted, if NPIs are lifted at vaccine introduction and that vaccination takes place over a longer period (one year), vaccine impact will be lower (Figures 3E–F and S9). Furthermore, a greater public health impact will be obtained by targeting the older ages first rather than the working age population because the overall vaccine coverage during the period in which the epidemic occurs is lower compared to if NPIs are lifted after the target population is vaccinated.

**Figure 3:**
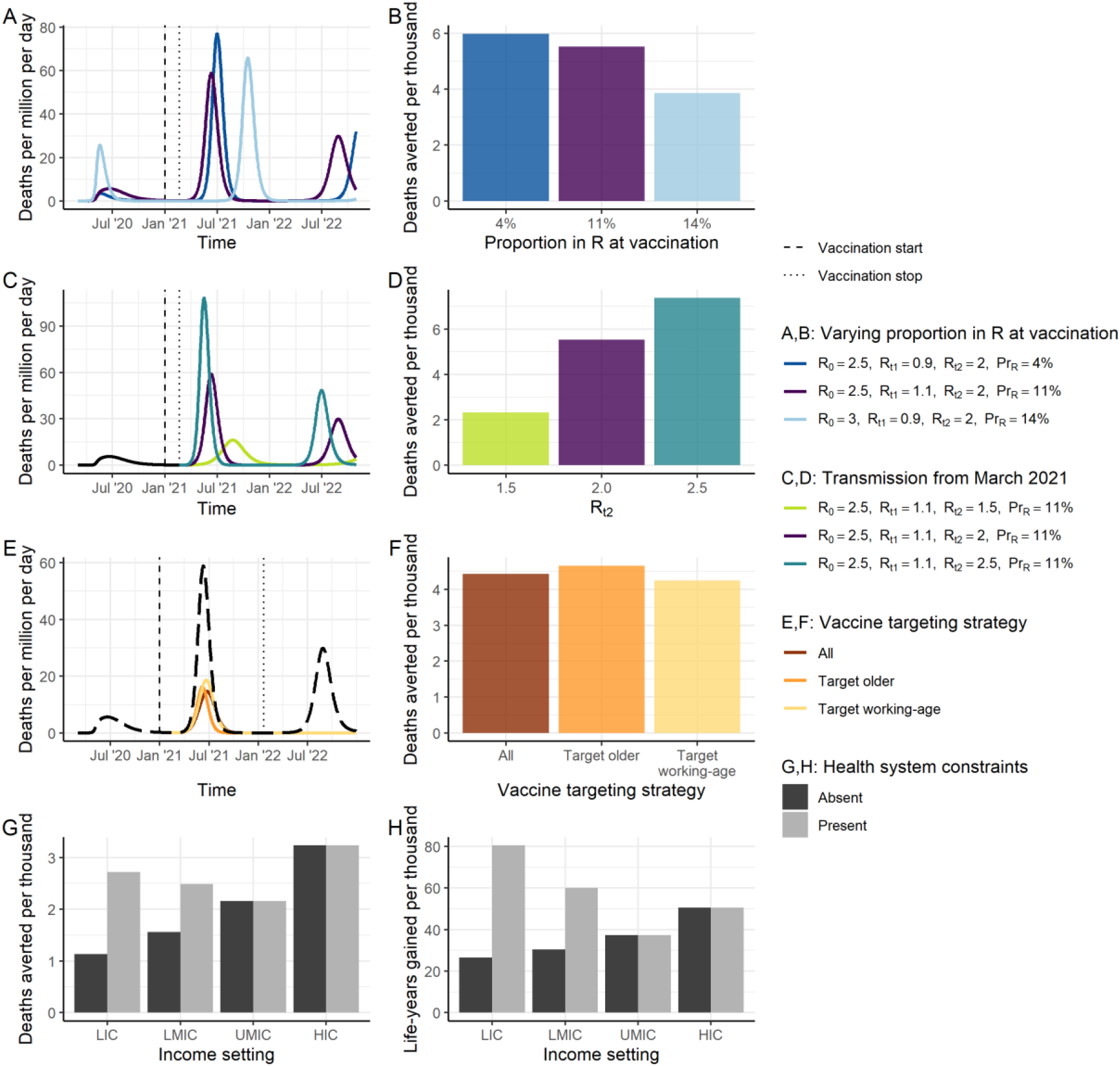
Epidemic Characteristics at Vaccine Introduction. (A) Three scenarios for the stage of the epidemic at vaccine introduction. The dark blue line shows a scenario where transmission has previously been suppressed and therefore the proportion immune at vaccine introduction is low (4%). The purple line shows the default scenario in which the proportion immune at vaccine introduction is 11%. The light blue line shows a scenario in which more widespread transmission occurred during 2020 and the proportion immune at vaccine introduction is higher (14%). (B) The projected impact of vaccination in terms of total deaths averted per thousand individuals over 2021–2022, for the scenarios in A. All other vaccine characteristics are set to the default assumptions. (C) Three scenarios for the course of the epidemic from February 2021 onwards assuming the default scenario up until this time of vaccine introduction. The light green line shows the scenario for R_t2_=1.5, purple R_t2_=2, and turquoise R_t2_=2.5. (D) Deaths averted per thousand individuals over 2021–2022, for the scenarios in C. All other vaccine characteristics are set to the default assumptions. (E) Three scenarios for the course of the epidemic from February 2021 onwards where NPIs are assumed to be lifted when the vaccine is introduced, and the target population is vaccinated at a constant rate over 2021, for three vaccine targeting strategies (coloured lines). The black long-dashed line shows the counterfactual scenario. (F) Deaths averted per thousand individuals over 2021–2022, for the scenarios in E. “All”: all age groups vaccinated simultaneously. “Target older”: the 80+ group is vaccinated first, then additional groups (75–79, 70–74 and so on) are consecutively vaccinated. “Target working-age”: the 15–64-year-old group is vaccinated first, and then the older group, and then children. (G, H) Deaths averted (G) and life-years gained (H) per thousand population in 2021 for each income setting, where health systems are either unconstrained (dark grey) or constrained (light grey). Default vaccine parameters are in Table 1.

The public health value of vaccination will also depend on the risk profile of the population and whether other therapeutic means are available to reduce morbidity and mortality. In the absence of any health system constraints, the public health value of a vaccine is predicted to be greatest in HIC since these countries have the largest elderly populations (Figure 3G–H). For example, we predict 3.2 deaths averted per 1,000 population in a HIC compared to 1.1 in a LIC in 2021. However, once health system constraints are incorporated into the model in lower-income settings, the public health value of the vaccine becomes more similar across settings. Furthermore, if the metric for assessing public health value accounts for the age at death (i.e. life-years gained) we predict a further shift towards benefiting the lowest income settings (Figure 3H), – with 81 life-years gained per 1,000 population in a LIC compared to 50 in a HIC in 2021.

Even accounting for differences in demography, contact patterns and health system capacity, vaccination targeting those at highest risk of death is beneficial across all income settings. Given the strong age-gradient in the estimated IFR for SARS-CoV-2, here we illustrate the value of targeting by age (noting that similar principles will apply to other identified risk factors or vulnerable groups). Under a strategy in which the most vulnerable age groups are targeted first, the required number of vaccine doses differs between income settings. This reflects the different age distributions, with relatively fewer doses required to fully cover the elderly populations in lower-income settings (Figure S10). The predicted public health impact of such a strategy is also highest in HIC, LMIC, and LIC settings, reflecting the overall higher IFR experienced in HIC countries and the presence of health system constraints in the LMIC and LIC settings. However, the impact of age prioritisation is substantially lower if the vaccine is less efficacious in the elderly population (Figure 4B, 4D, 4F, 4H, Figure S13, Table S5).

**Figure 4:**
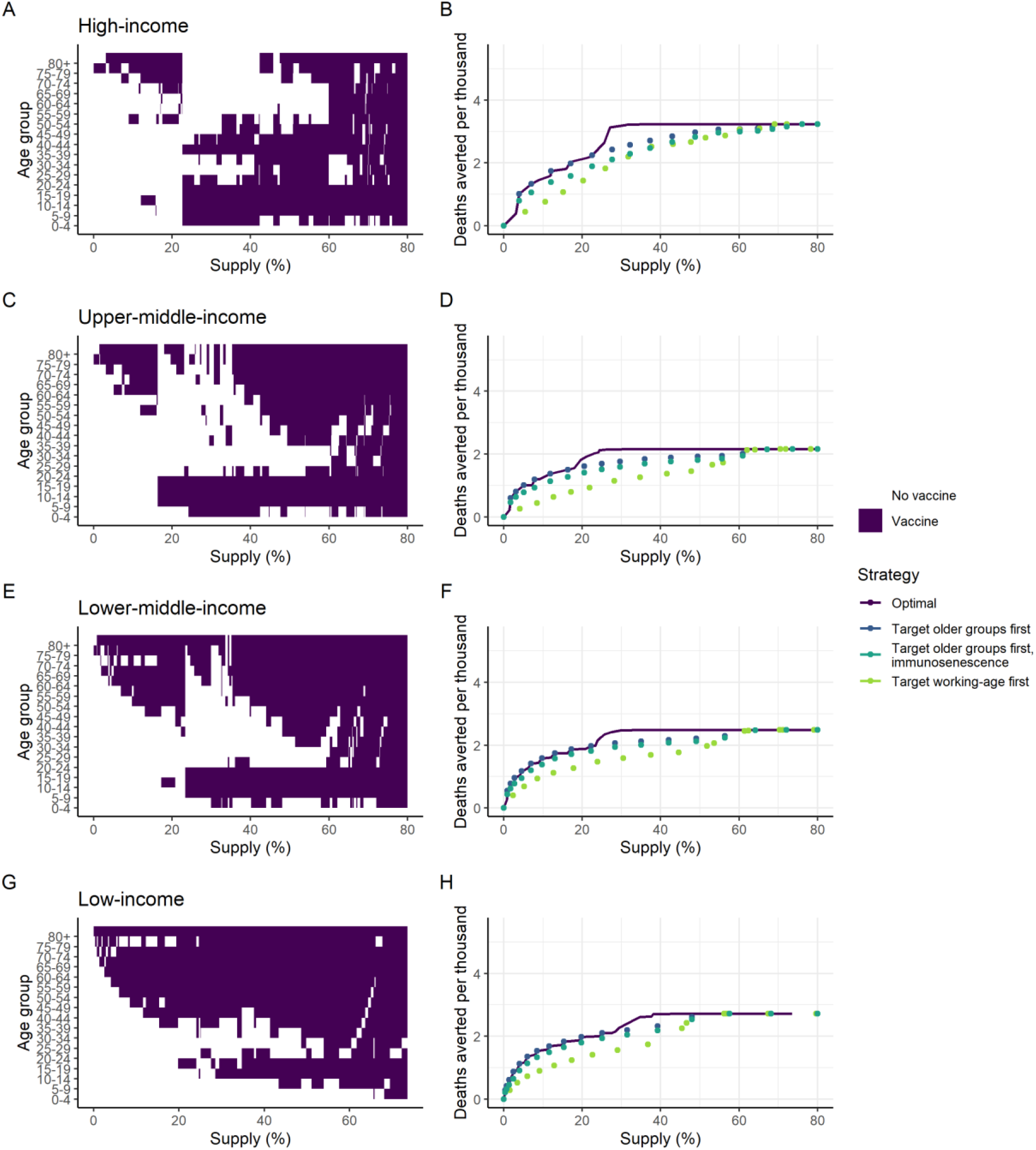
Age-targeting of Vaccine Introduction. These panels illustrate the most efficient allocation under different supply constraints, where the supply is defined as the proportion of the total population able to access two doses. Panels A, C, E and G show the age groups allocated the vaccine under the optimal strategy for different levels of vaccine supply, where the purple shaded regions indicate the age groups prioritised. Panels B, D, F and H show the total health impact expressed as deaths averted per thousand population as a function vaccine supply. The optimal strategies from the left-hand panels are shown in purple on the right-hand panels. The dark blue points show the strategy that prioritises the older at-risk age: 80+ for the lowest coverage level, and sequentially including additional age groups (75–79, 70–74 and so on) as additional doses are available. The turquoise points show the same strategy, but where vaccine efficacy in the 65+ population is 35% (immunosenescence). The green points show the strategy that prioritises the working age population first (beginning with the 60–64 age group and sequentially adding younger groups), then vaccinates the elderly and children as doses become available. Health system constraints in LICs and LMICs are assumed to be present These allocations are generated using the default vaccine characteristics in Table 1, with 80% coverage in the target age group vaccinated; additional scenarios are shown in Figures S12–S19.

Figure 4A–H shows the most efficient allocation of the vaccine for each of the four country income settings. In all settings, if doses are limited, the most efficient approach is to vaccinate the most vulnerable elderly population, starting at the oldest age group and working downwards. In LIC settings, this approach broadly remains optimal throughout. However, in HIC, UMIC and LMIC settings we find that above a given threshold for dose availability and for a vaccine that reduces the risk of infection, the optimal allocation strategy switches from “direct protection of the vulnerable” to one of “herd impact”, whereby the vaccine is allocated to younger populations (including children and adults). Under such a scenario, transmission is reduced in the wider community and this indirectly reduces the risk to the vulnerable elderly population to a greater extent than through direct protection. Assuming that targeted age groups are vaccinated at 80% coverage, this switch occurs when there are sufficient doses to cover between 20% and 40% of the total population, although the precise value depends on both the relative size of the elderly population and on mixing patterns between the older population and the general population. Furthermore, in UMIC and LMIC settings we obtain a “mixed” approach at these intermediate dose availability levels, with both the highest risk elderly and younger populations included. This is due to greater mixing between the older and general populations in these settings, as well as the size of the high-risk older populations, which are generally smaller than in HIC settings. In all settings our optimisation includes the vaccination of children when this switch occurs.

Our model does not explicitly capture a lower transmission risk of children; if this is included, we find that they are de-prioritised (Figure S18). The most efficient strategy is also sensitive to vaccine efficacy (where for a lower efficacy, the level of vaccine supply at which a herd impact strategy becomes optimal is higher, Figure S12). For a vaccine that reduces the risk of severe disease only, the optimal strategy is to directly protect the most vulnerable (Figure S14). If NPIs are maintained at a greater level following vaccine introduction (explored by reducing R_t2_ from 2.0 to 1.5), then we find that younger populations are targeted at a lower dose supply threshold (around 10%, Figure S15). Conversely, if transmission is higher (R_t2_ increased from 2.0 to 2.5), it is efficient to continue to target older age groups even at a higher level of dose availability (Figure S16).

At a global level and assuming an initial vaccine dose supply of 2 billion, allocating doses equitably across all income settings relative to population size (Strategy B), or relative to population size in the 65+ age group (Strategy C), are the most efficient of the fixed approaches considered, assuming that within a country the vaccine is targeted to the at-risk population (Table 2). This finding is consistent across all sensitivity analyses considered (Table S6). If high-income countries can preferentially obtain a large proportion of the available vaccine doses at the expense of lower income countries (and assuming health systems are constrained in LIC and LMIC settings), then we would expect an additional 900 deaths per million from this less efficient global allocation, assuming that all countries have similar levels of pre-existing natural immunity (Table 2, Strategy D). Consistent with our earlier results, the projected impact is greater if, within each country, the highest risk older age groups are vaccinated at a high level of coverage, averting 1,400 deaths per million (Strategy C), compared to averting 650 deaths per million if all ages are vaccinated at a lower coverage level (Strategy A). Under a fully-optimised global allocation – in which allocation both within- and between-countries is optimised – we estimate the most efficient strategy can avert 1.43 deaths per 100 fully vaccinated persons, marginally higher than the equitable allocation with prioritisation to older age groups scenario (1.25 deaths averted per 100 fully vaccinated persons, Strategy C).

**Table 2:** Global Allocation of Vaccine Doses for both Non-Optimised and Optimised Scenarios. The global vaccine supply was assumed to be constrained to 2 billion doses, with a two-dose schedule and 15% buffer and wastage (resulting in 0.85 billion vaccine courses available). Table S4 shows impact for the same non-optimised allocation strategies, but with the assumption that limited countries within each income setting are allocated doses at high (80%) coverage. Table S6 shows the sensitivity analysis for each of the non-optimised scenarios. FVP: fully vaccinated persons.

| Allocation strategy |  | Income setting | Target age group | Deaths averted per million | Deaths averted per 100 FVP | Total deaths averted per million global population | Total deaths averted per 100 FVP |
| --- | --- | --- | --- | --- | --- | --- | --- |
| Allocated to all countries at varying coverage | A: Countries receive doses in proportion to population | HIC | all | 831 | 0.76 | 650 | 0.59 |
|  |  | UMIC | all | 531 | 0.48 |  |  |
|  |  | LMIC | all | 730 | 0.66 |  |  |
|  |  | LIC | all | 482 | 0.44 |  |  |
|  | B: Countries receive doses in proportion to population, with 65+ group prioritised and remaining doses allocated to 15-64 age groups | HIC | 15+ | 1389 | 1.26 | 1324 | 1.20 |
|  |  | UMIC | 15+ | 1270 | 1.14 |  |  |
|  |  | LMIC | 15+ | 1374 | 1.25 |  |  |
|  |  | LIC | 15+ | 1221 | 1.11 |  |  |
|  | C: Countries receive doses in proportion to 65+ population, with 65+ group prioritised and remaining doses allocated to 15-64 age groups | HIC | 15+ | 2238 | 0.9 | 1386 | 1.25 |
|  |  | UMIC | 15+ | 1285 | 1.09 |  |  |
|  |  | LMIC | 15+ | 1237 | 1.91 |  |  |
|  |  | LIC | 15+ | 918 | 2.56 |  |  |
|  | D: Allocated first to high-income countries | HIC | all | 3234 | 0.46 | 513 | 0.46 |
|  |  | UMIC | all | 0 | 0 |  |  |
|  |  | LMIC | all | 0 | 0 |  |  |
|  |  | LIC | all | 0 | 0 |  |  |
|  | E: Allocated first to low-income and lower-middle-income countries | HIC | all | 0 | 0 | 597 | 0.53 |
|  |  | UMIC | all | 0 | 0 |  |  |
|  |  | LMIC | all | 1323 | 0.55 |  |  |
|  |  | LIC | all | 1074 | 0.45 |  |  |
|  | F: Countries receive doses in proportion to population, plus 1.15 b doses to HIC and 1.1 b doses to MIC | HIC | all | 3231 | 0.63 | 1308 | 0.56 |
|  |  | UMIC | all | 898 | 0.47 |  |  |
|  |  | LMIC | all | 1099 | 0.58 |  |  |
|  |  | LIC | all | 482 | 0.44 |  |  |
| Optimised allocation | Allocation algorithm selects countries and age groups within targets to maximise deaths averted | HIC | optimised | 3135 | 1.16 | 1672 | 1.43 |
|  |  | UMIC | optimised | 1241 | 1.44 |  |  |
|  |  | LMIC | optimised | 1581 | 1.62 |  |  |
|  |  | LIC | optimised | 1533 | 1.81 |  |  |

A fully optimised global allocation could prevent 1,700 deaths per million people in 2021. Under this strategy, a higher proportion of doses are allocated to HICs (33% of global doses) with sufficient coverage (27%) in most HICs to pursue a herd impact strategy, assuming vaccination is efficacious against infection and can be implemented while transmission remains suppressed (Table 3), relative to 28%, 33% and 6% of global doses in UMIC, LMIC and LIC settings, respectively. This is due primarily to the older populations in HIC but is also dependent on absolute population sizes in each income band. Under the optimal allocation scenario, in LIC, LMIC and UMIC settings the proportional coverage is lower (between 8–10% coverage) and hence doses are targeted to the elderly population. Overall, assuming a vaccine reduces the risk of infection rather than only severe disease, the optimal allocation of 2 billion doses balances herd impact strategies in some countries with direct protection of the vulnerable in others.

**Table 3:** Identified Optimal Global Allocation by Income Setting. The optimal allocated coverage is the fully vaccinated persons per population, and the value in parentheses represents the proportion of total global doses allocated to that income setting. The Default scenario represents the default vaccine assumptions in Table 1. We also present the sensitivity of the allocation to changes in the assumptions about vaccine efficacy, vaccine efficacy in the 65+ years age group, mode of action of the vaccine, NPIs at vaccine introduction, health system constraints, reduced infectiousness in children younger than 10 years, and life-years gained (LYG) as an optimisation measure. The within-setting results are shown in Figures S12–S19, and the public health impact for each scenario is in Table S5. The estimated proportion of the global population in each income setting is included for reference.

|  |  | Income setting |  |  |  |
| --- | --- | --- | --- | --- | --- |
|  |  | HIC | UMIC | LMIC | LIC |
| Proportion of global population |  | 15.9% | 37.3% | 38.1% | 8.7% |
| Allocated coverage of the population (proportion of global doses allocated to income setting) | Default | 27.1% (33.2%) | 8.6% (27.9%) | 9.8% (32.6%) | 8.5% (6.2%) |
|  | Lower vaccine efficacy (70%) | 32.7% (40.2%) | 7.8% (25.3%) | 8.5% (28.3%) | 8.5% (6.2%) |
|  | Reduced vaccine efficacy (scaled by 50%) in 65+ years population | 27.1% (33.2%) | 11.5% (37.3%) | 6.9% (23.2%) | 8.5% (6.2%) |
|  | Vaccine efficacious against disease only | 18.3% (22.5%) | 7.8% (25.3%) | 13.1% (43.7%) | 11.6% (8.5%) |
| | NPIs maintained at higher level following vaccine introduction (such that $R_{t2}=1.5$) | 11.3% (13.9%) | 11.5% (37.1%) | 11.2% (37.5%) | 15.6% (11.5%) |
| | NPIs maintained at lower level following vaccine introduction (such that $R_{t2}=2.5$) | 29.2% (35.8%) | 7.8% (25.3%) | 9.8% (32.6%) | 8.5% (6.2%) |
|  | Health system constraints absent | 27.1% (33.2%) | 15% (48.7%) | 4.5% (15.2%) | 4% (3%) |
|  | Reduced infectiousness in children younger than 10 years | 29.3% (35.9%) | 7.8% (25.3%) | 9.7% (32.5%) | 8.5% (6.2%) |
|  | LYG as optimisation outcome measure | 27.1% (33.2%) | 1.7% (5.5%) | 10.7% (35.8%) | 34.7% (25.6%) |

The global optimised strategy is sensitive to assumptions about the vaccine mode of action, with a more equitable allocation found to be optimal for a vaccine that is efficacious against disease only (Table 3). For a scenario with higher transmission following vaccine introduction relative to the default assumption, additional doses are allocated to HIC countries, however, for a scenario where NPIs are maintained to a greater extent following vaccination, a more equitable global allocation is optimal as the difference in averted deaths between income settings is less marked (Table 3, Figure S20). We also find that if life years gained, rather than deaths averted, is used as the outcome measure, doses are diverted away from UMICs and allocated to LMIC and LIC settings, whereas removing assumptions about health system constraints causes doses to be allocated preferentially to HIC and UMIC countries (Table 3, Figures S17 and S19).

## Discussion

An effective vaccine against SARS-CoV-2 will have enormous global public health and economic value. As the leading candidate vaccines progress further through late-stage clinical trials and receive approval from government regulatory authorities, preparation is underway to scale up manufacturing capacity, supply chains, and delivery systems. This preparation should enable the first vaccines to be rapidly distributed in the coming year, however, it is unlikely that any candidate vaccine could be manufactured and distributed in sufficient quantity in 2021 to enable all countries to vaccinate their populations to achieve herd immunity. Furthermore, evidence on the extent to which the current vaccines are efficacious against infection (and therefore have potential to generate indirect protection) is still emerging from implementation studies, as phase III trials are generally not designed to measure this property. Current COVAX plans favour a global allocation strategy that prioritises the highest risk groups – including the elderly – and suggest an “equitable” vaccine allocation strategy in which each country receives doses in proportion to their population size and their epidemic status^7,47^. Our results support such a strategy being close to optimal in terms of reducing the potential global mortality from SARS-CoV-2. Once all countries have been allocated doses to vaccinate 20% of the population, COVAX currently proposes that allocation of further doses should occur based on the epidemiological situation and local vulnerabilities. While we did not specifically explore this, our modelling framework can help support future prioritisation in the coming months.

For most infectious disease vaccines, one of the underlying goals is to vaccinate a sufficiently high proportion of the population to achieve herd immunity. Our results demonstrate that such indirect protection remains important for SARS-CoV-2 despite the strong risk-profile with increasing age. Indirect protection has a particularly important benefit for vulnerable groups – such as the immunocompromised – who cannot be directly protected through vaccination. However, at a population level, the reduction in transmission that occurs also benefits those that receive the vaccine but who for whatever reason do not achieve sterile protection. The coverage required to achieve herd immunity will depend on both the underlying transmissibility of the virus (R_0_) and vaccine efficacy (the effective coverage) and requires a vaccine that is efficacious against infection rather than only disease. For our assumed R_0_ of 2.5, the theoretical coverage required is 60% for a 100% efficacious vaccine. Thus, for a vaccine efficacy of 90%, population coverage of nearly 70% would be required. It is possible that heterogeneity in contact rates could reduce this theoretical coverage if vaccination is targeted to those with the highest contact rates^48,49^. Furthermore, in the presence of ongoing NPIs, the effective coverage required to interrupt transmission (i.e. to reduce R_t_ to less than 1) will be lower. Nevertheless, aiming for high coverage will clearly be important given the uncertainty in the precise value of this threshold.

One of the challenges with quantifying the public health value of a SARS-CoV-2 vaccine is determining the appropriate “counterfactual” scenario against which its value can be assessed. Here we chose to generate some simplified scenarios that illustrate three factors determined by the epidemic to date: the proportion of the population with naturally acquired immunity, the likely forward trajectory of the epidemic in the absence of a vaccine, and the duration of both naturally acquired and vaccine-induced immunity. The interaction between these factors is predicted to generate complex immune landscapes^50^. The duration of vaccine-induced immunity is perhaps best addressed through repeat vaccination; however, it is important to note that such data will only become available after several months, and possibly years, of follow-up of trial participants. Similarly, pre-existing naturally acquired immunity is unlikely to be a barrier to immunisation, provided there are no vaccine-associated risks from antibody-dependent enhancement^51^. Thus, the value of vaccination should be quantified by the degree to which it enables NPIs to be lifted. Given the broader economic and societal costs of NPIs, combined epidemiological-economic models are required to address this.

We focused on two metrics to capture the public health value of the vaccine: deaths averted, and life-years gained. These metrics were chosen because the IFR is the best characterised endpoint and is easily translatable between settings. However, the impact of SARS-CoV-2 in terms of longer-term morbidity – so-called “Long COVID”^52^ – could also be considerable. In health economics longer-term morbidity would normally be captured via Quality-Adjusted Life Years (QALYS). If morbidity is significant then it could favour the prioritisation of vaccination towards younger age-groups. However, at the current time, the extent and longer-term consequences of COVID-19 remain relatively poorly understood, with no other comparable respiratory infections available with which to quantify QALYs. This will be a research priority in the coming months and years.

Several risk groups for more severe outcomes following SARS-CoV-2 infection have been identified, including individuals in some minority ethnic groups (although such studies are currently biased towards countries with a majority White population)^53^; those with obesity, who have a higher risk of a COVID-19 positive test, hospitalisation, and death^54^; and those with other underlying health conditions^55^. However, age remains one of the strongest determinants of increased risk of mortality^56^. Our results demonstrate that targeting a limited vaccine supply to older age groups is likely to be an efficient way from a public health perspective to reduce mortality if the number of available doses is insufficient to adopt a herd impact approach. However, even if sufficient doses of a vaccine that is efficacious against infection become available in 2021 to target vaccination to the working-age population to reduce transmission, if the early supply in the first quarter is constrained then targeting the most vulnerable will likely remain optimal. This result is consistent with other modelling studies that have considered either a full optimisation^57,58^ (in which a similar trade-off between direct protection and herd impact is observed) or sequential efficiency^59,60^ (in which the most vulnerable are initially prioritised). Such an approach also has the advantage that delivery systems are in place in many (though not all) settings that can access such groups. It is possible that for some vaccines the immune response in older individuals could be weaker; more data are required to characterise this effect for some products^2^. However, both the Pfizer/BioNTech and Moderna phase III trials demonstrated high efficacy in older age groups, providing further impetus for vaccinating older groups^4,5^.

One of the most difficult challenges facing decision-makers in 2021 will be the allocation of a limited dose supply. At the global level, under our baseline assumptions, the strategy that maximises the total deaths averted is one in which the available vaccine doses are allocated preferentially to higher-income countries who have the largest at-risk elderly populations but also have the strongest health systems. However, the optimality of this allocation is sensitive to many assumptions and will depend on both the vaccine characteristics and on the stage of the epidemic in each country throughout vaccine introduction. Considering the optimisation framework applied, it is possible that there are other dose allocation strategies that would be close to the optimal solution, but that distribute doses between countries in the same income setting differently. Given this uncertainty, allocating vaccine doses according to population size appears to be the next most efficient approach.

Within a country, the most efficient strategy for individual countries will generally be to prioritise vaccination of the elderly and other high-risk groups, particularly if doses are only available for less than approximately 20% of the population. If sufficient doses are available and if vaccination reduces the risk of infection, targeting the working age population, or a combination of older and working age groups, could be more efficient in reducing mortality, compared to sequentially vaccinating the oldest to youngest age groups. This finding is consistent with other modelling studies of vaccine allocation within a single country^57,61–63^. However, while such a strategy would favour economic and social recovery, it would rely on all vaccine doses being immediately available and able to be administered while NPIs are maintained, therefore the optimality of targeting younger groups may not hold if vaccines are delivered in a gradual phased approach. Also, this approach requires a higher dose supply if transmission is higher, indicating that in the presence of new variant strains, prioritising vaccination of the most vulnerable groups will likely remain the most appropriate strategy.

In our study we assessed the impact of vaccination for broad income strata, rather than individual countries. Decision-making at the individual country level will need to account for local epidemic history, levels of transmission, other control measures and how long they can be sustained, and the sizes of the populations at highest risk within those settings^55^. Some countries have identified prioritisation frameworks for allocating vaccines, with general agreement that frontline health care workers and high-risk populations should be allocated the first available doses, and our user-friendly COVID-19 vaccine simulation interface could help in supporting such decisions about country-level introductions (Box 1). However, there are many other setting-specific factors that will also be important for governments to consider, including supply chains, logistics, access to populations, costs and budgets, and competing health, economic, and social priorities^14^.

As with any modelling study, there are several limitations to our approach. First, while phase III trial efficacy results have been announced for several vaccine candidates, durability data are not yet available, and data on the extent to which the current vaccine products can prevent infection are still emerging^2,3,28^; our model assumptions may therefore need to be updated as trials and effectiveness studies progress. Second, the number of doses and the timing of their availability are also uncertain; for this reason, we have illustrated simplified scenarios in which vaccination occurs over a one-month period for a range of epidemic stages. In practice, each country will have experienced a different epidemic when the first vaccine is introduced and will scale up coverage over a period of months. Third, the model used here is relatively simple in structure and can only simulate the impact of a single vaccine product, with one value for average vaccine efficacy, meaning we could not include complexities such as multiple vaccine products, nor the potential partial efficacy following the first dose in a multi-dose vaccine schedule. These considerations will be important for future studies. Fourth, our study focuses only on the health benefits of vaccination. It will also be important to consider other therapeutic interventions, as well as the capacity of countries to suppress transmission using NPIs, and to better capture specific risk groups as appropriate to individual countries. Furthermore, the direct health outcome is only one dimension; it will be important in the near-term to capture the impact of vaccination on non-COVID-19 health, as well as to integrate epidemiological and economic models to evaluate the impact of different vaccine allocation strategies on the economic outputs of countries and the livelihoods of their citizens.

Research and development of a SARS-CoV-2 vaccine has taken place at unprecedented speed, with efficacy and safety results now available for several vaccine candidates within one year of the pandemic being declared. Our results demonstrate that the global public health value of the vaccine can be maximised by ensuring equitable access. Acting collectively in this way during the early stages of vaccine deployment remains the ethical approach to take, even if this is not the most beneficial short-term strategy from a national perspective.

## Data Availability

The open-source model code is available as an R package at https://github.com/mrc-ide/nimue, and the code to perform the analysis and generate the tables and figures, is available to download at https://github.com/mrc-ide/covid_vaccine_allocation.

https://github.com/mrc-ide/covid_vaccine_allocation

## Author contributions

ABH, PW and ACG designed the study with input from FM, KAMG, KH, MB, and NMF. ABH, PW, GC and OJW coded the model, ran the simulations and undertook the analysis with support from CW, NB, PGTW, and RV. AH, EK, JAL, and LW provided additional technical assistance with the model development and parameterisation. ABH, PW and ACG produced the first draft of the manuscript with additional input from AL, CW, DJL, KAMG, FM, KH, MB, NMF, OJW, and PGTW. All authors approved the final draft.

