## Supplementary file for "Within-country age-based prioritisation, global allocation, and public health impact of a vaccine against SARS-CoV-2: a mathematical modelling analysis"

### 1 Mathematical model

#### 1.1 Description

We extended an age-structured deterministic SEIR compartmental model of SARS-CoV-2 transmission to capture the impact of different vaccination schedules. The main model captures the infection status of individuals as being in one of nine states (Figure S1):

- S = uninfected and therefore **susceptible** to infection
- E = **exposed** to infection but not yet infectious
- $I_{MILD}$ = **infected** and infectious with mild infection that does not require hospitalisation (this includes both symptomatic and asymptomatic infection)
- $I_{CASE}$ = **infected** and infectious with disease that will require hospitalisation
- $I_{HOSP}$ = cases that have been **hospitalised** in a general ward bed
- $I_{ICU}$ = cases that have been admitted to **an intensive care unit (ICU)**
- $I_{REC}$ = cases that have been stepped down from ICU into a general ward bed for **recovery**
- D = cases that have **died**
- R = infections and cases that have **recovered** and are immune to re-infection

Those that are susceptible become infected at a rate that depends on the level of infection in the community (i.e. the number of people in states $I_{MILD}$ and $I_{CASE}$), the transmission probability and an age-stratified mixing matrix which captures the relative number of contacts that each age group has with their own age group and other age groups. This mixing matrix, along with the demography of the population, is obtained from studies that are representative of each of the four different World Bank income groups. Given the other model parameters, the transmission probability is varied to generate a different basic reproductive number, $R_0$. Following infection, cases proceed as shown by the arrows with the durations in each state and the probability of each pathway described below (see Tables S1 and S2). Following recovery, we allow individuals to lose naturally acquired immunity and therefore return to the susceptible state. Additional constraints are included in the hospitalisation pathway to capture situations in which the need exceeds capacity; with those that do not receive appropriate care experiencing higher death rates.

The process of vaccination is captured by mirroring the basic transmission model to also capture different states of vaccination (Figure S1):

- $v_0$ – unvaccinated
- $v_1$ and $v_2$ – vaccinated but not yet protected, reflecting the two-dose vaccine schedule and need to wait approximately 28 days from dose 1 for protection to develop
- $v_3$ and $v_4$ – vaccinated and protected
- $v_5$ – previously vaccinated but no longer protected, used to capture waning vaccine efficacy.

If the vaccine is not 100% efficacious, then in the protected states we assume there is partial efficacy. For example, if the vaccine is 90% efficacious then the risk of infection in states $v_3$ and $v_4$ is 90% lower than in state $v_0$, $v_1$, $v_2$, and $v_5$. We assume that vaccine efficacy is the same for susceptible and immune individuals. For those protected by both vaccine-derived and naturally acquired immunity, we assume that the most protective effect is dominant. For example, if natural immunity lasts for one year and vaccine-derived immunity for five years then the period of protection for a recently infected and now immune individual who receives the vaccine would be one year of full naturally acquired protection, followed by four years of vaccine-derived (leaky) protection.

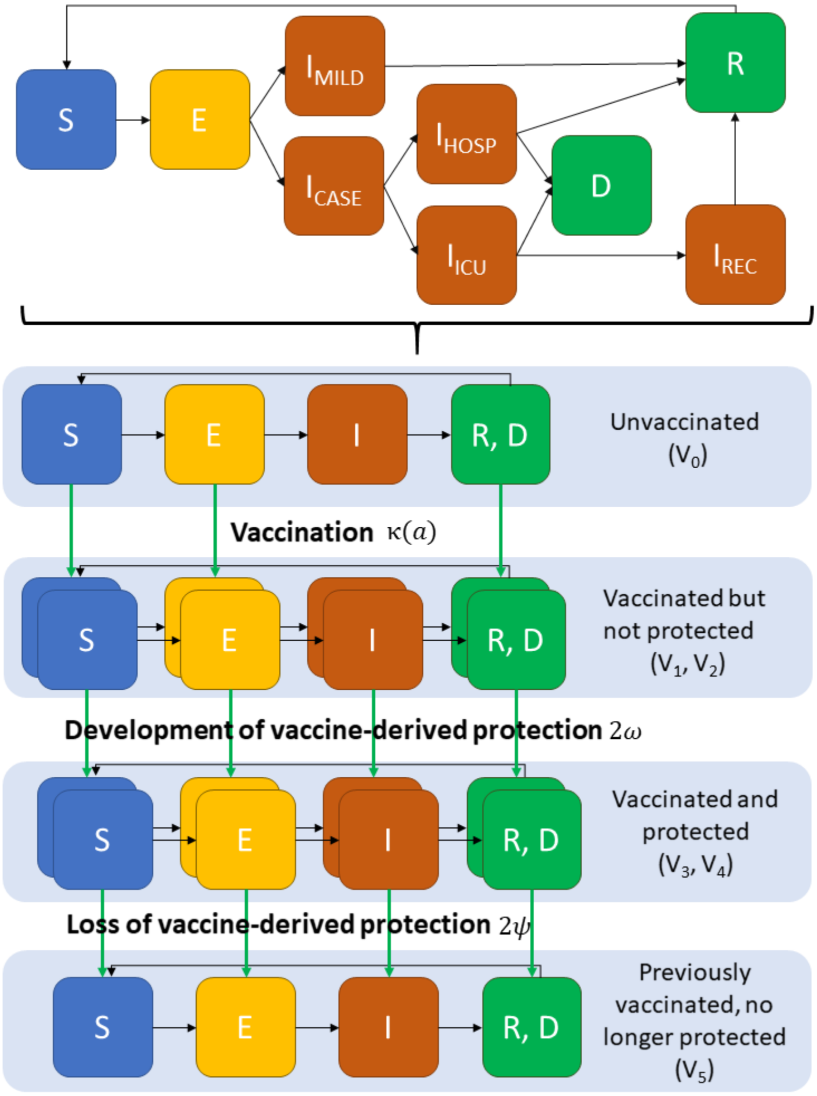


***Figure S1: Schematic of the SARS-CoV-2 transmission model.*** *The schematic in the upper section is based on the model in Walker et al.*[1] *Individuals in the susceptible (S), exposed (E) and recovered (R) compartments can be vaccinated. Vaccination status is stratified into 6 categories – those who are unvaccinated ($v_0$), those who have recently been vaccinated but are not yet protected ($v_1$ and $v_2$) and those who are vaccinated and protected ($v_3$ and $v_4$) and those that have previously been vaccinated but are no longer protected ($v_5$). Protection may refer to partial protection. We do not model revaccination of previously vaccinated individuals, due to all vaccination occurring within a one-month period. Pathways for receiving healthcare for the proportions of the population in the $I_{HOSPITAL}$ and $I_{ICU}$ compartments are described in Walker et al.* [1]

#### 1.2 Formal mathematical details

We stratify the population by age, a, and vaccination status, v. For the latter, we incorporate six categories – those who are unvaccinated ($v_0$), those who have recently been vaccinated but are not yet protected ($v_1$ and $v_2$) and those who are vaccinated and protected ($v_3$ and $v_4$) and those who have previously been vaccinated but are no longer protected ($v_5$). Note that we use the term "protected" here to denote partial protection because we model a leaky vaccine rather than sterile immunity.

Vaccines are targeted by age groups at a rate $\kappa(a)$. To specify prioritisation strategies, we define a vector of current vaccine coverage $C_a$ by age group $(a)$ and a matrix of coverage targets $T_{sa}$ where rows $(s)$ represent ordered prioritisation steps and columns $(a)$ the age group. For a given prioritisation step, e.g. $s = 1$, vaccination, up to a maximum specified rate expressed as the total number of vaccinees per day, is provided to all age groups that satisfy $C_a < T_{1a}$. Vaccinations are distributed to target age groups in proportion to the size of the unvaccinated populations. When all target coverages in the current prioritisation step are met, the step $(s)$ is incremented $(+1)$ and the process repeated. When all coverage targets in the final prioritisation step are met, vaccination is ceased. We assume that individuals may only be vaccinated once. Protection is provided either against infection – reducing the transmission parameter $\beta$ by a constant factor $1 - v_{inf}(a)$; against severe disease – reducing the rate of hospitalisation $\phi(a)$ by a constant factor $1 - v_{dis}(a)$; or against infection, with additional protection against severe disease in vaccinated individuals who experience breakthrough infection (combined modes of efficacy).

The transmission model within each vaccine category is identical to that described in Walker *et al.*[1] with the inclusion of reversion of natural immunity which is modelled by an additional flow from the recovered compartment back to the susceptible compartment (Figure S1). Let $S(t,a,v)$ denote the susceptible population in age group a with vaccination status v at time t , $E_1(t,a,v)$ and $E_2(t,a,v)$ two sequential latent periods, $I_{MILD}(t,a,v)$ infections that are either asymptomatic or symptomatic but do not require hospitalisation, $I_{CASE,0}(t,a,v)$ and $I_{CASE,1}(t,a,v)$ two sequential states for infections that are symptomatic and will subsequently require hospitalisation. $I_{HOSPITAL,,0}(t,a,v)$ and $I_{HOSPITAL,1}(t,a,v)$ are two sequential states for infections requiring a general hospital bed. $I_{ICU,0}(t,a,v)$ and $I_{ICU,1}(t,a,v)$ are two sequential states for infections requiring an ICU bed. $I_{REC,0}(t,a,v)$ and $I_{REC,1}(t,a,v)$ are two sequential states for hospitalised infections in general beds recovering from ICU whilst $R_1(t,a,v)$ and $R_2(t,a,v)$ are two sequential compartments capturing those that have recovered and are currently immune to re-infection, and $D(t,a,v,)$ are those that have died from the disease in age group a and with vaccination status v. We further split $I_{HOSPITAL,i}(t,a,v)$ and $I_{ICU,i}(t,a,v)$ into states i=1,2 to track those that either receive or do not receive their hospital or ICU bed respectively (1, 0 respectively depending on capacity constraints) and through these route either die or recover (0 and 1 respectively) in order to capture different durations of stay in hospital dependent on outcome. For example, the state tracking those that require a general bed, receive it and go on to die is $I_{HOSPITAL,i}(t,a,v,1,0)$ whilst the state tracking those that require a general bed, do not receive it and go on to die is $I_{HOSPITAL,i}(t,a,v,0,0)$. In the equations below we use the Kronecker Delta function $\delta(\cdot)$ to capture capacity constraints with this equal to 1 if there is capacity (Hospital or ICU) and zero otherwise. Contacts between age-classes are captured using the social contact mixing matrix where $c(a, a')$ denotes the rate of contacts between individuals in age groups a and a'. Age-dependent severity of disease is captured with an age-dependent mortality rate $\mu(a)$. Note that hospitalised individuals and those in ICU do not contribute to the force of infection.

Given that our focus is on short-term dynamics we do not model births, deaths, or aging. The group of individuals therefore represents their age in 2020. The equations for each of the six vaccination groups are given in Section 1.3. The parameter symbols, description and values are shown in Tables S1 and S2.

A mean duration in ICU of 13.3 days is reported from a study of clinical outcomes of COVID-19 hospitalised patients across 42 countries[2]. In UK data, 60.1% of those entering ICU survive with the ratio of the duration of time spent in ICU in those that die to those who survive reported as 0.75[3]. We use these two additional pieces of information to obtain the mean duration of stay in ICU for individuals who die and those who survive.

The model incorporates three age-dependent parameters – the probability of hospitalisation given infection $\phi_1(a)$; the probability of requiring ICU given hospitalisation $\phi_2(a)$; and the probability of dying $\mu(a)$ given hospitalisation (when hospital capacity is not saturated, we make the simplifying assumption that all deaths occur in hospital) (Table S2). The probability of dying given hospitalisation is obtained from estimates of the infection fatality ratio from a recent systematic review and model-based analysis of the literature[4]. Deaths in hospital occur from both those entering ICU and those in general beds, with the proportion in each determined by an age-dependent parameter (Table S2). The shape of this distribution reflects a lower proportion of elderly patients being admitted to ICU and is informed by unpublished UK data. For a UK demography, this parameterisation results in 20% of all hospitalised cases requiring ICU (consistent with the ISARIC international study reporting 18%[2]), 26% of hospitalised patients dying (slightly lower the ISARIC international study reporting 32%[2]), a case fatality ratio in ICU of 48% (compared with the ISARIC international study reporting 54%[2]) and 41% of deaths occurring in ICU (slightly higher than the ISARIC international study reporting 36%[2]).

The IFR estimates presented in Brazeau *et al.*[4] also include a breakdown in five-year age bands for those above 80 years of age. To obtain the correct IFR in the 80+ age group we weighted the IFR reported by 5-year age bands by the proportion of the 80+ population in each 5-year band for each income setting to obtain an average IFR in the 80+ age group.

*Table S1: Parameter descriptions and values.*

| Parameter | Symbol | Value | Description |
|---|---|---|---|
| **Epidemiological Parameters** | | | |
| Transmission parameter | $\beta$ | - | Calculated from $R_0$ (See Table 1) |
| Mean latent period | $\frac{1}{\alpha}$ | 4.6 days | Estimated at 5.1 days[5–7]. The last 0.5 days are incorporated in the infectious periods to capture pre-symptomatic infectivity |
| Mean duration of mild infection | $\frac{1}{\gamma_1}$ | 2.1 days | Incorporates 0.5 days of infectiousness prior to symptoms. In combination with mean duration of severe illness this gives a mean serial interval of 6.75 days[8]. |
| Mean duration of severe infection prior to hospitalisation | $\frac{1}{\gamma_2}$ | 4.5 days | Mean onset-to-admission of 4 days[9]. Values in the wider literature range from 1.2 days to 12 days[5–7,10,11]. Includes 0.5 days of infectiousness prior to symptom onset. |
| Mean duration of hospitalisation for non-critical cases if survive | $\frac{1}{\gamma_{3,1}}$ | 9 days | Median of values identified in[10–14]. |
| Mean duration of hospitalisation for non-critical cases if die | $\frac{1}{\gamma_{3,0}}$ | 9 days | Median of values identified in[10–14]. |
| Mean duration in ICU if survive | $\frac{1}{\gamma_{4,1}}$ | 14.8 days | Mean duration in ICU of 13.3 days from a study across 42 countries[2]. Ratio of duration in critical care if die: duration in critical care if survive of 0.75 and 60.1% probability of survival in ICU[3]. |
| Mean duration in ICU if die | $\frac{1}{\gamma_{4,0}}$ | 11.1 days | Mean duration in ICU of 13.3 days from a study across 42 countries[2]. Ratio of duration in critical care if die: duration in critical care if survive of 0.75 and 60.1% probability of survival in ICU[3]. |
| Mean duration in recovery after ICU | $\frac{1}{\gamma_5}$ | 3.0 days | Working assumption |
| Infection fatality ratio (IFR) | $\mu(a)$ | - | Age-dependent[4] (see below) |
| Hospitalisation proportion | $\phi_1(a)$ | - | Age-dependent[15] (see below) |
| Proportion of hospitalisations requiring ICU | $\phi_2(a)$ | - | Age-dependent[15] (see below) |
| Mean duration of naturally acquired immunity | $1/\rho$ | 365 days (default), infinite, or 183 days | [16–20] |
| **Vaccination Parameters** | | | |
| Age-dependent rate of vaccination | $\kappa(a)$ | - | Age-dependent assumption: set such that number of people vaccinated per day in each age group achieves target coverage by the end of the vaccination period |
| Mean time to develop vaccine-acquired immunity following second dose | $1/\omega$ | 7 days | [21–25] |
| Mean duration of vaccine-acquired immunity | $1/\psi$ | 5000 days (default), 365 days or 183 days | [26] |
| Efficacy against infection | $1 - \upsilon_{inf}(a)$ | - | Age-dependent (see Table S3)[27–32] |
| Efficacy against disease | $1 - \upsilon_{dis}(a)$ | - | Age-dependent (see Table S3)[28,30,33–36] |
| Dose schedule | - | 2 doses (default); 1 dose | [27,30–32] |

***Table S2: Age-dependent parameters for hospitalisation and death.***

| Age group | Proportion of infections hospitalised[15] | Proportion of hospitalised cases requiring ICU[15] | Proportion of hospital deaths occurring in ICU | Proportion of non-ICU cases dying | Proportion of ICU cases dying | Infection fatality ratio (IFR)[4] |
|---|---|---|---|---|---|---|
| 0 to 4 | 0.001 | 0.181 | 0.8 | 0.013 | 0.227 | 0.00004 |
| 5 to 9 | 0.001 | 0.181 | 0.8 | 0.014 | 0.252 | 0.00007 |
| 10 to 14 | 0.002 | 0.181 | 0.8 | 0.016 | 0.281 | 0.00011 |
| 15 to 19 | 0.002 | 0.137 | 0.8 | 0.016 | 0.413 | 0.00017 |
| 20 to 24 | 0.003 | 0.122 | 0.8 | 0.018 | 0.518 | 0.00026 |
| 25 to 29 | 0.005 | 0.123 | 0.8 | 0.020 | 0.573 | 0.00041 |
| 30 to 34 | 0.007 | 0.136 | 0.8 | 0.023 | 0.576 | 0.00064 |
| 35 to 39 | 0.009 | 0.161 | 0.8 | 0.026 | 0.543 | 0.00100 |
| 40 to 44 | 0.013 | 0.197 | 0.8 | 0.030 | 0.494 | 0.00156 |
| 45 to 49 | 0.018 | 0.242 | 0.8 | 0.036 | 0.447 | 0.00245 |
| 50 to 54 | 0.025 | 0.289 | 0.8 | 0.042 | 0.417 | 0.00384 |
| 55 to 59 | 0.036 | 0.327 | 0.8 | 0.050 | 0.411 | 0.00601 |
| 60 to 64 | 0.050 | 0.337 | 0.8 | 0.056 | 0.443 | 0.00941 |
| 65 to 69 | 0.071 | 0.309 | 0.8 | 0.060 | 0.539 | 0.01473 |
| 70 to 74 | 0.100 | 0.244 | 0.6 | 0.123 | 0.570 | 0.02307 |
| 75 to 79 | 0.140 | 0.160 | 0.4 | 0.184 | 0.643 | 0.03613 |
| 80+ | 0.233 | 0.057 | 0.15 | 0.341 | 0.993 | -* |
| 80 to 84 | - | - | - | - | - | 0.05659 |
| 85 to 89 | - | - | - | - | - | 0.08862 |
| 90+ | - | - | - | - | - | 0.17370 |

* To standardise input age groups for modelling, IFRs in 80 to 85, 85 to 90 and 90+ age groups are aggregated to the 80+ age group using country-specific demography.

***Table S3: Scenarios for modes of vaccine efficacy.***

| **Mode of efficacy scenario** | **Efficacy against infection** | **Efficacy against disease** |
|---|---|---|
| Combined (default) | 90% | Additional 60% efficacy for breakthrough infections |
| Infection only | 90% | No additional protection |
| Disease only | 0% | 90% |
| Combined, with immunosenescence | 90%; 45% in 65+ year age group | Additional 60% efficacy for breakthrough infections |
| Combined, lower efficacy | 70% | Additional 60% efficacy for breakthrough infections |

#### 1.3 Mathematical model equations

##### 1.3.1 Vaccination group $v_0$ - unvaccinated

$$\frac{dS(t,a,v_0)}{dt} = 2\rho R_2(t,a,v_0) - \beta\frac{S(t,a,v_0)}{N}\sum_{a'} c(a,a')[\sum_{v}(I_{MILD}(t,a',v) + I_{CASE}(t,a',v))] - \kappa(a)S(t,a,v_0)$$

$$\frac{dE_1(t,a,v_0)}{dt} = \beta\frac{S(t,a,v_0)}{N}\sum_{a'} c(a,a')[\sum_{v}(I_{MILD}(t,a',v) + I_{CASE}(t,a',v))] - 2\alpha E_1(t,a,v_0) - \kappa(a)E_1(t,a,v_0)$$

$$\frac{dE_2(t,a,v_0)}{dt} = 2\alpha E_1(t,a,v_0) - 2\alpha E_2(t,a,v_0) - \kappa(a)E_2(t,a,v_0)$$

$$\frac{dI_{MILD}(t,a,v_0)}{dt} = (1-\phi_1(a))(2\alpha E_2(t,a,v_0)) - \gamma_1 I_{MILD}(t,a,v_0)$$

$$\frac{dI_{CASE,0}(t,a,v_0)}{dt} = \phi_1(a)(2\alpha E_2(t,a,v_0)) - 2\gamma_2 I_{CASE,0}(t,a,v_0)$$

$$\frac{dI_{CASE,1}(t,a,v_0)}{dt} = 2\gamma_2 I_{CASE,0}(t,a,v_0) - 2\gamma_2 I_{CASE,1}(t,a,v_0)$$

$$\frac{dI_{HOSPITAL,0}(t,a,v_0,0,0)}{dt} = (1-\delta(H))\mu(a)(1-\phi_2(a))2\gamma_2 I_{CASE,1}(t,a,v_0) - 2\gamma_{3,0} I_{HOSPITAL,0}(t,a,v_0,0,0)$$

$$\frac{dI_{HOSPITAL,1}(t,a,v_0,0,0)}{dt} = 2\gamma_{3,0} I_{HOSPITAL,0}(t,a,v_0,0,0) - 2\gamma_{3,0} I_{HOSPITAL,1}(t,a,v_0,0,0)$$

$$\frac{dI_{HOSPITAL,0}(t,a,v_0,1,0)}{dt} = \delta(H)\mu(a)(1-\phi_2(a))2\gamma_2 I_{CASE,1}(t,a,v_0) - 2\gamma_{3,0} I_{HOSPITAL,0}(t,a,v_0,1,0)$$

$$\frac{dI_{HOSPITAL,1}(t,a,v_0,1,0)}{dt} = 2\gamma_{3,0} I_{HOSPITAL,0}(t,a,v_0,1,0) - 2\gamma_{3,0} I_{HOSPITAL,1}(t,a,v_0,1,0)$$

$$\frac{dI_{HOSPITAL,0}(t,a,v_0,0,1)}{dt} = (1-\delta(H))(1-\mu(a))(1-\phi_2(a))2\gamma_2 I_{CASE,1}(t,a,v_0) - 2\gamma_{3,1} I_{HOSPITAL,0}(t,a,v_0,0,1)$$

$$\frac{dI_{HOSPITAL,1}(t,a,v_0,0,1)}{dt} = 2\gamma_{3,1} I_{HOSPITAL,0}(t,a,v_0,0,1) - 2\gamma_{3,1} I_{HOSPITAL,1}(t,a,v_0,0,1)$$

$$\frac{dI_{HOSPITAL,0}(t,a,v_0,1,1)}{dt} = \delta(H)(1-\mu(a))(1-\phi_2(a))2\gamma_2 I_{CASE,1}(t,a,v_0) - 2\gamma_{3,1} I_{HOSPITAL,0}(t,a,v_0,1,1)$$

$$\frac{dI_{HOSPITAL,1}(t,a,v_0,1,1)}{dt} = 2\gamma_{3,1} I_{HOSPITAL,0}(t,a,v_0,1,1) - 2\gamma_{3,1} I_{HOSPITAL,1}(t,a,v_0,1,1)$$

$$\frac{dI_{ICU,0}(t,a,v_0,0,0)}{dt} = (1-\delta(ICU))\mu(a)\phi_2(a)2\gamma_2 I_{CASE,1}(t,a,v_0) - 2\gamma_{4,0} I_{ICU,0}(t,a,v_0,0,0)$$

$$\frac{dI_{ICU,1}(t,a,v_0,0,0)}{dt} = 2\gamma_{4,0} I_{ICU,0}(t,a,v_0,0,0) - 2\gamma_{4,0} I_{ICU,1}(t,a,v_0,0,0)$$

$$\frac{dI_{ICU,0}(t,a,v_0,1,0)}{dt} = \delta(ICU)\mu(a)\phi_2(a)2\gamma_2 I_{CASE,1}(t,a,v_0) - 2\gamma_{4,0} I_{ICU,0}(t,a,v_0,1,0)$$

$$\frac{dI_{ICU,1}(t,a,v_0,1,0)}{dt} = 2\gamma_{4,0} I_{ICU,0}(t,a,v_0,1,0) - 2\gamma_{4,0} I_{ICU,1}(t,a,v_0,1,0)$$

$$\frac{dI_{ICU,0}(t,a,v_0,0,1)}{dt} = (1-\delta(ICU))(1-\mu(a))\phi_2(a)2\gamma_2 I_{CASE,1}(t,v_0,a) - 2\gamma_{4,1} I_{ICU,0}(t,a,v_0,0,1)$$

$$\frac{dI_{ICU,1}(t,a,v_0,0,1)}{dt} = 2\gamma_{4,1} I_{ICU,0}(t,a,v_0,0,1) - 2\gamma_{4,1} I_{ICU,1}(t,a,v_0,0,1)$$

$$\frac{dI_{ICU,0}(t,a,v_0,1,1)}{dt} = \delta(ICU)(1-\mu(a))\phi_2(a)2\gamma_2 I_{CASE,1}(t,a,v_0) - 2\gamma_{4,1} I_{ICU,0}(t,a,v_0,1,1)$$

$$\frac{dI_{ICU,1}(t,a,v_0,1,1)}{dt} = 2\gamma_{4,1} I_{ICU,0}(t,a,v_0,1,1) - 2\gamma_{4,1} I_{ICU,1}(t,a,v_0,1,1)$$

$$\frac{dI_{REC,0}(t,a,v_0)}{dt} = 2\gamma_{4,1} I_{ICU,1}(t,a,v_0,0,1) + 2\gamma_{4,1} I_{ICU,1}(t,a,v_0,1,1) - 2\gamma_5 I_{REC,0}(t,a,v_0)$$

$$\frac{dI_{REC,1}(t,a,v_0)}{dt} = 2\gamma_5 I_{REC,0}(t,a,v_0) - 2\gamma_5 I_{REC,1}(t,a,v_0)$$

$$\begin{aligned}\frac{dR_1(t,a,v_0)}{dt} = {} & \gamma_1 I_{MILD}(t,a,v_0) + 2\gamma_{3,1} I_{HOSPITAL,1}(t,a,v_0,0,1) + 2\gamma_{3,1} I_{HOSPITAL,1}(t,a,v_0,1,1) + 2\gamma_5 I_{REC,1}(t,a,v_0) \\ & + 2\gamma_{4,1} I_{ICU,1}(t,a,v_0,0,1) + 2\gamma_{4,1} I_{ICU,1}(t,a,v_0,1,1) - 2\rho R_1(t,a,v_0) - \kappa(a)R_1(t,a,v_0)\end{aligned}$$

$$\frac{dR_2(t,a,v_0)}{dt} = 2\rho R_1(t,a,v_0) - 2\rho R_2(t,a,v_0) - \kappa(a)R_2(t,a,v_0)$$

$$\begin{aligned}\frac{dD(t,a,v_0)}{dt} = {} & 2\gamma_{3,0} I_{HOSPITAL,1}(t,a,v_0,0,0) + 2\gamma_{3,0} I_{HOSPITAL,1}(t,a,v_0,1,0) + 2\gamma_{4,0} I_{ICU,1}(t,a,v_0,0,0) \\ & + 2\gamma_{4,0} I_{ICU,1}(t,a,v_0,1,0)\end{aligned}$$

##### 1.3.2 Vaccination group $v_1$ - vaccinated but not yet protected (state 1)

$$\frac{dS(t,a,v_1)}{dt} = \kappa(a)S(t,a,v_0) + 2\rho R_2(t,a,v_1) - \beta\frac{S(t,a,v_1)}{N}\sum_{a'} c(a,a')[\sum_{v}(I_{MILD}(t,a',v) + I_{CASE}(t,a',v))] - 2\omega S(t,a,v_1)$$

$$\frac{dE_1(t,a,v_1)}{dt} = \kappa(a)E_1(t,a,v_0) + \beta\frac{S(t,a,v_1)}{N}\sum_{a'} c(a,a')[\sum_{v}(I_{MILD}(t,a',v) + I_{CASE}(t,a',v))] - 2\alpha E_1(t,a,v_1) - 2\omega E_1(t,a,v_1)$$

$$\frac{dE_2(t,a,v_1)}{dt} = \kappa(a)E_2(t,a,v_0) + 2\alpha E_1(t,a,v_1) - 2\alpha E_2(t,a,v_1) - 2\omega E_2(t,a,v_1)$$

$$\frac{dI_{MILD}(t,a,v_1)}{dt} = (1-\phi_1(a))(2\alpha E_2(t,a,v_1)) - \gamma_1 I_{MILD}(t,a,v_1) - 2\omega I_{MILD}(t,a,v_1)$$

$$\frac{dI_{CASE,0}(t,a,v_1)}{dt} = \phi_1(a)(2\alpha E_2(t,a,v_1)) - 2\gamma_2 I_{CASE,0}(t,a,v_1) - 2\omega I_{CASE,0}(t,a,v_1)$$

$$\frac{dI_{CASE,1}(t,a,v_1)}{dt} = 2\gamma_2 I_{CASE,0}(t,a,v_1) - 2\gamma_2 I_{CASE,1}(t,a,v_1) - 2\omega I_{CASE,1}(t,a,v_1)$$

$$\frac{dI_{HOSPITAL,0}(t,a,v_1,0,0)}{dt} = (1-\delta(H))\mu(a)(1-\phi_2(a))2\gamma_2 I_{CASE,1}(t,a,v_1) - 2\gamma_{3,0} I_{HOSPITAL,0}(t,a,v_1,0,0) - 2\omega I_{HOSPITAL,0}(t,a,v_1,0,0)$$

$$\frac{dI_{HOSPITAL,1}(t,a,v_1,0,0)}{dt} = 2\gamma_{3,0} I_{HOSPITAL,0}(t,a,v_1,0,0) - 2\gamma_{3,0} I_{HOSPITAL,1}(t,a,v_1,0,0) - 2\omega I_{HOSPITAL,1}(t,a,v_1,0,0)$$

$$\frac{dI_{HOSPITAL,0}(t,a,v_1,1,0)}{dt} = \delta(H)\mu(a)(1-\phi_2(a))2\gamma_2 I_{CASE,1}(t,a,v_1) - 2\gamma_{3,0} I_{HOSPITAL,0}(t,a,v_1,1,0) - 2\omega I_{HOSPITAL,0}(t,a,v_1,1,0)$$

$$\frac{dI_{HOSPITAL,1}(t,a,v_1,1,0)}{dt} = 2\gamma_{3,0} I_{HOSPITAL,0}(t,a,v_1,1,0) - 2\gamma_{3,0} I_{HOSPITAL,1}(t,a,v_1,1,0) - 2\omega I_{HOSPITAL,1}(t,a,v_1,1,0)$$

$$\frac{dI_{HOSPITAL,0}(t,a,v_1,0,1)}{dt} = (1-\delta(H))(1-\mu(a))(1-\phi_2(a))2\gamma_2 I_{CASE,1}(t,a,v_1) - 2\gamma_{3,1} I_{HOSPITAL,0}(t,a,v_1,0,1) - 2\omega I_{HOSPITAL,0}(t,a,v_1,0,1)$$

$$\frac{dI_{HOSPITAL,1}(t,a,v_1,0,1)}{dt} = 2\gamma_{3,1} I_{HOSPITAL,0}(t,a,v_1,0,1) - 2\gamma_{3,1} I_{HOSPITAL,1}(t,a,v_1,0,1) - 2\omega I_{HOSPITAL,1}(t,a,v_1,0,1)$$

$$\frac{dI_{HOSPITAL,0}(t,a,v_1,1,1)}{dt} = \delta(H)(1-\mu(a))(1-\phi_2(a))2\gamma_2 I_{CASE,1}(t,a,v_1) - 2\gamma_{3,1} I_{HOSPITAL,0}(t,a,v_1,1,1) - 2\omega I_{HOSPITAL,0}(t,a,v_1,1,1)$$

$$\frac{dI_{HOSPITAL,1}(t,a,v_1,1,1)}{dt} = 2\gamma_{3,1} I_{HOSPITAL,0}(t,a,v_1,1,1) - 2\gamma_{3,1} I_{HOSPITAL,1}(t,a,v_1,1,1) - 2\omega I_{HOSPITAL,1}(t,a,v_1,1,1)$$

$$\frac{dI_{ICU,0}(t,a,v_1,0,0)}{dt} = (1-\delta(ICU))\mu(a)\phi_2(a)2\gamma_2 I_{CASE,1}(t,a,v_1) - 2\gamma_{4,0} I_{ICU,0}(t,a,v_1,0,0) - 2\omega I_{ICU,0}(t,a,v_1,0,0)$$

$$\frac{dI_{ICU,1}(t,a,v_1,0,0)}{dt} = 2\gamma_{4,0} I_{ICU,0}(t,a,v_1,0,0) - 2\gamma_{4,0} I_{ICU,1}(t,a,v_1,0,0) - 2\omega I_{ICU,1}(t,a,v_1,0,0)$$

$$\frac{dI_{ICU,0}(t,a,v_1,1,0)}{dt} = \delta(ICU)\mu(a)\phi_2(a)2\gamma_2 I_{CASE,1}(t,a,v_1) - 2\gamma_{4,0} I_{ICU,0}(t,a,v_1,1,0) - 2\omega I_{ICU,0}(t,a,v_1,1,0)$$

$$\frac{dI_{ICU,1}(t,a,v_1,1,0)}{dt} = 2\gamma_{4,0} I_{ICU,0}(t,a,v_1,1,0) - 2\gamma_{4,0} I_{ICU,1}(t,a,v_1,1,0) - 2\omega I_{ICU,1}(t,a,v_1,1,0)$$

$$\frac{dI_{ICU,0}(t,a,v_1,0,1)}{dt} = (1-\delta(ICU))(1-\mu(a))\phi_2(a)2\gamma_2 I_{CASE,1}(t,v_1,a) - 2\gamma_{4,1} I_{ICU,0}(t,a,v_1,0,1) - 2\omega I_{ICU,0}(t,a,v_1,0,1)$$

$$\frac{dI_{ICU,1}(t,a,v_1,0,1)}{dt} = 2\gamma_{4,1} I_{ICU,0}(t,a,v_1,0,1) - 2\gamma_{4,1} I_{ICU,1}(t,a,v_1,0,1) - 2\omega I_{ICU,1}(t,a,v_1,0,1)$$

$$\frac{dI_{ICU,0}(t,a,v_1,1,1)}{dt} = \delta(ICU)(1-\mu(a))\phi_2(a)2\gamma_2 I_{CASE,1}(t,a,v_1) - 2\gamma_{4,1} I_{ICU,0}(t,a,v_1,1,1) - 2\omega I_{ICU,0}(t,a,v_1,1,1)$$

$$\frac{dI_{ICU,1}(t,a,v_1,1,1)}{dt} = 2\gamma_{4,1} I_{ICU,0}(t,a,v_1,1,1) - 2\gamma_{4,1} I_{ICU,1}(t,a,v_1,1,1) - 2\omega I_{ICU,1}(t,a,v_1,1,1)$$

$$\frac{dI_{REC,0}(t,a,v_1)}{dt} = 2\gamma_{4,1} I_{ICU,1}(t,a,v_1,0,1) + 2\gamma_{4,1} I_{ICU,1}(t,a,v_1,1,1) - 2\gamma_5 I_{REC,0}(t,a,v_1) - 2\omega I_{REC,0}(t,a,v_1)$$

$$\frac{dI_{REC,1}(t,a,v_1)}{dt} = 2\gamma_5 I_{REC,0}(t,a,v_1) - 2\gamma_5 I_{REC,1}(t,a,v_1) - 2\omega I_{REC,1}(t,a,v_1)$$

$$\frac{dR_1(t,a,v_1)}{dt} = \kappa(a)R_1(t,a,v_0) + \gamma_1 I_{MILD}(t,a,v_1) + 2\gamma_{3,1} I_{HOSPITAL,1}(t,a,v_1,0,1) + 2\gamma_{3,1} I_{HOSPITAL,1}(t,a,v_1,1,1) + 2\gamma_5 I_{REC,1}(t,a,v_1) + 2\gamma_{4,1} I_{ICU,1}(t,a,v_1,0,1) + 2\gamma_{4,1} I_{ICU,1}(t,a,v_1,1,1) - 2\rho R_1(t,a,v_1) - 2\omega R(t,a,v_1)$$

$$\frac{dR_2(t,a,v_1)}{dt} = \kappa(a)R_2(t,a,v_0) + 2\rho R_1(t,a,v_1) - 2\rho R_2(t,a,v_1)$$

$$\frac{dD(t,a,v_1)}{dt} = 2\gamma_{3,0} I_{HOSPITAL,1}(t,a,v_1,0,0) + 2\gamma_{3,0} I_{HOSPITAL,1}(t,a,v_1,1,0) + 2\gamma_{4,0} I_{ICU,1}(t,a,v_1,0,0) + 2\gamma_{4,0} I_{ICU,1}(t,a,v_1,1,0)$$

##### 1.3.3 Vaccination group $v_2$ - vaccinated but not yet protected (state 2)

$$\frac{dS(t,a,v_2)}{dt} = 2\rho R_2(t,a,v_2) - \beta\frac{S(t,a,v_2)}{N}\sum_{a'} c(a,a')[\sum_{v}(I_{MILD}(t,a',v) + I_{CASE}(t,a',v))] + 2\omega S(t,a,v_1) - 2\omega S(t,a,v_2)$$

$$\frac{dE_1(t,a,v_2)}{dt} = \beta\frac{S(t,a,v_2)}{N}\sum_{a'} c(a,a')[\sum_{v}(I_{MILD}(t,a',v) + I_{CASE}(t,a',v))] - 2\alpha E_1(t,a,v_2) + 2\omega E_1(t,a,v_1) - 2\omega E_1(t,a,v_2)$$

$$\frac{dE_2(t,a,v_2)}{dt} = 2\alpha E_1(t,a,v_2) - 2\alpha E_2(t,a,v_2) + 2\omega E_2(t,a,v_1) - 2\omega E_2(t,a,v_2)$$

$$\frac{dI_{MILD}(t,a,v_2)}{dt} = (1-\phi_1(a))(2\alpha E_2(t,a,v_2)) - \gamma_1 I_{MILD}(t,a,v_2) + 2\omega I_{MILD}(t,a,v_1) - 2\omega I_{MILD}(t,a,v_2)$$

$$\frac{dI_{CASE,0}(t,a,v_2)}{dt} = \phi_1(a)(2\alpha E_2(t,a,v_2)) - 2\gamma_2 I_{CASE,0}(t,a,v_2) + 2\omega I_{CASE,0}(t,a,v_1) - 2\omega I_{CASE,0}(t,a,v_2)$$

$$\frac{dI_{CASE,1}(t,a,v_2)}{dt} = 2\gamma_2 I_{CASE,0}(t,a,v_2) - 2\gamma_2 I_{CASE,1}(t,a,v_2) + 2\omega I_{CASE,1}(t,a,v_1) - 2\omega I_{CASE,1}(t,a,v_2)$$

$$\frac{dI_{HOSPITAL,0}(t,a,v_2,0{,}0)}{dt} = (1-\delta(H))\mu(a)(1-\phi_2(a))2\gamma_2 I_{CASE,1}(t,a,v_2) - 2\gamma_{3,0} I_{HOSPITAL,0}(t,a,v_2,0{,}0) + 2\omega I_{HOSPITAL,0}(t,a,v_1,0{,}0) - 2\omega I_{HOSPITAL,0}(t,a,v_2,0{,}0)$$

$$\frac{dI_{HOSPITAL,1}(t,a,v_2,0{,}0)}{dt} = 2\gamma_{3,0} I_{HOSPITAL,0}(t,a,v_2,0{,}0) - 2\gamma_{3,0} I_{HOSPITAL,1}(t,a,v_2,0{,}0) + 2\omega I_{HOSPITAL,1}(t,a,v_1,0{,}0) - 2\omega I_{HOSPITAL,1}(t,a,v_2,0{,}0)$$

$$\frac{dI_{HOSPITAL,0}(t,a,v_2,1{,}0)}{dt} = \delta(H)\mu(a)(1-\phi_2(a))2\gamma_2 I_{CASE,1}(t,a,v_2) - 2\gamma_{3,0} I_{HOSPITAL,0}(t,a,v_2,1{,}0) + 2\omega I_{HOSPITAL,0}(t,a,v_1,1{,}0) - 2\omega I_{HOSPITAL,0}(t,a,v_2,1{,}0)$$

$$\frac{dI_{HOSPITAL,1}(t,a,v_2,1{,}0)}{dt} = 2\gamma_{3,0} I_{HOSPITAL,0}(t,a,v_2,1{,}0) - 2\gamma_{3,0} I_{HOSPITAL,1}(t,a,v_2,1{,}0) + 2\omega I_{HOSPITAL,1}(t,a,v_1,1{,}0) - 2\omega I_{HOSPITAL,1}(t,a,v_2,1{,}0)$$

$$\frac{dI_{HOSPITAL,0}(t,a,v_2,0{,}1)}{dt} = (1-\delta(H))(1-\mu(a))(1-\phi_2(a))2\gamma_2 I_{CASE,1}(t,a,v_2) - 2\gamma_{3,1} I_{HOSPITAL,0}(t,a,v_2,0{,}1) + 2\omega I_{HOSPITAL,0}(t,a,v_1,0{,}1) - 2\omega I_{HOSPITAL,0}(t,a,v_2,0{,}1)$$

$$\frac{dI_{HOSPITAL,1}(t,a,v_2,0{,}1)}{dt} = 2\gamma_{3,1} I_{HOSPITAL,0}(t,a,v_2,0{,}1) - 2\gamma_{3,1} I_{HOSPITAL,1}(t,a,v_2,0{,}1) + 2\omega I_{HOSPITAL,1}(t,a,v_1,0{,}1) - 2\omega I_{HOSPITAL,1}(t,a,v_2,0{,}1)$$

$$\frac{dI_{HOSPITAL,0}(t,a,v_2,1{,}1)}{dt} = \delta(H)(1-\mu(a))(1-\phi_2(a))2\gamma_2 I_{CASE,1}(t,a,v_2) - 2\gamma_{3,1} I_{HOSPITAL,0}(t,a,v_2,1{,}1) + 2\omega I_{HOSPITAL,0}(t,a,v_1,1{,}1) - 2\omega I_{HOSPITAL,0}(t,a,v_2,1{,}1)$$

$$\frac{dI_{HOSPITAL,1}(t,a,v_2,1{,}1)}{dt} = 2\gamma_{3,1} I_{HOSPITAL,0}(t,a,v_2,1{,}1) - 2\gamma_{3,1} I_{HOSPITAL,1}(t,a,v_2,1{,}1) + 2\omega I_{HOSPITAL,1}(t,a,v_1,1{,}1) - 2\omega I_{HOSPITAL,1}(t,a,v_2,1{,}1)$$

$$\frac{dI_{ICU,0}(t,a,v_2,0{,}0)}{dt} = (1-\delta(ICU))\mu(a)\phi_2(a)2\gamma_2 I_{CASE,1}(t,a,v_2) - 2\gamma_{4,0} I_{ICU,0}(t,a,v_2,0{,}0) + 2\omega I_{ICU,0}(t,a,v_1,0{,}0) - 2\omega I_{ICU,0}(t,a,v_2,0{,}0)$$

$$\frac{dI_{ICU,1}(t,a,v_2,0{,}0)}{dt} = 2\gamma_{4,0} I_{ICU,0}(t,a,v_2,0{,}0) - 2\gamma_{4,0} I_{ICU,1}(t,a,v_2,0{,}0) + 2\omega I_{ICU,1}(t,a,v_1,0{,}0) - 2\omega I_{ICU,1}(t,a,v_2,0{,}0)$$

$$\frac{dI_{ICU,0}(t,a,v_2,1{,}0)}{dt} = \delta(ICU)\mu(a)\phi_2(a)2\gamma_2 I_{CASE,1}(t,a,v_2) - 2\gamma_{4,0} I_{ICU,0}(t,a,v_2,1{,}0) + 2\omega I_{ICU,0}(t,a,v_1,1{,}0) - 2\omega I_{ICU,0}(t,a,v_2,1{,}0)$$

$$\frac{dI_{ICU,1}(t,a,v_2,1{,}0)}{dt} = 2\gamma_{4,0}I_{ICU,0}(t,a,v_2,1{,}0) - 2\gamma_{4,0}I_{ICU,1}(t,a,v_2,1{,}0) + 2\omega I_{ICU,1}(t,a,v_1,1{,}0) - 2\omega I_{ICU,1}(t,a,v_2,1{,}0)$$

$$\frac{dI_{ICU,0}(t,a,v_2,0{,}1)}{dt} = (1-\delta(ICU))(1-\mu(a))\phi_2(a)2\gamma_2 I_{CASE,1}(t,v_2,a) - 2\gamma_{4,1}I_{ICU,0}(t,a,v_2,0{,}1) + 2\omega I_{ICU,0}(t,a,v_1,0{,}1) - 2\omega I_{ICU,0}(t,a,v_2,0{,}1)$$

$$\frac{dI_{ICU,1}(t,a,v_2,0{,}1)}{dt} = 2\gamma_{4,1}I_{ICU,0}(t,a,v_2,0{,}1) - 2\gamma_{4,1}I_{ICU,1}(t,a,v_2,0{,}1) + 2\omega I_{ICU,1}(t,a,v_1,0{,}1) - 2\omega I_{ICU,1}(t,a,v_2,0{,}1)$$

$$\frac{dI_{ICU,0}(t,a,v_2,1{,}1)}{dt} = \delta(ICU)(1-\mu(a))\phi_2(a)2\gamma_2 I_{CASE,1}(t,a,v_2) - 2\gamma_{4,1}I_{ICU,0}(t,a,v_2,1{,}1) + 2\omega I_{ICU,0}(t,a,v_1,1{,}1) - 2\omega I_{ICU,0}(t,a,v_2,1{,}1)$$

$$\frac{dI_{ICU,1}(t,a,v_2,1{,}1)}{dt} = 2\gamma_{4,1}I_{ICU,0}(t,a,v_2,1{,}1) - 2\gamma_{4,1}I_{ICU,1}(t,a,v_2,1{,}1) + 2\omega I_{ICU,1}(t,a,v_1,1{,}1) - 2\omega I_{ICU,1}(t,a,v_2,1{,}1)$$

$$\frac{dI_{REC,0}(t,a,v_2)}{dt} = 2\gamma_{4,1}I_{ICU,1}(t,a,v_2,0{,}1) + 2\gamma_{4,1}I_{ICU,1}(t,a,v_2,1{,}1) - 2\gamma_5 I_{REC,0}(t,a,v_2) + 2\omega I_{REC,0}(t,a,v_1) - 2\omega I_{REC,0}(t,a,v_2)$$

$$\frac{dI_{REC,1}(t,a,v_2)}{dt} = 2\gamma_5 I_{REC,0}(t,a,v_2) - 2\gamma_5 I_{REC,1}(t,a,v_2) + 2\omega I_{REC,1}(t,a,v_1) - 2\omega I_{REC,1}(t,a,v_2)$$

$$\frac{dR_1(t,a,v_2)}{dt} = \gamma_1 I_{MILD}(t,a,v_2) + 2\gamma_{3,1}I_{HOSPITAL,1}(t,a,v_2,0{,}1) + 2\gamma_{3,1}I_{HOSPITAL,1}(t,a,v_2,1{,}1) + 2\gamma_5 I_{REC,1}(t,a,v_2) + 2\gamma_{4,1}I_{ICU,1}(t,a,v_2,0{,}1) + 2\gamma_{4,1}I_{ICU,1}(t,a,v_2,1{,}1) - 2\rho R_1(t,a,v_2) + 2\omega R_1(t,a,v_1) - 2\omega R_1(t,a,v_2)$$

$$\frac{dR_2(t,a,v_2)}{dt} = 2\rho R_1(t,a,v_2) - 2\rho R_2(t,a,v_2) + 2\omega R_2(t,a,v_1) - 2\omega R_2(t,a,v_2)$$

$$\frac{dD(t,a,v_2)}{dt} = 2\gamma_{3,0}I_{HOSPITAL,1}(t,a,v_2,0{,}0) + 2\gamma_{3,0}I_{HOSPITAL,1}(t,a,v_2,1{,}0) + 2\gamma_{4,0}I_{ICU,1}(t,a,v_2,0{,}0) + 2\gamma_{4,0}I_{ICU,1}(t,a,v_2,1{,}0)$$

##### 1.3.4 Vaccination group $v_3$ - vaccinated and protected (state 1)

$$\frac{dS(t,a,v_3)}{dt} = 2\rho R_2(t,a,v_3) - \upsilon_{inf}(a)\beta\frac{S(t,a,v_3)}{N}\sum_{a'} c(a,a')[\sum_{v}\left(I_{MILD}(t,a',v) + I_{CASE}(t,a',v)\right)] + 2\omega S(t,a,v_2) - 2\psi S(t,a,v_3)$$

$$\frac{dE_1(t,a,v_3)}{dt} = \upsilon_{inf}(a)\beta\frac{S(t,a,v_3)}{N}\sum_{a'} c(a,a')[\sum_{v}(I_{MILD}(t,a',v) + I_{CASE}(t,a',v))] - 2\alpha E_1(t,a,v_2) + 2\omega E_1(t,a,v_2) - 2\psi E_1(t,a,v_3)$$

$$\frac{dE_2(t,a,v_3)}{dt} = 2\alpha E_1(t,a,v_3) - 2\alpha E_2(t,a,v_3) + 2\omega E_2(t,a,v_2) - 2\psi E_2(t,a,v_3)$$

$$\frac{dI_{MILD}(t,a,v_3)}{dt} = (1 - \upsilon_{dis}(a)\phi_1(a))(2\alpha E_2(t,a,v_3)) - \gamma_1 I_{MILD}(t,a,v_3) + 2\omega I_{MILD}(t,a,v_2) - 2\psi I_{MILD}(t,a,v_3)$$

$$\frac{dI_{CASE,0}(t,a,v_3)}{dt} = \upsilon_{dis}(a)\phi_1(a)(2\alpha E_2(t,a,v_3)) - 2\gamma_2 I_{CASE,0}(t,a,v_3) + 2\omega I_{CASE,0}(t,a,v_2) - 2\psi I_{CASE,0}(t,a,v_3)$$

$$\frac{dI_{CASE,1}(t,a,v_3)}{dt} = 2\gamma_2 I_{CASE,0}(t,a,v_3) - 2\gamma_2 I_{CASE,1}(t,a,v_3) + 2\omega I_{CASE,1}(t,a,v_2) - 2\psi I_{CASE,1}(t,a,v_3)$$

$$\frac{dI_{HOSPITAL,0}(t,a,v_3,0,0)}{dt} = (1 - \delta(H))\mu(a)(1 - \phi_2(a))2\gamma_2 I_{CASE,1}(t,a,v_3) - 2\gamma_{3,0} I_{HOSPITAL,0}(t,a,v_3,0,0) + 2\omega I_{HOSPITAL,0}(t,a,v_2,0,0) - 2\psi I_{HOSPITAL,0}(t,a,v_3,0,0)$$

$$\frac{dI_{HOSPITAL,1}(t,a,v_3,0,0)}{dt} = 2\gamma_{3,0} I_{HOSPITAL,0}(t,a,v_3,0,0) - 2\gamma_{3,0} I_{HOSPITAL,1}(t,a,v_3,0,0) + 2\omega I_{HOSPITAL,1}(t,a,v_2,0,0) - 2\psi I_{HOSPITAL,1}(t,a,v_3,0,0)$$

$$\frac{dI_{HOSPITAL,0}(t,a,v_3,1,0)}{dt} = \delta(H)\mu(a)(1 - \phi_2(a))2\gamma_2 I_{CASE,1}(t,a,v_3) - 2\gamma_{3,0} I_{HOSPITAL,0}(t,a,v_3,1,0) + 2\omega I_{HOSPITAL,0}(t,a,v_2,1,0) - 2\psi I_{HOSPITAL,0}(t,a,v_3,1,0)$$

$$\frac{dI_{HOSPITAL,1}(t,a,v_3,1,0)}{dt} = 2\gamma_{3,0} I_{HOSPITAL,0}(t,a,v_3,1,0) - 2\gamma_{3,0} I_{HOSPITAL,1}(t,a,v_3,1,0) + 2\omega I_{HOSPITAL,1}(t,a,v_2,1,0) - 2\psi I_{HOSPITAL,1}(t,a,v_3,1,0)$$

$$\frac{dI_{HOSPITAL,0}(t,a,v_3,0,1)}{dt} = (1 - \delta(H))(1 - \mu(a))(1 - \phi_2(a))2\gamma_2 I_{CASE,1}(t,a,v_3) - 2\gamma_{3,1} I_{HOSPITAL,0}(t,a,v_3,0,1) + 2\omega I_{HOSPITAL,0}(t,a,v_2,0,1) - 2\psi I_{HOSPITAL,0}(t,a,v_3,0,1)$$

$$\frac{dI_{HOSPITAL,1}(t,a,v_3,0,1)}{dt} = 2\gamma_{3,1} I_{HOSPITAL,0}(t,a,v_3,0,1) - 2\gamma_{3,1} I_{HOSPITAL,1}(t,a,v_3,0,1) + 2\omega I_{HOSPITAL,1}(t,a,v_2,0,1) - 2\psi I_{HOSPITAL,1}(t,a,v_3,0,1)$$

$$\frac{dI_{HOSPITAL,0}(t,a,v_3,1,1)}{dt} = \delta(H)(1 - \mu(a))(1 - \phi_2(a))2\gamma_2 I_{CASE,1}(t,a,v_3) - 2\gamma_{3,1} I_{HOSPITAL,0}(t,a,v_3,1,1) + 2\omega I_{HOSPITAL,0}(t,a,v_2,1,1) - 2\psi I_{HOSPITAL,0}(t,a,v_3,1,1)$$

$$\frac{dI_{HOSPITAL,1}(t,a,v_3,1,1)}{dt} = 2\gamma_{3,1} I_{HOSPITAL,0}(t,a,v_3,1,1) - 2\gamma_{3,1} I_{HOSPITAL,1}(t,a,v_3,1,1) + 2\omega I_{HOSPITAL,1}(t,a,v_2,1,1) - 2\psi I_{HOSPITAL,1}(t,a,v_3,1,1)$$

$$\frac{dI_{ICU,0}(t,a,v_3,0,0)}{dt} = (1 - \delta(ICU))\mu(a)\phi_2(a)2\gamma_2 I_{CASE,1}(t,a,v_3) - 2\gamma_{4,0} I_{ICU,0}(t,a,v_3,0,0) + 2\omega I_{ICU,0}(t,a,v_2,0,0) - 2\psi I_{ICU,0}(t,a,v_3,0,0)$$

$$\frac{dI_{ICU,1}(t,a,v_3,0,0)}{dt} = 2\gamma_{4,0} I_{ICU,0}(t,a,v_3,0,0) - 2\gamma_{4,0} I_{ICU,1}(t,a,v_3,0,0) + 2\omega I_{ICU,1}(t,a,v_2,0,0) - 2\psi I_{ICU,1}(t,a,v_3,0,0)$$

$$\frac{dI_{ICU,0}(t,a,v_3,1,0)}{dt} = \delta(ICU)\mu(a)\phi_2(a)2\gamma_2 I_{CASE,1}(t,a,v_3) - 2\gamma_{4,0} I_{ICU,0}(t,a,v_3,1,0) + 2\omega I_{ICU,0}(t,a,v_2,1,0) - 2\psi I_{ICU,0}(t,a,v_3,1,0)$$

$$\frac{dI_{ICU,1}(t,a,v_3,1,0)}{dt} = 2\gamma_{4,0}I_{ICU,0}(t,a,v_3,1,0) - 2\gamma_{4,0}I_{ICU,1}(t,a,v_3,1,0) + 2\omega I_{ICU,1}(t,a,v_2,1,0) - 2\psi I_{ICU,1}(t,a,v_3,1,0)$$

$$\frac{dI_{ICU,0}(t,a,v_3,0,1)}{dt} = (1-\delta(ICU))(1-\mu(a))\phi_2(a)2\gamma_2 I_{CASE,1}(t,v_3,a) - 2\gamma_{4,1}I_{ICU,0}(t,a,v_3,0,1) + 2\omega I_{ICU,0}(t,a,v_2,0,1) - 2\psi I_{ICU,0}(t,a,v_3,0,1)$$

$$\frac{dI_{ICU,1}(t,a,v_3,0,1)}{dt} = 2\gamma_{4,1}I_{ICU,0}(t,a,v_3,0,1) - 2\gamma_{4,1}I_{ICU,1}(t,a,v_3,0,1) + 2\omega I_{ICU,1}(t,a,v_2,0,1) - 2\psi I_{ICU,1}(t,a,v_3,0,1)$$

$$\frac{dI_{ICU,0}(t,a,v_3,1,1)}{dt} = \delta(ICU)(1-\mu(a))\phi_2(a)2\gamma_2 I_{CASE,1}(t,a,v_3) - 2\gamma_{4,1}I_{ICU,0}(t,a,v_3,1,1) + 2\omega I_{ICU,0}(t,a,v_2,1,1) - 2\psi I_{ICU,0}(t,a,v_3,1,1)$$

$$\frac{dI_{ICU,1}(t,a,v_3,1,1)}{dt} = 2\gamma_{4,1}I_{ICU,0}(t,a,v_3,1,1) - 2\gamma_{4,1}I_{ICU,1}(t,a,v_3,1,1) + 2\omega I_{ICU,1}(t,a,v_2,1,1) - 2\psi I_{ICU,1}(t,a,v_3,1,1)$$

$$\frac{dI_{REC,0}(t,a,v_3)}{dt} = 2\gamma_{4,1}I_{ICU,1}(t,a,v_3,0,1) + 2\gamma_{4,1}I_{ICU,1}(t,a,v_3,1,1) - 2\gamma_5 I_{REC,0}(t,a,v_3) + 2\omega I_{REC,0}(t,a,v_2) - 2\psi I_{REC,0}(t,a,v_3)$$

$$\frac{dI_{REC,1}(t,a,v_3)}{dt} = 2\gamma_5 I_{REC,0}(t,a,v_3) - 2\gamma_5 I_{REC,1}(t,a,v_3) + 2\omega I_{REC,1}(t,a,v_2) - 2\psi I_{REC,1}(t,a,v_3)$$

$$\frac{dR_1(t,a,v_3)}{dt} = \gamma_1 I_{MILD}(t,a,v_3) + 2\gamma_{3,1}I_{HOSPITAL,1}(t,a,v_3,0,1) + 2\gamma_{3,1}I_{HOSPITAL,1}(t,a,v_3,1,1) + 2\gamma_5 I_{REC,1}(t,a,v_3) + 2\gamma_{4,1}I_{ICU,1}(t,a,v_3,0,1) + 2\gamma_{4,1}I_{ICU,1}(t,a,v_3,1,1) - 2\rho R_1(t,a,v_3) + 2\omega R_1(t,a,v_2) - 2\psi R_1(t,a,v_3)$$

$$\frac{dR_2(t,a,v_3)}{dt} = 2\rho R_1(t,a,v_3) - 2\rho R_2(t,a,v_3) + 2\omega R_2(t,a,v_2) - 2\psi R_2(t,a,v_3)$$

$$\frac{dD(t,a,v_3)}{dt} = 2\gamma_{3,0}I_{HOSPITAL,1}(t,a,v_3,0,0) + 2\gamma_{3,0}I_{HOSPITAL,1}(t,a,v_3,1,0) + 2\gamma_{4,0}I_{ICU,1}(t,a,v_3,0,0) + 2\gamma_{4,0}I_{ICU,1}(t,a,v_3,1,0)$$

##### 1.3.5 Vaccination group v4 - vaccinated and protected (state 2)

$$\frac{dS(t,a,v_4)}{dt} = 2\rho R_2(t,a,v_4) - v_{inf}(a)\beta\frac{S(t,a,v_4)}{N}\sum_{a'} c(a,a')[\sum_{v}\left(I_{MILD}(t,a',v) + I_{CASE}(t,a',v)\right)] + 2\psi S(t,a,v_3) - 2\psi S(t,a,v_4)$$

$$\frac{dE_1(t,a,v_4)}{dt} = v_{inf}(a)\beta\frac{S(t,a,v_4)}{N}\sum_{a'} c(a,a')[\sum_{v}(I_{MILD}(t,a',v) + I_{CASE}(t,a',v))] - 2\alpha E_1(t,a,v_4) + 2\psi E_1(t,a,v_3) - 2\psi E_1(t,a,v_4)$$

$$\frac{dE_2(t,a,v_4)}{dt} = 2\alpha E_1(t,a,v_4) - 2\alpha E_2(t,a,v_4) + 2\psi E_2(t,a,v_3) - 2\psi E_2(t,a,v_4)$$

$$\frac{dI_{MILD}(t,a,v_4)}{dt} = (1 - v_{dis}(a)\phi_1(a))(2\alpha E_2(t,a,v_4)) - \gamma_1 I_{MILD}(t,a,v_4) + 2\psi I_{MILD}(t,a,v_3) - 2\psi I_{MILD}(t,a,v_4)$$

$$\frac{dI_{CASE,0}(t,a,v_4)}{dt} = v_{dis}(a)\phi_1(a)(2\alpha E_2(t,a,v_4)) - 2\gamma_2 I_{CASE,0}(t,a,v_4) + 2\psi I_{CASE,0}(t,a,v_3) - 2\psi I_{CASE,0}(t,a,v_4)$$

$$\frac{dI_{CASE,1}(t,a,v_4)}{dt} = 2\gamma_2 I_{CASE,0}(t,a,v_4) - 2\gamma_2 I_{CASE,1}(t,a,v_4) + 2\psi I_{CASE,1}(t,a,v_3) - 2\psi I_{CASE,1}(t,a,v_4)$$

$$\frac{dI_{HOSPITAL,0}(t,a,v_4,0,0)}{dt} = (1-\delta(H))\mu(a)(1-\phi_2(a))2\gamma_2 I_{CASE,1}(t,a,v_4) - 2\gamma_{3,0} I_{HOSPITAL,0}(t,a,v_4,0,0) + 2\psi I_{HOSPITAL,0}(t,a,v_3,0,0) - 2\psi I_{HOSPITAL,0}(t,a,v_4,0,0)$$

$$\frac{dI_{HOSPITAL,1}(t,a,v_4,0,0)}{dt} = 2\gamma_{3,0} I_{HOSPITAL,0}(t,a,v_4,0,0) - 2\gamma_{3,0} I_{HOSPITAL,1}(t,a,v_4,0,0) + 2\psi I_{HOSPITAL,1}(t,a,v_3,0,0) - 2\psi I_{HOSPITAL,1}(t,a,v_4,0,0)$$

$$\frac{dI_{HOSPITAL,0}(t,a,v_4,1,0)}{dt} = \delta(H)\mu(a)(1-\phi_2(a))2\gamma_2 I_{CASE,1}(t,a,v_4) - 2\gamma_{3,0} I_{HOSPITAL,0}(t,a,v_4,1,0) + 2\psi I_{HOSPITAL,0}(t,a,v_3,1,0) - 2\psi I_{HOSPITAL,0}(t,a,v_3,1,0)$$

$$\frac{dI_{HOSPITAL,1}(t,a,v_4,1,0)}{dt} = 2\gamma_{3,0} I_{HOSPITAL,0}(t,a,v_4,1,0) - 2\gamma_{3,0} I_{HOSPITAL,1}(t,a,v_4,1,0) + 2\psi I_{HOSPITAL,1}(t,a,v_3,1,0) - 2\psi I_{HOSPITAL,1}(t,a,v_4,1,0)$$

$$\frac{dI_{HOSPITAL,0}(t,a,v_4,0,1)}{dt} = (1-\delta(H))(1-\mu(a))(1-\phi_2(a))2\gamma_2 I_{CASE,1}(t,a,v_4) - 2\gamma_{3,1} I_{HOSPITAL,0}(t,a,v_4,0,1) + 2\psi I_{HOSPITAL,0}(t,a,v_3,0,1) - 2\psi I_{HOSPITAL,0}(t,a,v_4,0,1)$$

$$\frac{dI_{HOSPITAL,1}(t,a,v_4,0,1)}{dt} = 2\gamma_{3,1} I_{HOSPITAL,0}(t,a,v_4,0,1) - 2\gamma_{3,1} I_{HOSPITAL,1}(t,a,v_4,0,1) + 2\psi I_{HOSPITAL,1}(t,a,v_3,0,1) - 2\psi I_{HOSPITAL,1}(t,a,v_4,0,1)$$

$$\frac{dI_{HOSPITAL,0}(t,a,v_4,1,1)}{dt} = \delta(H)(1-\mu(a))(1-\phi_2(a))2\gamma_2 I_{CASE,1}(t,a,v_4) - 2\gamma_{3,1} I_{HOSPITAL,0}(t,a,v_4,1,1) + 2\psi I_{HOSPITAL,0}(t,a,v_3,1,1) - 2\psi I_{HOSPITAL,0}(t,a,v_4,1,1)$$

$$\frac{dI_{HOSPITAL,1}(t,a,v_4,1,1)}{dt} = 2\gamma_{3,1} I_{HOSPITAL,0}(t,a,v_4,1,1) - 2\gamma_{3,1} I_{HOSPITAL,1}(t,a,v_4,1,1) + 2\psi I_{HOSPITAL,1}(t,a,v_3,1,1) - 2\psi I_{HOSPITAL,1}(t,a,v_4,1,1)$$

$$\frac{dI_{ICU,0}(t,a,v_4,0,0)}{dt} = (1-\delta(ICU))\mu(a)\phi_2(a)2\gamma_2 I_{CASE,1}(t,a,v_4) - 2\gamma_{4,0} I_{ICU,0}(t,a,v_4,0,0) + 2\psi I_{ICU,0}(t,a,v_3,0,0) - 2\psi I_{ICU,0}(t,a,v_4,0,0)$$

$$\frac{dI_{ICU,1}(t,a,v_4,0,0)}{dt} = 2\gamma_{4,0} I_{ICU,0}(t,a,v_4,0,0) - 2\gamma_{4,0} I_{ICU,1}(t,a,v_4,0,0) + 2\psi I_{ICU,1}(t,a,v_3,0,0) - 2\psi I_{ICU,1}(t,a,v_4,0,0)$$

$$\frac{dI_{ICU,0}(t,a,v_4,1{,}0)}{dt} = \delta(ICU)\mu(a)\phi_2(a)2\gamma_2 I_{CASE,1}(t,a,v_4) - 2\gamma_{4,0}I_{ICU,0}(t,a,v_4,1{,}0) + 2\psi I_{ICU,0}(t,a,v_3,1{,}0) - 2\psi I_{ICU,0}(t,a,v_4,1{,}0)$$

$$\frac{dI_{ICU,1}(t,a,v_4,1{,}0)}{dt} = 2\gamma_{4,0}I_{ICU,0}(t,a,v_4,1{,}0) - 2\gamma_{4,0}I_{ICU,1}(t,a,v_4,1{,}0) + 2\psi I_{ICU,1}(t,a,v_3,1{,}0) - 2\psi I_{ICU,1}(t,a,v_4,1{,}0)$$

$$\frac{dI_{ICU,0}(t,a,v_4,0{,}1)}{dt} = (1-\delta(ICU))(1-\mu(a))\phi_2(a)2\gamma_2 I_{CASE,1}(t,v_4,a) - 2\gamma_{4,1}I_{ICU,0}(t,a,v_4,0{,}1) + 2\psi I_{ICU,0}(t,a,v_3,0{,}1) - 2\psi I_{ICU,0}(t,a,v_4,0{,}1)$$

$$\frac{dI_{ICU,1}(t,a,v_4,0{,}1)}{dt} = 2\gamma_{4,1}I_{ICU,0}(t,a,v_4,0{,}1) - 2\gamma_{4,1}I_{ICU,1}(t,a,v_4,0{,}1) + 2\psi I_{ICU,1}(t,a,v_3,0{,}1) - 2\psi I_{ICU,1}(t,a,v_4,0{,}1)$$

$$\frac{dI_{ICU,0}(t,a,v_4,1{,}1)}{dt} = \delta(ICU)(1-\mu(a))\phi_2(a)2\gamma_2 I_{CASE,1}(t,a,v_4) - 2\gamma_{4,1}I_{ICU,0}(t,a,v_4,1{,}1) + 2\psi I_{ICU,0}(t,a,v_3,1{,}1) - 2\psi I_{ICU,0}(t,a,v_4,1{,}1)$$

$$\frac{dI_{ICU,1}(t,a,v_4,1{,}1)}{dt} = 2\gamma_{4,1}I_{ICU,0}(t,a,v_4,1{,}1) - 2\gamma_{4,1}I_{ICU,1}(t,a,v_4,1{,}1) + 2\psi I_{ICU,1}(t,a,v_3,1{,}1) - 2\psi I_{ICU,1}(t,a,v_4,1{,}1)$$

$$\frac{dI_{REC,0}(t,a,v_4)}{dt} = 2\gamma_{4,1}I_{ICU,1}(t,a,v_4,0{,}1) + 2\gamma_{4,1}I_{ICU,1}(t,a,v_4,1{,}1) - 2\gamma_5 I_{REC,0}(t,a,v_4) + 2\psi I_{REC,0}(t,a,v_3) - 2\psi I_{REC,0}(t,a,v_4)$$

$$\frac{dI_{REC,1}(t,a,v_4)}{dt} = 2\gamma_5 I_{REC,0}(t,a,v_4) - 2\gamma_5 I_{REC,1}(t,a,v_4) + 2\psi I_{REC,1}(t,a,v_3) - 2\psi I_{REC,1}(t,a,v_4)$$

$$\frac{dR_1(t,a,v_4)}{dt} = \gamma_1 I_{MILD}(t,a,v_4) + 2\gamma_{3,1}I_{HOSPITAL,1}(t,a,v_4,0{,}1) + 2\gamma_{3,1}I_{HOSPITAL,1}(t,a,v_4,1{,}1) + 2\gamma_5 I_{REC,1}(t,a,v_4) + 2\gamma_{4,1}I_{ICU,1}(t,a,v_4,0{,}1) + 2\gamma_{4,1}I_{ICU,1}(t,a,v_4,1{,}1) - 2\rho R_1(t,a,v_4) + 2\psi R_1(t,a,v_3) - 2\psi R_1(t,a,v_4)$$

$$\frac{dR_2(t,a,v_4)}{dt} = 2\rho R_1(t,a,v_4) - 2\rho R_2(t,a,v_4) + 2\psi R_2(t,a,v_3) - 2\psi R_2(t,a,v_4)$$

$$\frac{dD(t,a,v_4)}{dt} = 2\gamma_{3,0}I_{HOSPITAL,1}(t,a,v_4,0{,}0) + 2\gamma_{3,0}I_{HOSPITAL,1}(t,a,v_4,1{,}0) + 2\gamma_{4,0}I_{ICU,1}(t,a,v_4,0{,}0) + 2\gamma_{4,0}I_{ICU,1}(t,a,v_4,1{,}0)$$

##### 1.3.6 Vaccination group v5 - previously vaccinated but no longer protected

$$\frac{dS(t,a,v_5)}{dt} = 2\rho R_2(t,a,v_5) - \beta\frac{S(t,a,v_5)}{N}\sum_{a'} c(a,a')[\sum_{v}(I_{MILD}(t,a',v) + I_{CASE}(t,a',v))] + 2\psi S(t,a,v_4)$$

$$\frac{dE_1(t,a,v_5)}{dt} = \beta\frac{S(t,a,v_5)}{N}\sum_{a'} c(a,a')[\sum_{v}(I_{MILD}(t,a',v) + I_{CASE}(t,a',v))] - 2\alpha E_1(t,a,v_5) + 2\psi E_1(t,a,v_4)$$

$$\frac{dE_2(t,a,v_5)}{dt} = 2\alpha E_1(t,a,v_5) - 2\alpha E_2(t,a,v_5) + 2\psi E_2(t,a,v_4)$$

$$\frac{dI_{MILD}(t,a,v_5)}{dt} = (1-\phi_1(a))(2\alpha E_2(t,a,v_5)) - \gamma_1 I_{MILD}(t,a,v_5) + 2\psi I_{MILD}(t,a,v_4)$$

$$\frac{dI_{CASE,0}(t,a,v_5)}{dt} = \phi_1(a)(2\alpha E_2(t,a,v_5)) - 2\gamma_2 I_{CASE,0}(t,a,v_5) + 2\psi I_{CASE,0}(t,a,v_4)$$

$$\frac{dI_{CASE,1}(t,a,v_5)}{dt} = 2\gamma_2 I_{CASE,0}(t,a,v_5) - 2\gamma_2 I_{CASE,1}(t,a,v_5) + 2\psi I_{CASE,1}(t,a,v_4)$$

$$\frac{dI_{HOSPITAL,0}(t,a,v_5,0,0)}{dt} = (1-\delta(H))\mu(a)(1-\phi_2(a))2\gamma_2 I_{CASE,1}(t,a,v_5) - 2\gamma_{3,0} I_{HOSPITAL,0}(t,a,v_5,0,0) + 2\psi I_{HOSPITAL,0}(t,a,v_4,0,0)$$

$$\frac{dI_{HOSPITAL,1}(t,a,v_5,0,0)}{dt} = 2\gamma_{3,0} I_{HOSPITAL,0}(t,a,v_5,0,0) - 2\gamma_{3,0} I_{HOSPITAL,1}(t,a,v_5,0,0) + 2\psi I_{HOSPITAL,1}(t,a,v_4,0,0)$$

$$\frac{dI_{HOSPITAL,0}(t,a,v_5,1,0)}{dt} = \delta(H)\mu(a)(1-\phi_2(a))2\gamma_2 I_{CASE,1}(t,a,v_5) - 2\gamma_{3,0} I_{HOSPITAL,0}(t,a,v_5,1,0) + 2\psi I_{HOSPITAL,0}(t,a,v_3,1,0)$$

$$\frac{dI_{HOSPITAL,1}(t,a,v_5,1,0)}{dt} = 2\gamma_{3,0} I_{HOSPITAL,0}(t,a,v_5,1,0) - 2\gamma_{3,0} I_{HOSPITAL,1}(t,a,v_5,1,0) + 2\psi I_{HOSPITAL,1}(t,a,v_4,1,0)$$

$$\frac{dI_{HOSPITAL,0}(t,a,v_5,0,1)}{dt} = (1-\delta(H))(1-\mu(a))(1-\phi_2(a))2\gamma_2 I_{CASE,1}(t,a,v_5) - 2\gamma_{3,1} I_{HOSPITAL,0}(t,a,v_5,0,1) + 2\psi I_{HOSPITAL,0}(t,a,v_4,0,1)$$

$$\frac{dI_{HOSPITAL,1}(t,a,v_5,0,1)}{dt} = 2\gamma_{3,1} I_{HOSPITAL,0}(t,a,v_5,0,1) - 2\gamma_{3,1} I_{HOSPITAL,1}(t,a,v_5,0,1) + 2\psi I_{HOSPITAL,1}(t,a,v_4,0,1)$$

$$\frac{dI_{HOSPITAL,0}(t,a,v_5,1,1)}{dt} = \delta(H)(1-\mu(a))(1-\phi_2(a))2\gamma_2 I_{CASE,1}(t,a,v_5) - 2\gamma_{3,1} I_{HOSPITAL,0}(t,a,v_5,1,1) + 2\psi I_{HOSPITAL,0}(t,a,v_4,1,1)$$

$$\frac{dI_{HOSPITAL,1}(t,a,v_5,1,1)}{dt} = 2\gamma_{3,1} I_{HOSPITAL,0}(t,a,v_5,1,1) - 2\gamma_{3,1} I_{HOSPITAL,1}(t,a,v_5,1,1) + 2\psi I_{HOSPITAL,1}(t,a,v_4,1,1)$$

$$\frac{dI_{ICU,0}(t,a,v_5,0,0)}{dt} = (1-\delta(ICU))\mu(a)\phi_2(a)2\gamma_2 I_{CASE,1}(t,a,v_5) - 2\gamma_{4,0} I_{ICU,0}(t,a,v_5,0,0) + 2\psi I_{ICU,0}(t,a,v_4,0,0)$$

$$\frac{dI_{ICU,1}(t,a,v_5,0,0)}{dt} = 2\gamma_{4,0} I_{ICU,0}(t,a,v_5,0,0) - 2\gamma_{4,0} I_{ICU,1}(t,a,v_5,0,0) + 2\psi I_{ICU,1}(t,a,v_4,0,0)$$

$$\frac{dI_{ICU,0}(t,a,v_5,1,0)}{dt} = \delta(ICU)\mu(a)\phi_2(a)2\gamma_2 I_{CASE,1}(t,a,v_5) - 2\gamma_{4,0} I_{ICU,0}(t,a,v_5,1,0) + 2\psi I_{ICU,0}(t,a,v_4,1,0)$$

$$\frac{dI_{ICU,1}(t,a,v_5,1,0)}{dt} = 2\gamma_{4,0} I_{ICU,0}(t,a,v_5,1,0) - 2\gamma_{4,0} I_{ICU,1}(t,a,v_5,1,0) + 2\psi I_{ICU,1}(t,a,v_4,1,0)$$

$$\frac{dI_{ICU,0}(t,a,v_5,0,1)}{dt} = (1-\delta(ICU))(1-\mu(a))\phi_2(a)2\gamma_2 I_{CASE,1}(t,v_5,a) - 2\gamma_{4,1} I_{ICU,0}(t,a,v_5,0,1) + 2\psi I_{ICU,0}(t,a,v_4,0,1)$$

$$\frac{dI_{ICU,1}(t,a,v_5,0,1)}{dt} = 2\gamma_{4,1} I_{ICU,0}(t,a,v_5,0,1) - 2\gamma_{4,1} I_{ICU,1}(t,a,v_5,0,1) + 2\psi I_{ICU,1}(t,a,v_4,0,1)$$

$$\frac{dI_{ICU,0}(t,a,v_5,1,1)}{dt} = \delta(ICU)(1-\mu(a))\phi_2(a)2\gamma_2 I_{CASE,1}(t,a,v_5) - 2\gamma_{4,1} I_{ICU,0}(t,a,v_5,1,1) + 2\psi I_{ICU,0}(t,a,v_4,1,1)$$

$$\frac{dI_{ICU,1}(t,a,v_5,1,1)}{dt} = 2\gamma_{4,1} I_{ICU,0}(t,a,v_5,1,1) - 2\gamma_{4,1} I_{ICU,1}(t,a,v_5,1,1) + 2\psi I_{ICU,1}(t,a,v_4,1,1)$$

$$\frac{dI_{REC,0}(t,a,v_5)}{dt} = 2\gamma_{4,1} I_{ICU,1}(t,a,v_5,0,1) + 2\gamma_{4,1} I_{ICU,1}(t,a,v_5,1,1) - 2\gamma_5 I_{REC,0}(t,a,v_5) + 2\psi I_{REC,0}(t,a,v_4)$$

$$\frac{dI_{REC,1}(t,a,v_5)}{dt} = 2\gamma_5 I_{REC,0}(t,a,v_5) - 2\gamma_5 I_{REC,1}(t,a,v_5) + 2\psi I_{REC,1}(t,a,v_4)$$

$$\frac{dR_1(t,a,v_5)}{dt} = \gamma_1 I_{MILD}(t,a,v_5) + 2\gamma_{3,1} I_{HOSPITAL,1}(t,a,v_5,0,1) + 2\gamma_{3,1} I_{HOSPITAL,1}(t,a,v_5,1,1) + 2\gamma_5 I_{REC,1}(t,a,v_5) + 2\gamma_{4,1} I_{ICU,1}(t,a,v_5,0,1) + 2\gamma_{4,1} I_{ICU,1}(t,a,v_5,1,1) - 2\rho R_1(t,a,v_5) + 2\psi R_1(t,a,v_4)$$

$$\frac{dR_2(t,a,v_5)}{dt} = 2\rho R_1(t,a,v_5) - 2\rho R_2(t,a,v_5) + 2\psi R_2(t,a,v_4)$$

$$\frac{dD(t,a,v_5)}{dt} = 2\gamma_{3,0} I_{HOSPITAL,1}(t,a,v_5,0,0) + 2\gamma_{3,0} I_{HOSPITAL,1}(t,a,v_5,1,0) + 2\gamma_{4,0} I_{ICU,1}(t,a,v_5,0,0) + 2\gamma_{4,0} I_{ICU,1}(t,a,v_5,1,0)$$

#### 1.4 Model validation

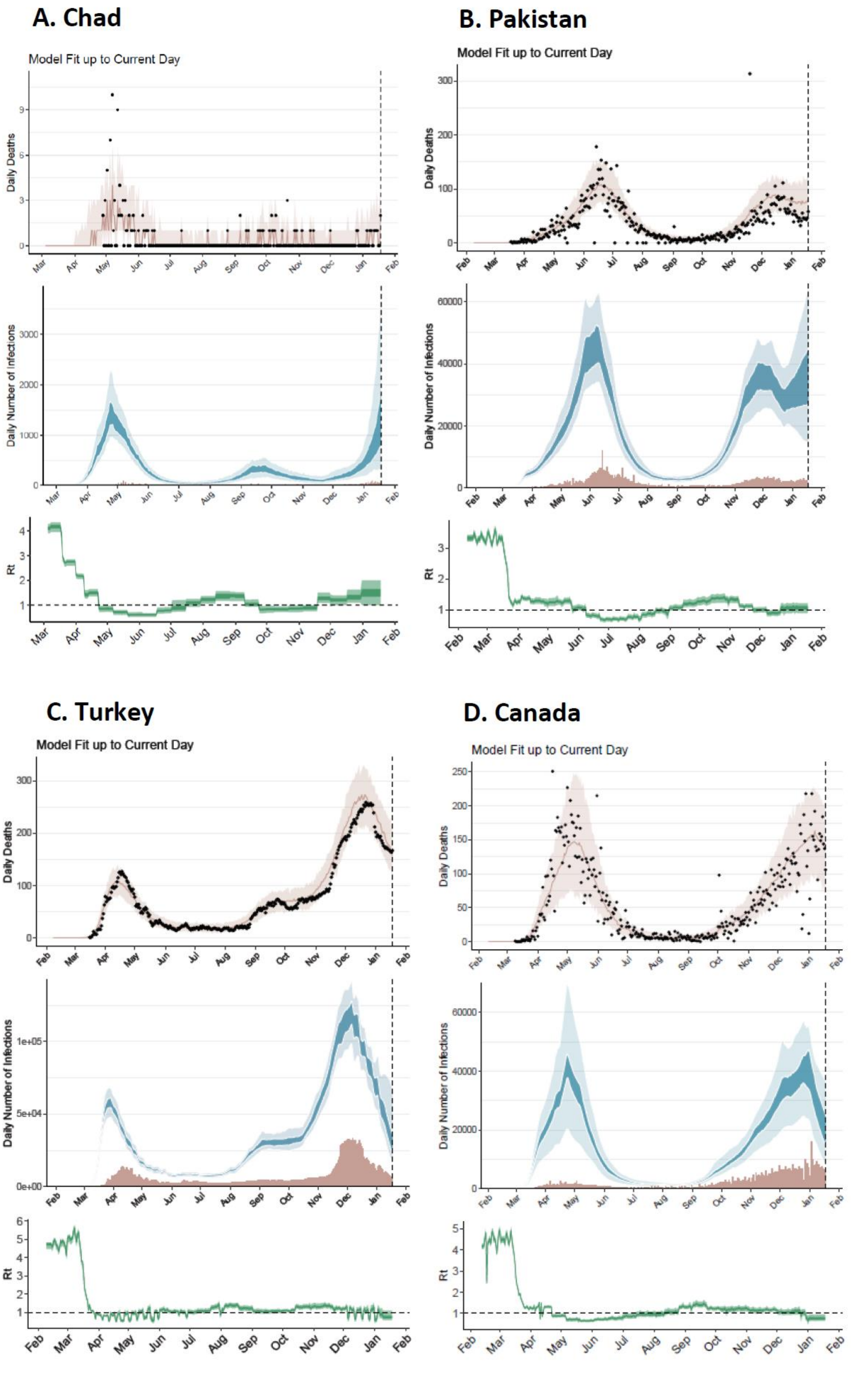


***Figure S2: Country-specific calibration of the SARS-CoV-2 transmission model (as at 18 January 2021)[37].*** *Four example countries are shown: Chad (low-income); Pakistan (lower-middle-income); Turkey (upper-middle-income); and Canada (high-income). Each upper panel shows the modelled daily deaths calibrated to the daily deaths as reported by the COVID-19 Data Repository by the Center for Systems Science and Engineering (CSSE) at Johns Hopkins University[38]. For countries not included in that repository, data are obtained from Worldometer[39]. Each middle panel shows the daily number of infections estimated by fitting to the current total of deaths, with reported cases shown in red and the model-estimated infections shown in blue (dark blue 50% interquartile range, light blue 95% quantile). The dashed vertical lines show the current day. Each lower panel shows the estimated reproduction number over time,* $R_t$.

#### 1.5 Timing of transmission and vaccination

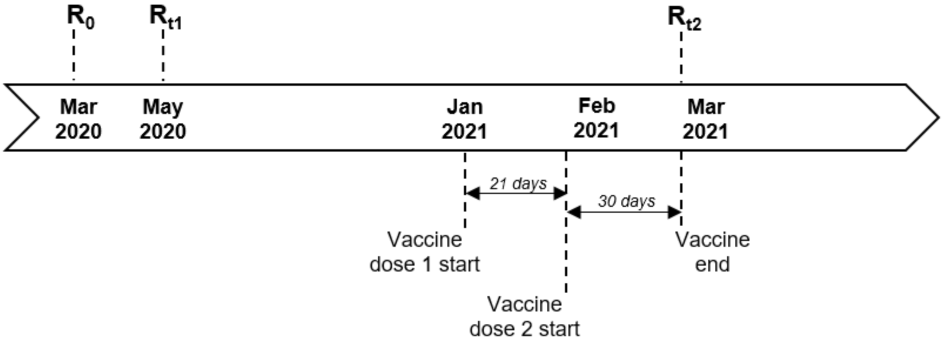


***Figure S3: Schematic illustration of the timing of changes in levels of transmission and the introduction of vaccination.***

#### 1.6 Additional methods for vaccine age-targeting

For the global optimisation of vaccine allocation by income setting and age target, we run the simulation for the vaccine distributed to combinations of 5-year age groups from 0–4 years up to 75–79 years and 80+ years. Rather than simulating impact for every possible combination of age groups targeted, we construct the parameter space such that the vaccine could be targeted to up to two distinct contiguous groups, or rather up two non-overlapping age groups that are each comprised of any number of consecutive 5-year groups. The age group combinations are depicted in Figure S4.

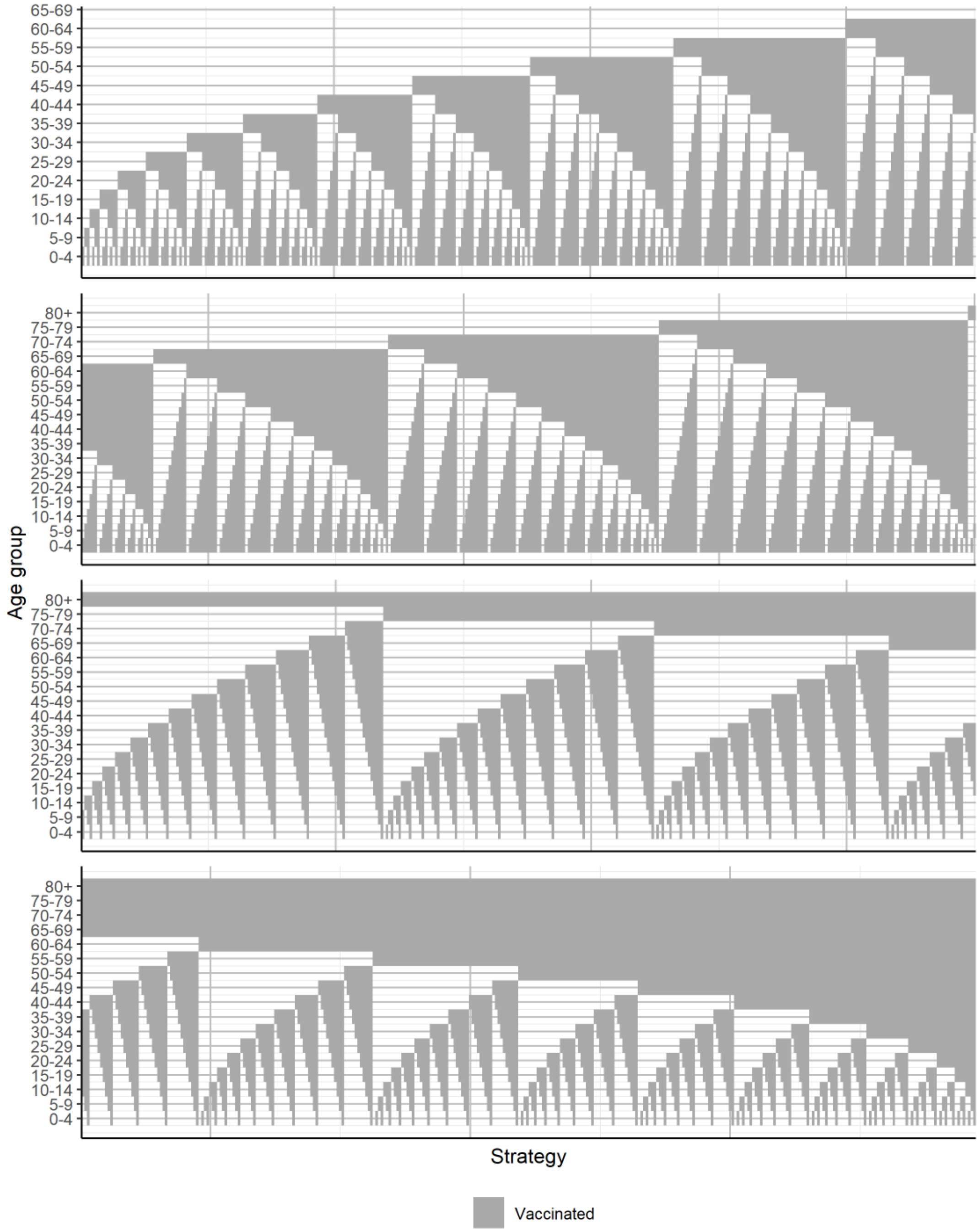


***Figure S4: Illustration of 5-year age group combinations.*** *Each column represents a unique vaccine age-targeting strategy indicating which age groups (rows) are vaccinated (grey shaded regions) or unvaccinated (unshaded regions) for that strategy.*

### 2 Extended results

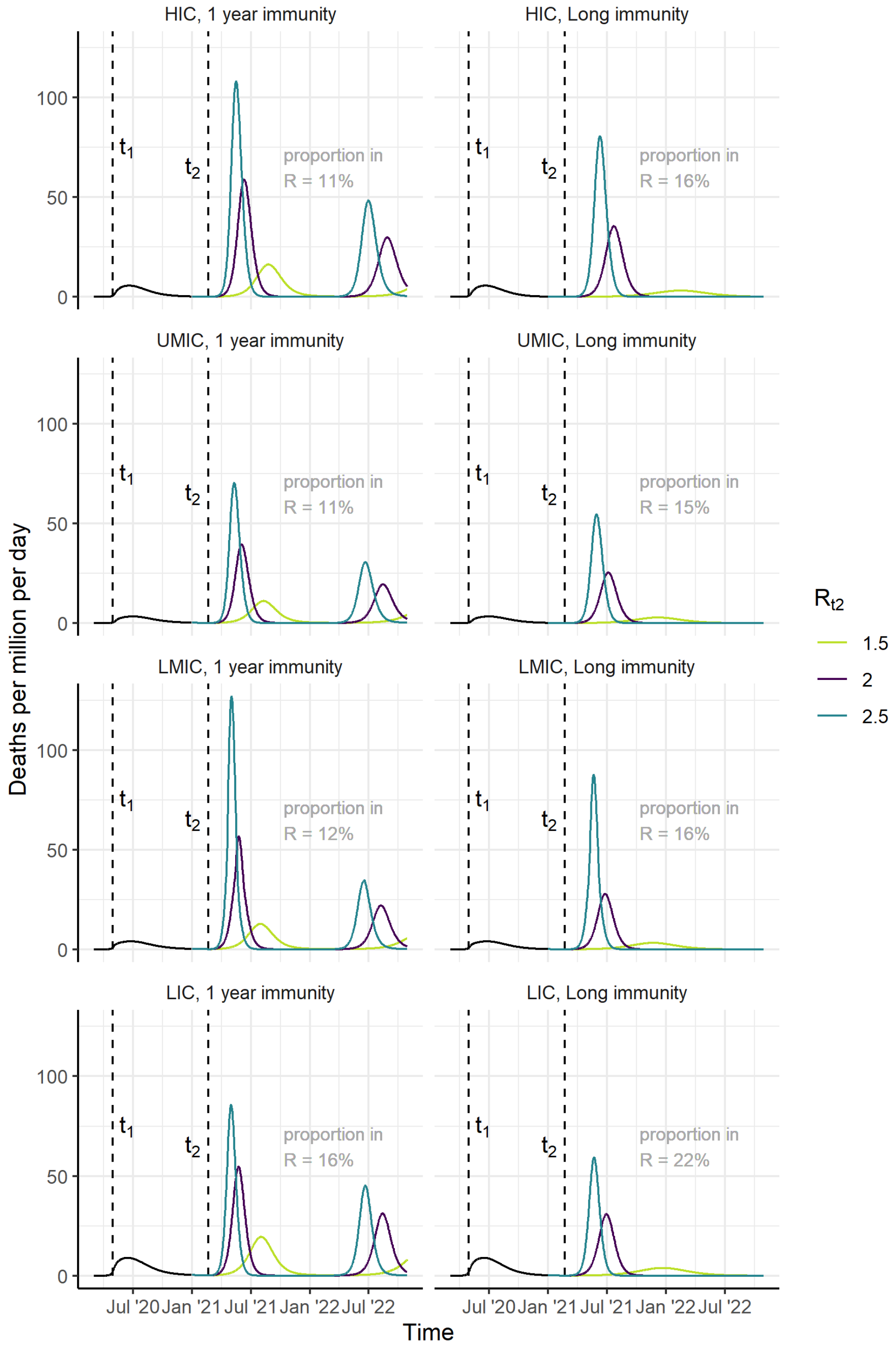


***Figure S5: Scenarios for the course of the epidemic from 2020–2022, for high-income, upper-middle-income, lower-middle-income and low-income country settings (HIC, UMIC, LMIC and LIC respectively).*** *(A, C, E, G) assuming an average duration of naturally acquired immunity of one year; (B, D, F, H) assuming long-term immunity. We assume $R_0$=2.5 up to time $t_1$ (May 2020) and that $R_{t1}$ drops to 1.1 between time $t_1$ and $t_2$ (end-February 2021). From time $t_2$ onwards, we consider three counterfactual scenarios, $R_{t2}$=1.5, 2.0 and 2.5, as shown in light green, purple and turquoise respectively. The grey annotated text indicates the proportion of the population in the recovered class at the time of vaccine introduction (dose 2).*

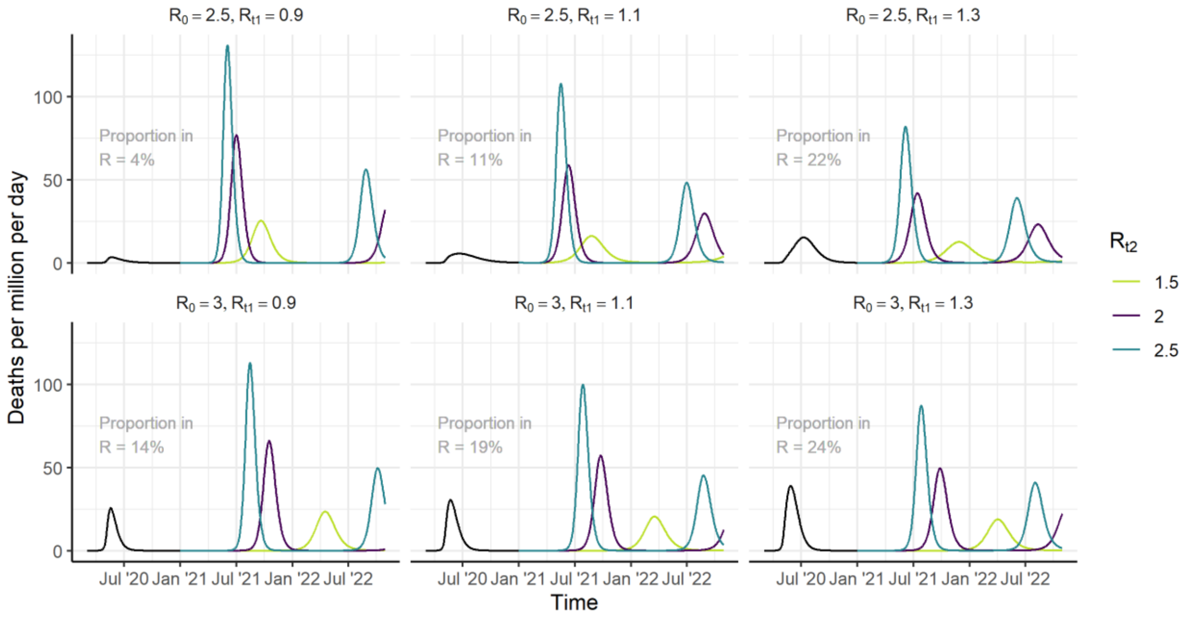


***Figure S6: Scenarios for the course of the epidemic from 2020–2022 (counterfactual scenarios).*** *Epidemic trajectories are shown for a high-income country setting, in the absence of a vaccine, for a range of values of* $R_0$ *(rows),* $R_{t1}$ *(columns), and* $R_{t2}$ *(coloured lines). The grey annotated text indicates the proportion of the population in the recovered class at the time of vaccine introduction (dose 2). Immunity following infection is assumed to persist for one year.*

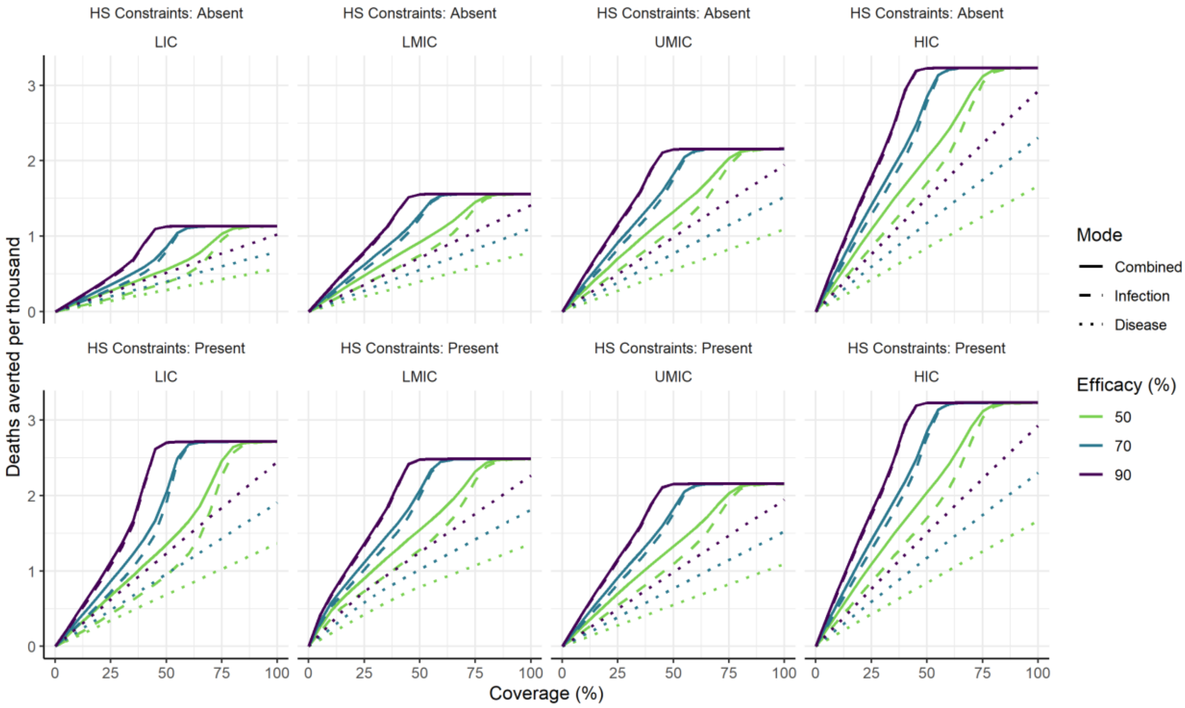


***Figure S7: Vaccine efficacy and herd immunity by income setting.*** *Projected total deaths averted per thousand population in 2021 under the default vaccine scenarios shown in Table 1, for the four income settings (columns), and with health system constraints either absent or present (rows). The colours show different vaccine efficacy assumptions (from 50% to 90%). Solid lines represent impact for a vaccine that is efficacious against infection, with additional efficacy against severe disease. Dashed lines represent a vaccine that is efficacious against infection only, and dotted lines represent a vaccine that prevents severe disease (and hence death) but does not reduce infection or onwards transmission (Table S3).*

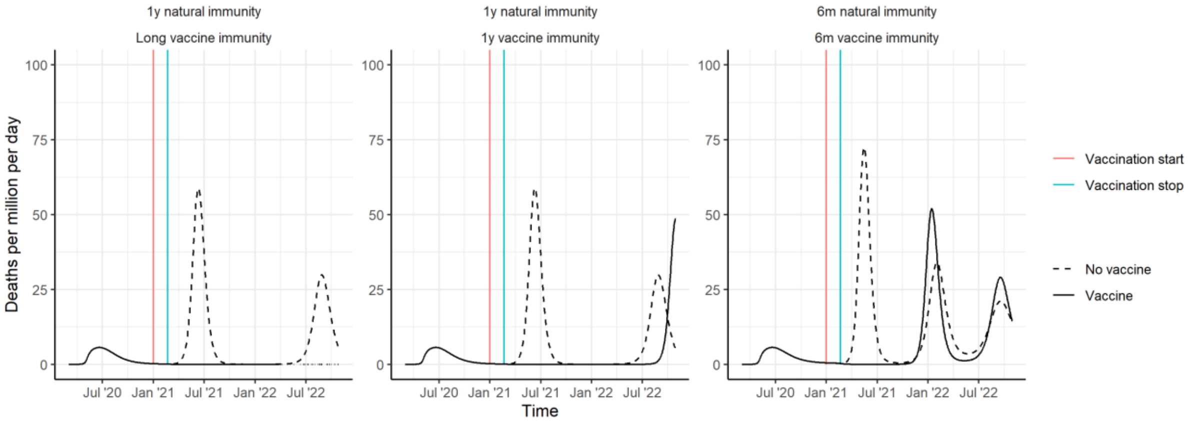


***Figure S8: Epidemic trajectories and impact of immunity.*** *Epidemic scenarios are shown for the period 2020–2022, both in the absence of a vaccine (dashed black lines) and following vaccine introduction (solid black lines). Vaccine implementation is indicated by the red and blue vertical lines. The left panel represents the scenario where vaccine-derived immunity is long-term, and naturally derived immunity is one year. The middle panel represents where both vaccine- and naturally derived immunity are one year, while the right panel shows trajectories where both durations are six months. Trajectories are shown for a high-income country setting, and assuming the default transmission and vaccine parameters as in Table 1.*

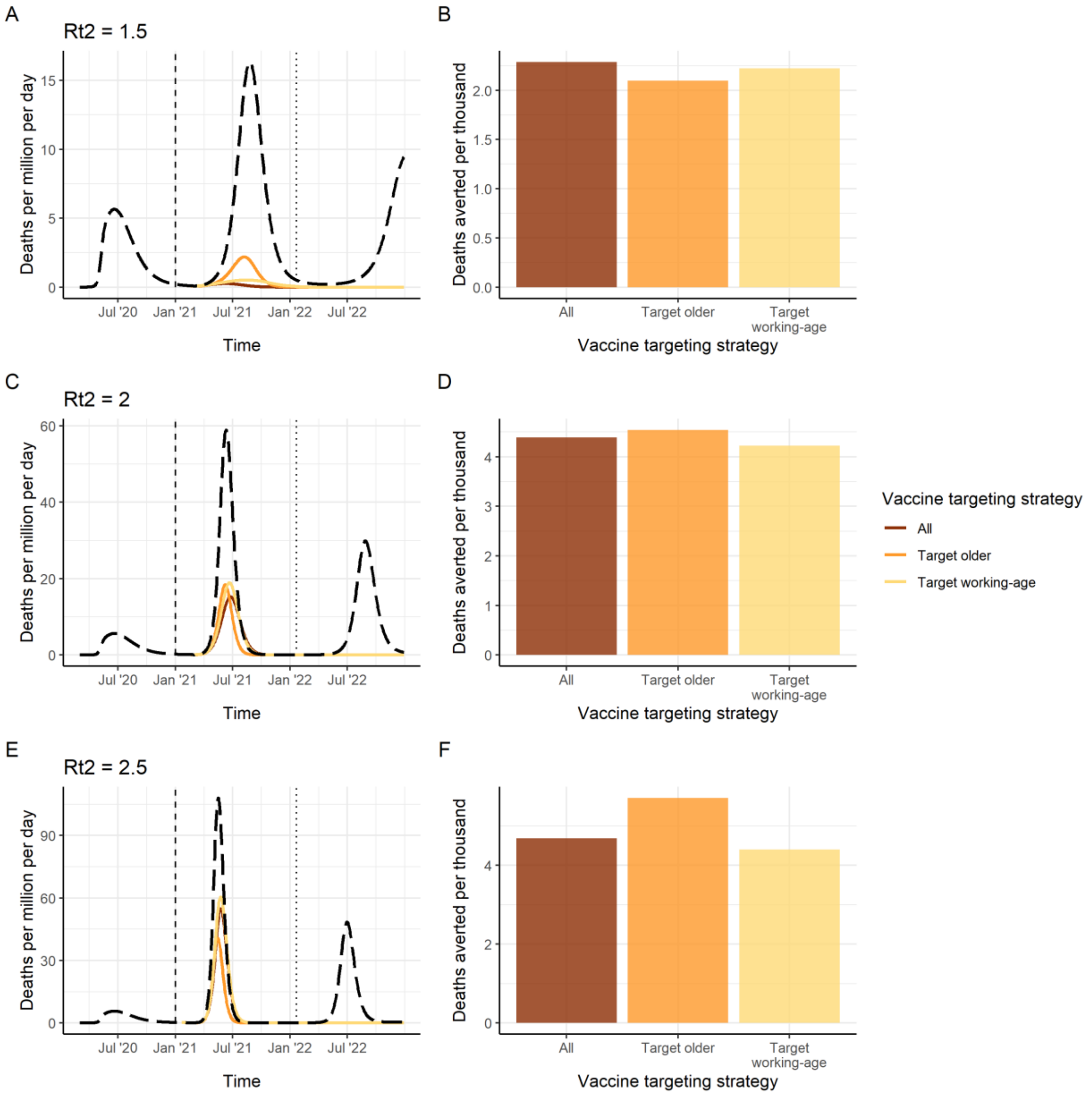


***Figure S9: Vaccine impact over a one-year vaccination period.*** *Here, NPIs are assumed to be lifted when the vaccine is introduced, rather than after the target population is vaccinated, and the target population is vaccinated over a period of one year. Vaccination start is shown by the vertical dashed line, and vaccination stop by the vertical dotted line. Three different values of $R_{t2}$ are shown: $R_{t2}$ = 1.5 (A,B); $R_{t2}$ = 2.0 (C,D; the default value) and $R_{t2}$ = 2.5 (E,F). The left-hand-side panels show the deaths per million per day, for the counterfactual scenario (long-dashed black line), and three vaccine targeting strategies (coloured lined). The right-hand-side panels show the total deaths averted in 2021–2022 per thousand population for three vaccine targeting strategies, where vaccination takes place at a constant rate over the course of 2021. "All": all age groups vaccinated simultaneously. "Target older": the 80+ group is vaccinated first, then additional groups (75–79, 70–74 and so on) are consecutively vaccinated. "Target working-age": the 15–64-year-old group is vaccinated first, and then the older group, and then children.*

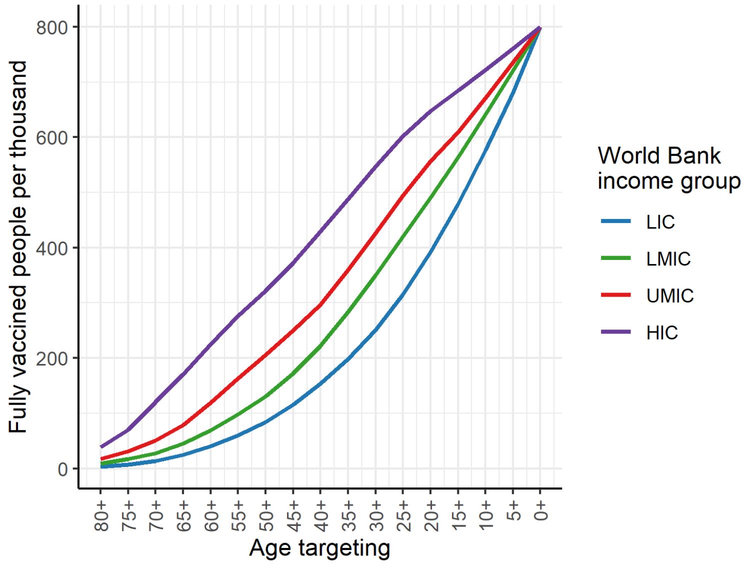


***Figure S10: Resources required when vaccine introduction is targeted by age.*** *Here, the highest risk age groups are prioritised. Resources required is presented as a proportion of the total population.*

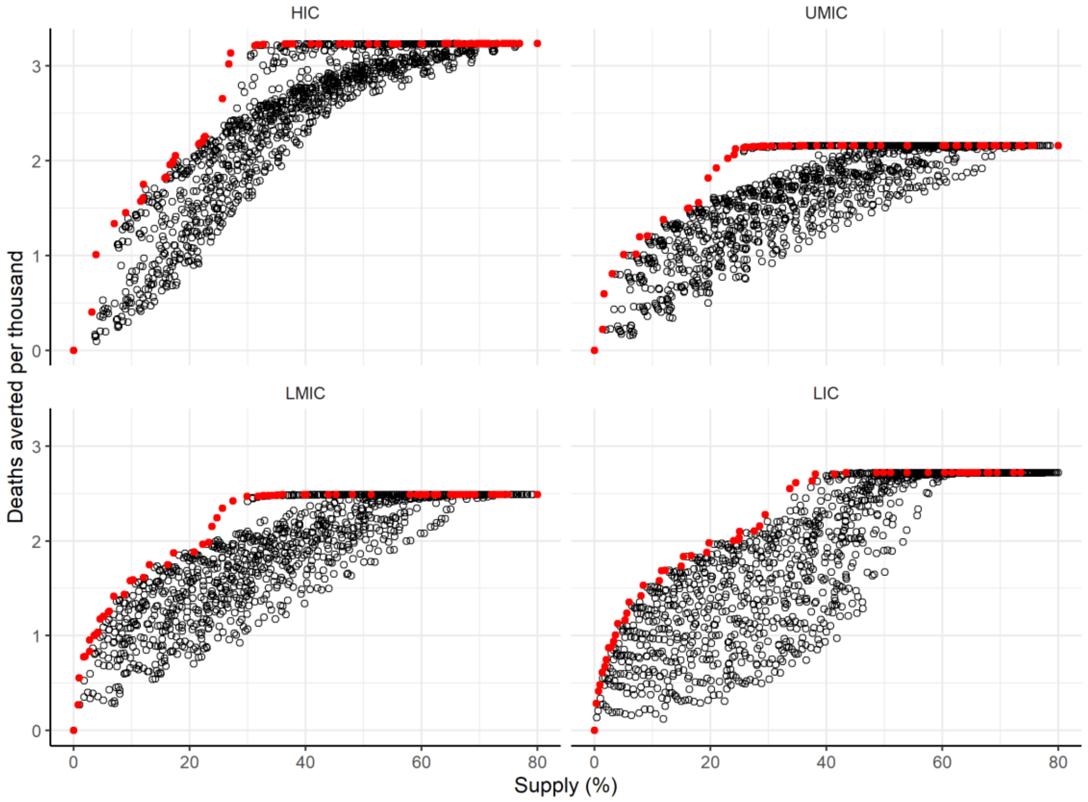


***Figure S11: Efficiency frontier for the age targeting of a vaccine within each income setting.*** *The black circles each represent a unique age targeting strategy, for each income setting, for increasing availability of doses on the x-axis, versus impact in terms of deaths averted per thousand population on the y-axis. The red points represent the most efficient (non-dominated) age-targeting strategies, or the maximum deaths averted as the vaccine supply is increased. These red points correspond to the age targeting strategies shown in Figure 4A, C, E and G, and the optimal allocation strategy in Figure 4B, D, F and H.*

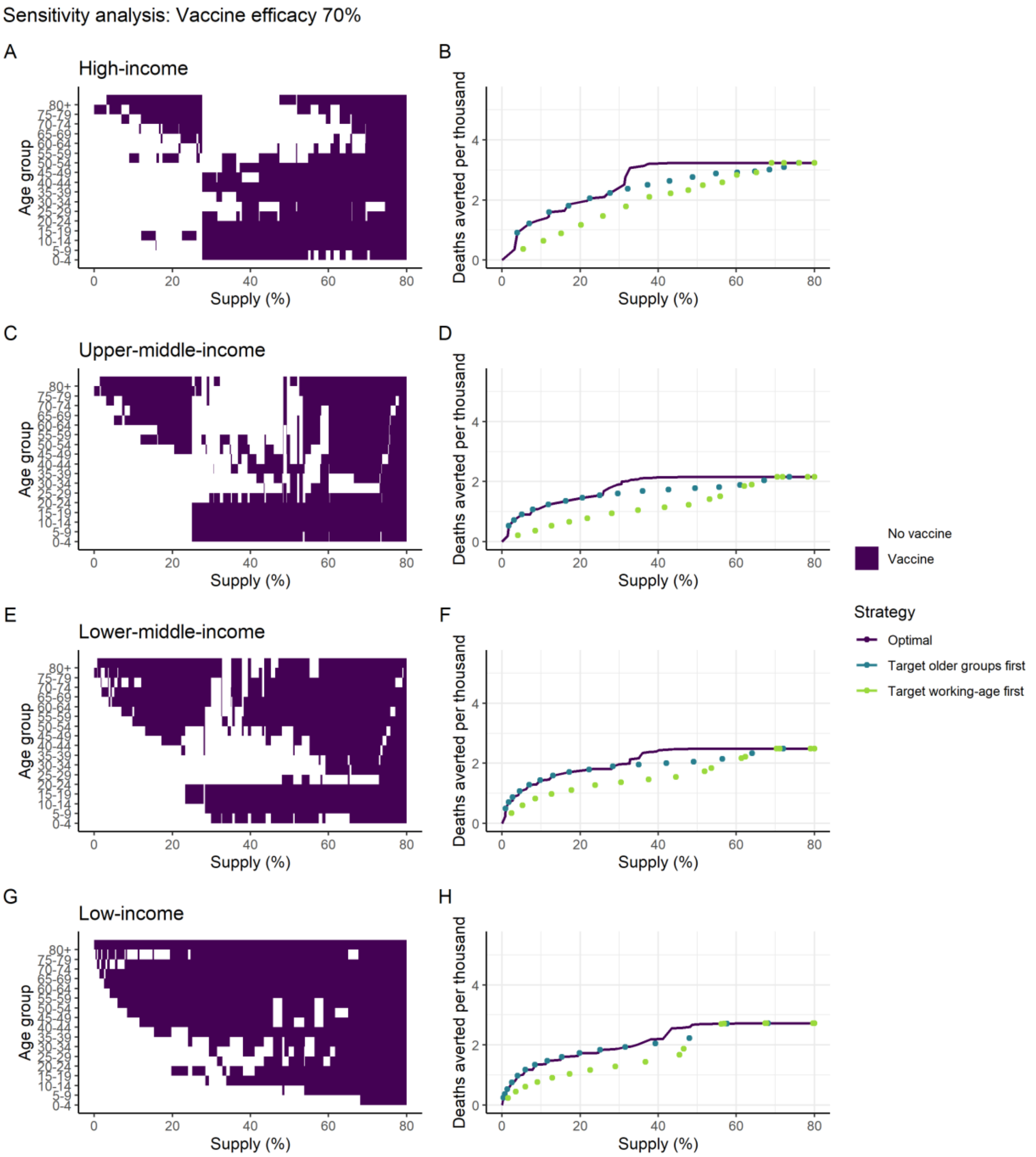


***Figure S12: Sensitivity analysis of targeting of vaccine introduction within each income setting; lower vaccine efficacy (70%).*** *These panels illustrate the most efficient allocation under different dose constraints, where the supply is defined as the proportion of the population able to access two doses. Panels A, C, E and G show the age groups allocated under each supply level, where the purple shaded regions indicate the age groups allocated the vaccine. Panels B, D, F and H show the efficiency frontiers expressed as deaths averted per thousand population as a function of vaccine supply. The optimal strategies from the left-hand panels are shown in purple. The turquoise shows the strategy that prioritises the older at-risk age: 80+ for the lowest coverage level, and sequentially including additional age groups (75–79, 70–74 and so on) as additional doses are available. The green strategy prioritises the working age population first (beginning with the 60–64 age group and sequentially adding younger groups), then vaccinates the elderly and children as doses become available.*

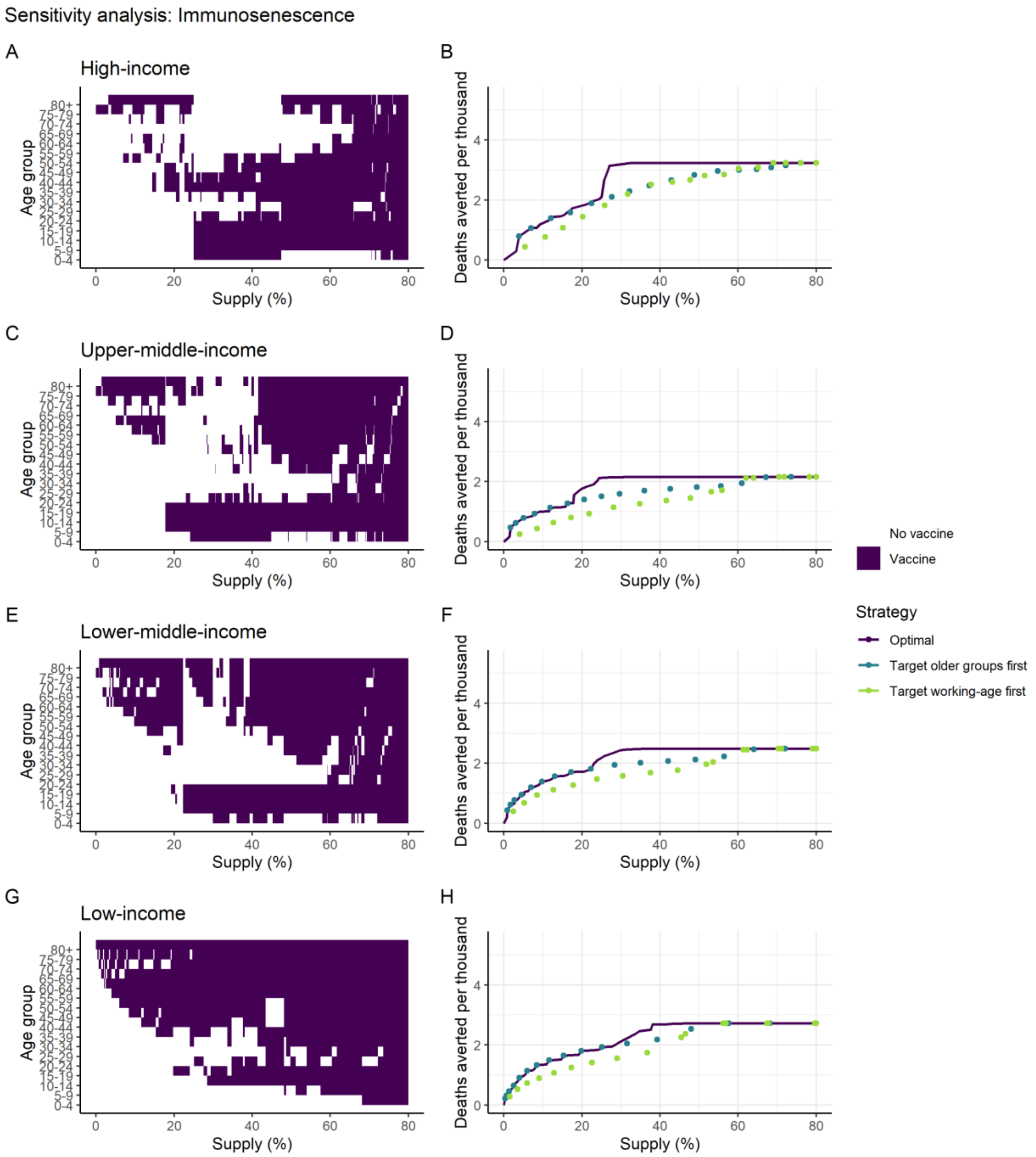


***Figure S13: Sensitivity analysis of targeting of vaccine introduction within each income setting; reduced vaccine impact in 65+ age group (immunosenescence).*** *Vaccine efficacy was reduced in the 65+ age group to 35%. These panels illustrate the most efficient allocation under different dose constraints, where the supply is defined as the proportion of the population able to access two doses. Panels A, C, E and G show the age groups allocated under each supply level, where the purple shaded regions indicate the age groups allocated the vaccine. Panels B, D, F and H show the efficiency frontiers expressed as deaths averted per thousand population as a function of vaccine supply. The optimal strategies from the left-hand panels are shown in purple. The turquoise shows the strategy that prioritises the older at-risk age: 80+ for the lowest coverage level, and sequentially including additional age groups (75–79, 70–74 and so on) as additional doses are available. The green strategy prioritises the working age population first (beginning with the 60–64 age group and sequentially adding younger groups), then vaccinates the elderly and children as doses become available.*

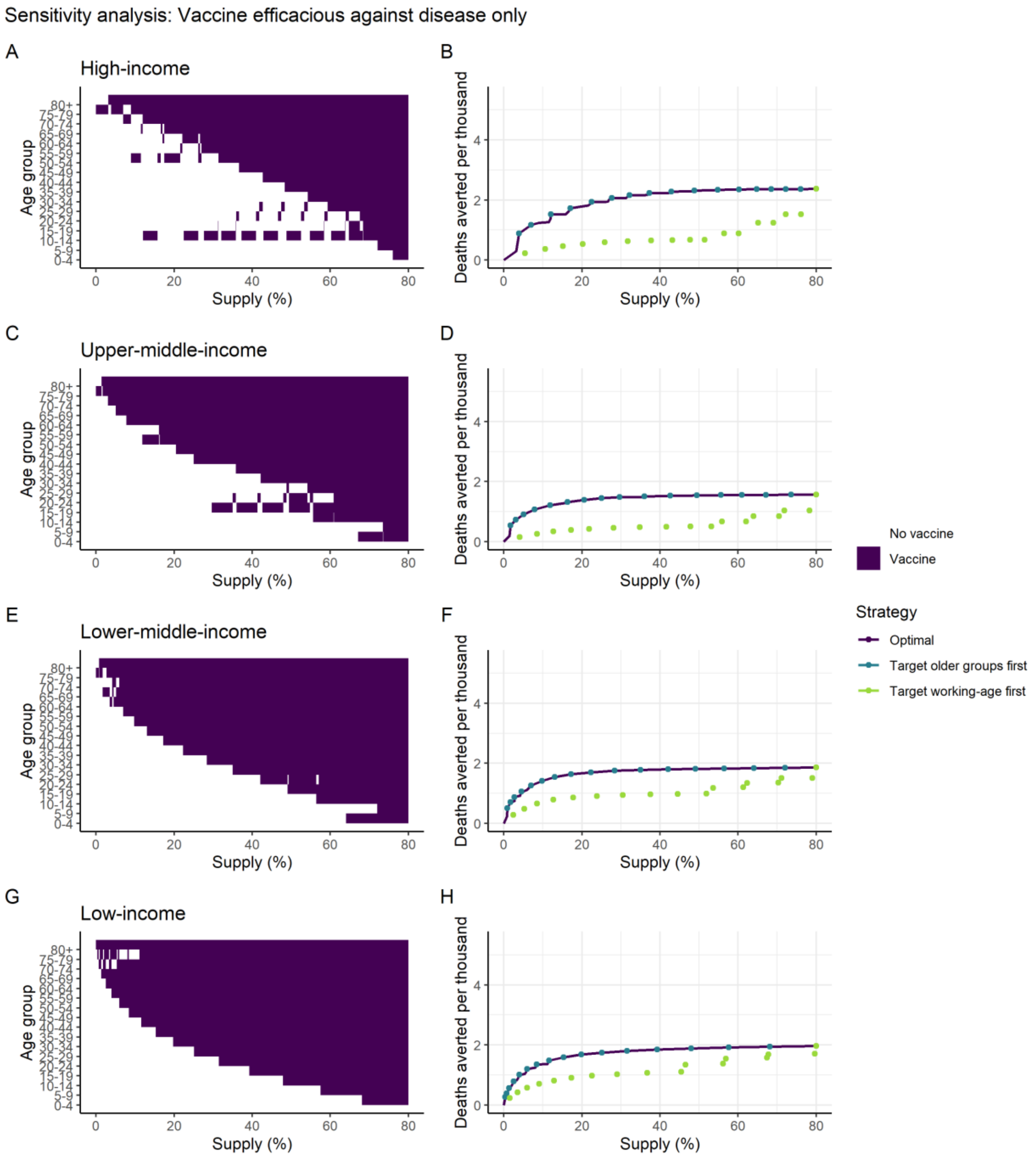


***Figure S14: Sensitivity analysis of targeting of vaccine introduction within each income setting; vaccine efficacious against disease only.*** *These panels illustrate the most efficient allocation under different dose constraints, where the supply is defined as the proportion of the population able to access two doses. Panels A, C, E and G show the age groups allocated under each supply level, where the purple shaded regions indicate the age groups allocated the vaccine. Panels B, D, F and H show the efficiency frontiers expressed as deaths averted per thousand population as a function of vaccine supply. The optimal strategies from the left-hand panels are shown in purple. The turquoise shows the strategy that prioritises the older at-risk age: 80+ for the lowest coverage level, and sequentially including additional age groups (75–79, 70–74 and so on) as additional doses are available. The green strategy prioritises the working age population first (beginning with the 60–64 age group and sequentially adding younger groups), then vaccinates the elderly and children as doses become available.*

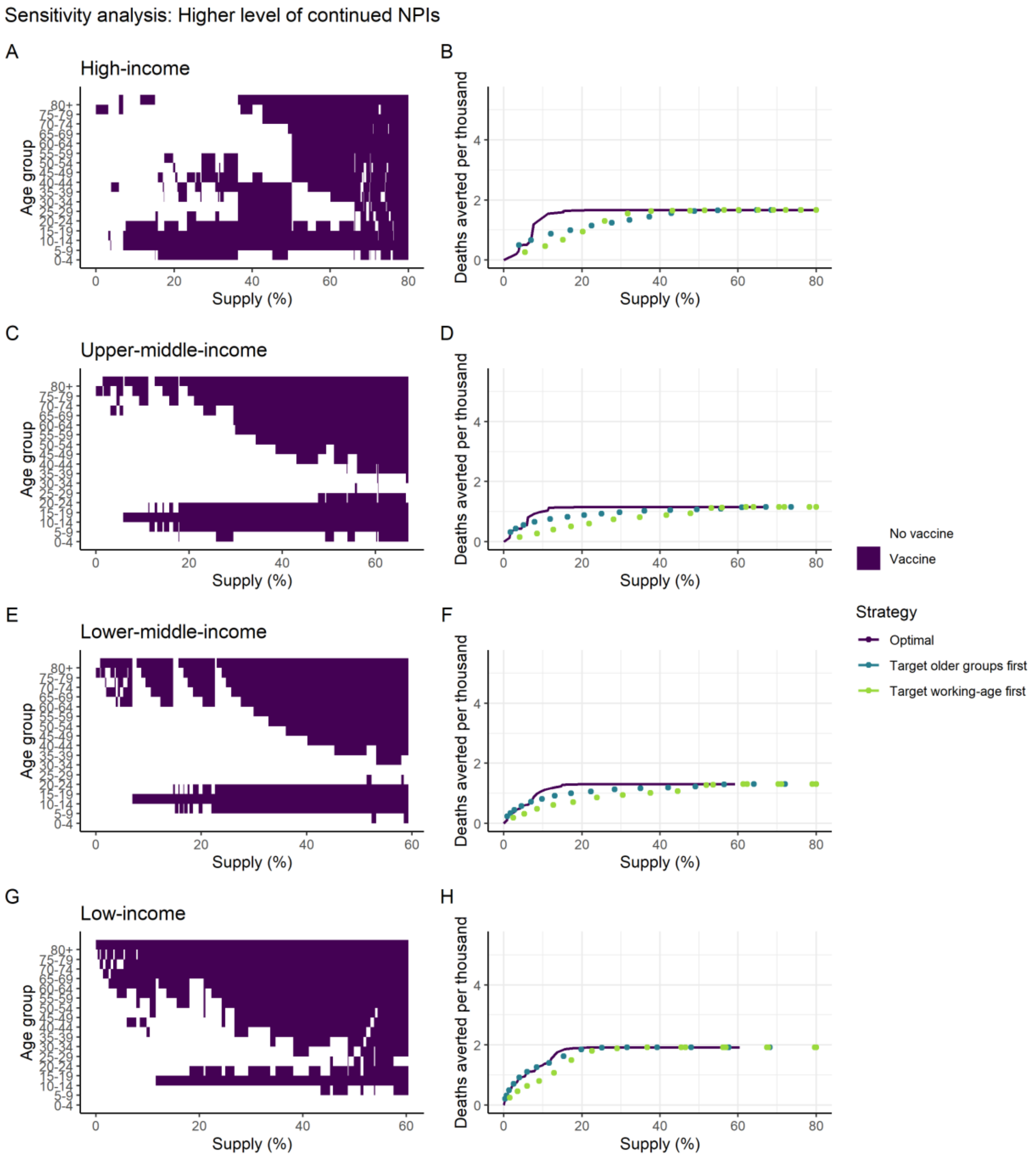


***Figure S15: Sensitivity analysis of targeting of vaccine introduction within each income setting; $R_{t2}$=1.5.*** *These panels illustrate the most efficient allocation under different dose constraints, where the supply is defined as the proportion of the population able to access two doses. Panels A, C, E and G show the age groups allocated under each supply level, where the purple shaded regions indicate the age groups allocated the vaccine. Panels B, D, F and H show the efficiency frontiers expressed as deaths averted per thousand population as a function of vaccine supply. The optimal strategies from the left-hand panels are shown in purple. The turquoise shows the strategy that prioritises the older at-risk age: 80+ for the lowest coverage level, and sequentially including additional age groups (75–79, 70–74 and so on) as additional doses are available. The green strategy prioritises the working age population first (beginning with the 60–64 age group and sequentially adding younger groups), then vaccinates the elderly and children as doses become available.*

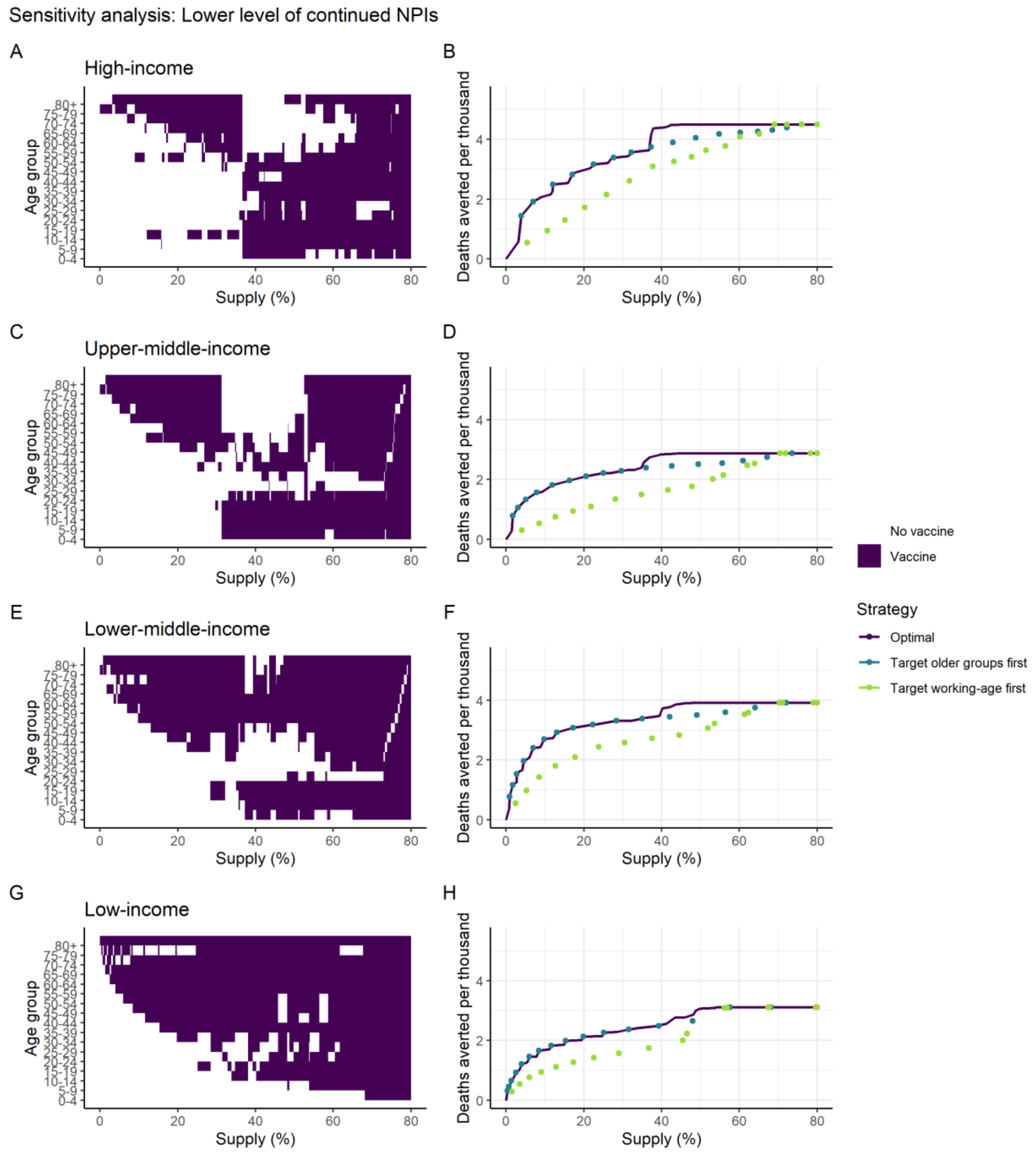


***Figure S16: Sensitivity analysis of targeting of vaccine introduction within each income setting; $R_{t2}$=2.5.*** *These panels illustrate the most efficient allocation under different dose constraints, where the supply is defined as the proportion of the population able to access two doses. Panels A, C, E and G show the age groups allocated under each supply level, where the purple shaded regions indicate the age groups allocated the vaccine. Panels B, D, F and H show the efficiency frontiers expressed as deaths averted per thousand population as a function of vaccine supply. The optimal strategies from the left-hand panels are shown in purple. The turquoise shows the strategy that prioritises the older at-risk age: 80+ for the lowest coverage level, and sequentially including additional age groups (75–79, 70–74 and so on) as additional doses are available. The green strategy prioritises the working age population first (beginning with the 60–64 age group and sequentially adding younger groups), then vaccinates the elderly and children as doses become available.*

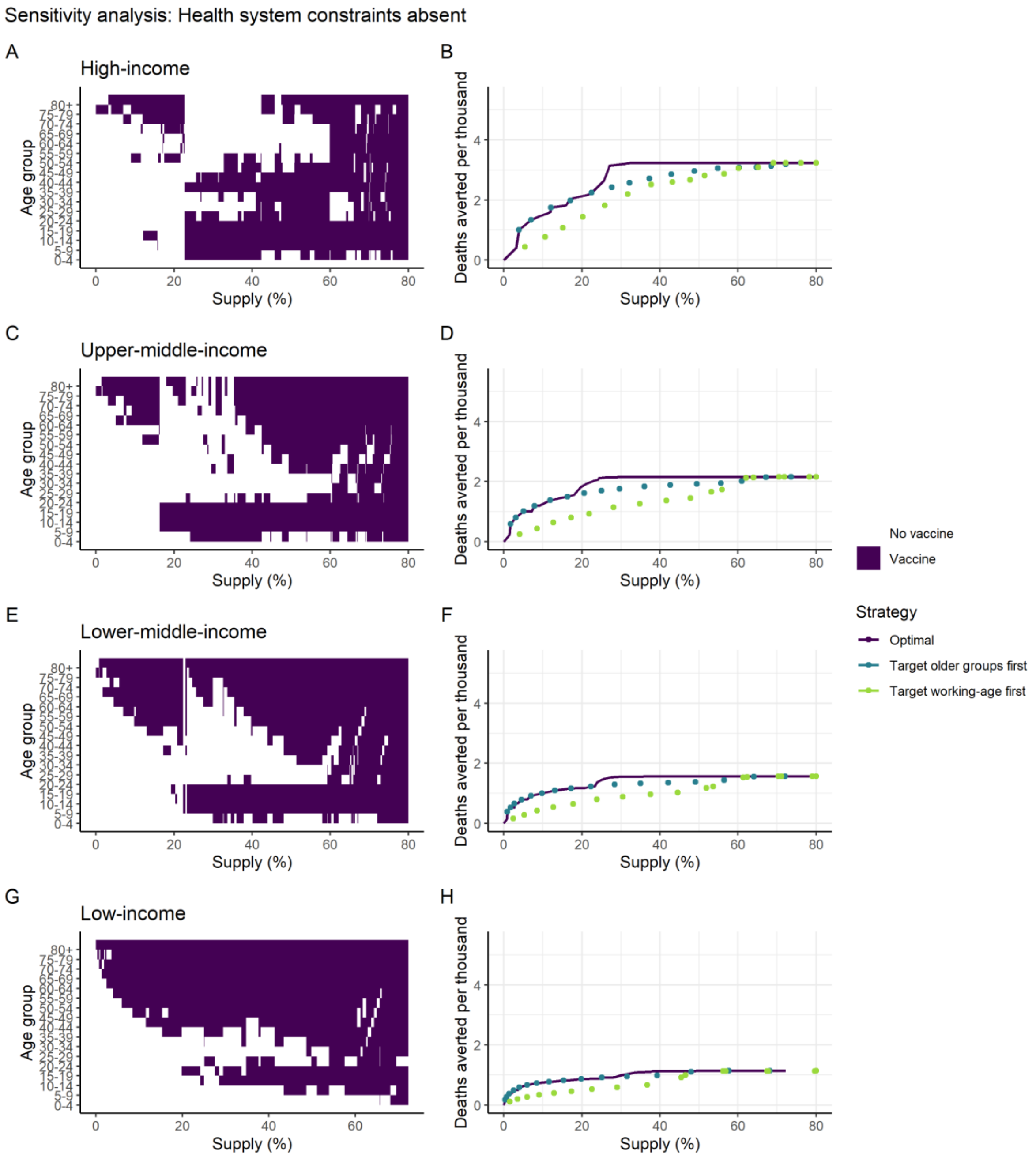


***Figure S17: Sensitivity analysis of targeting of vaccine introduction within each income setting; health system constraints absent.*** *These panels illustrate the most efficient allocation under different dose constraints, where the supply is defined as the proportion of the population able to access two doses. Panels A, C, E and G show the age groups allocated under each supply level, where the purple shaded regions indicate the age groups allocated the vaccine. Panels B, D, F and H show the efficiency frontiers expressed as deaths averted per thousand population as a function of vaccine supply. The optimal strategies from the left-hand panels are shown in purple. The turquoise shows the strategy that prioritises the older at-risk age: 80+ for the lowest coverage level, and sequentially including additional age groups (75–79, 70–74 and so on) as additional doses are available. The green strategy prioritises the working age population first (beginning with the 60–64 age group and sequentially adding younger groups), then vaccinates the elderly and children as doses become available.*

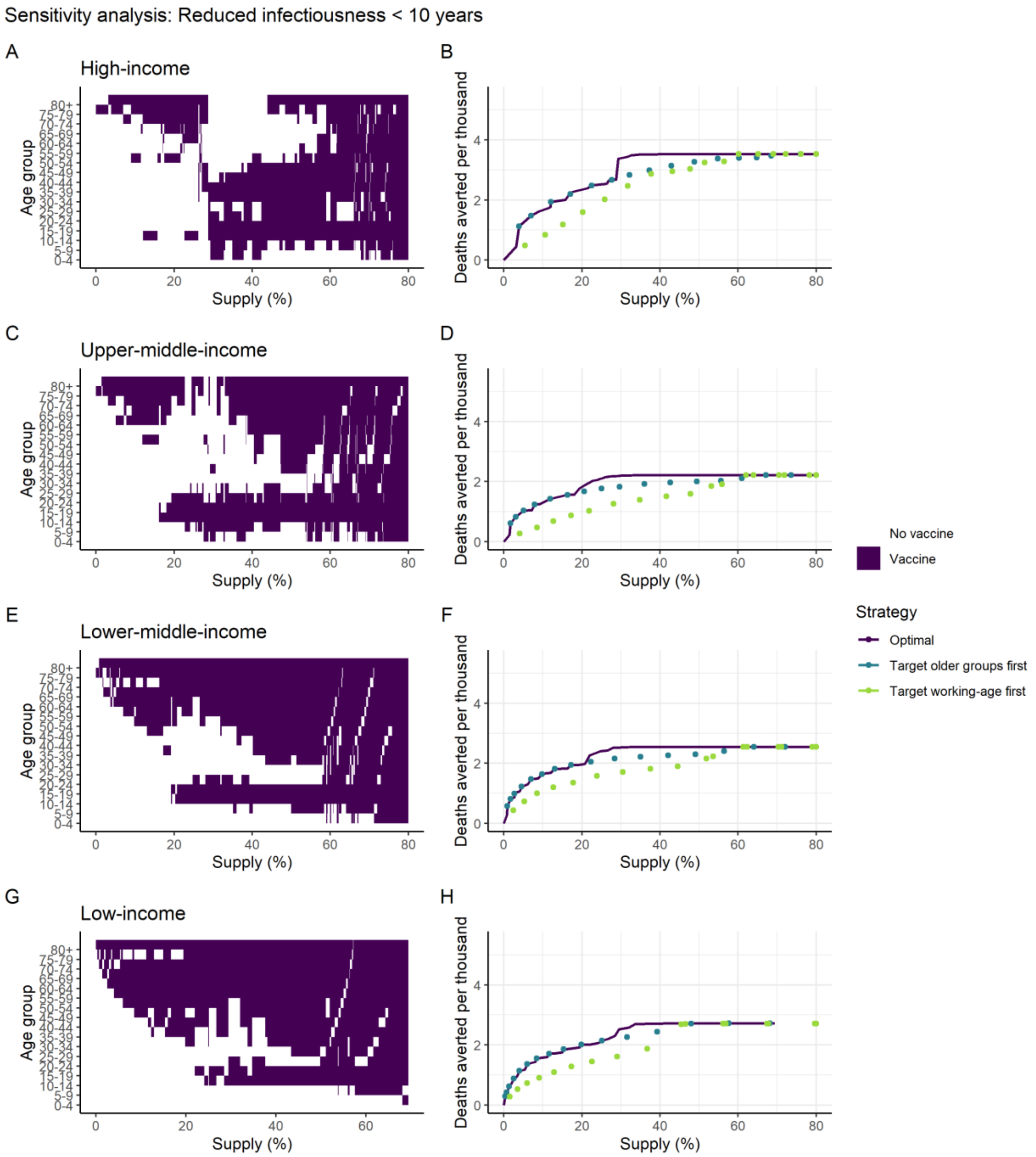


***Figure S18: Sensitivity analysis of targeting of vaccine introduction within each income setting; transmission from children younger than 10 years reduced by 50%.*** *These panels illustrate the most efficient allocation under different dose constraints, where the supply is defined as the proportion of the population able to access two doses. Panels A, C, E and G show the age groups allocated under each supply level, where the purple shaded regions indicate the age groups allocated the vaccine. Panels B, D, F and H show the efficiency frontiers expressed as deaths averted per thousand population as a function of vaccine supply. The optimal strategies from the left-hand panels are shown in purple. The turquoise shows the strategy that prioritises the older at-risk age: 80+ for the lowest coverage level, and sequentially including additional age groups (75–79, 70–74 and so on) as additional doses are available. The green strategy prioritises the working age population first (beginning with the 60–64 age group and sequentially adding younger groups), then vaccinates the elderly and children as doses become available.*

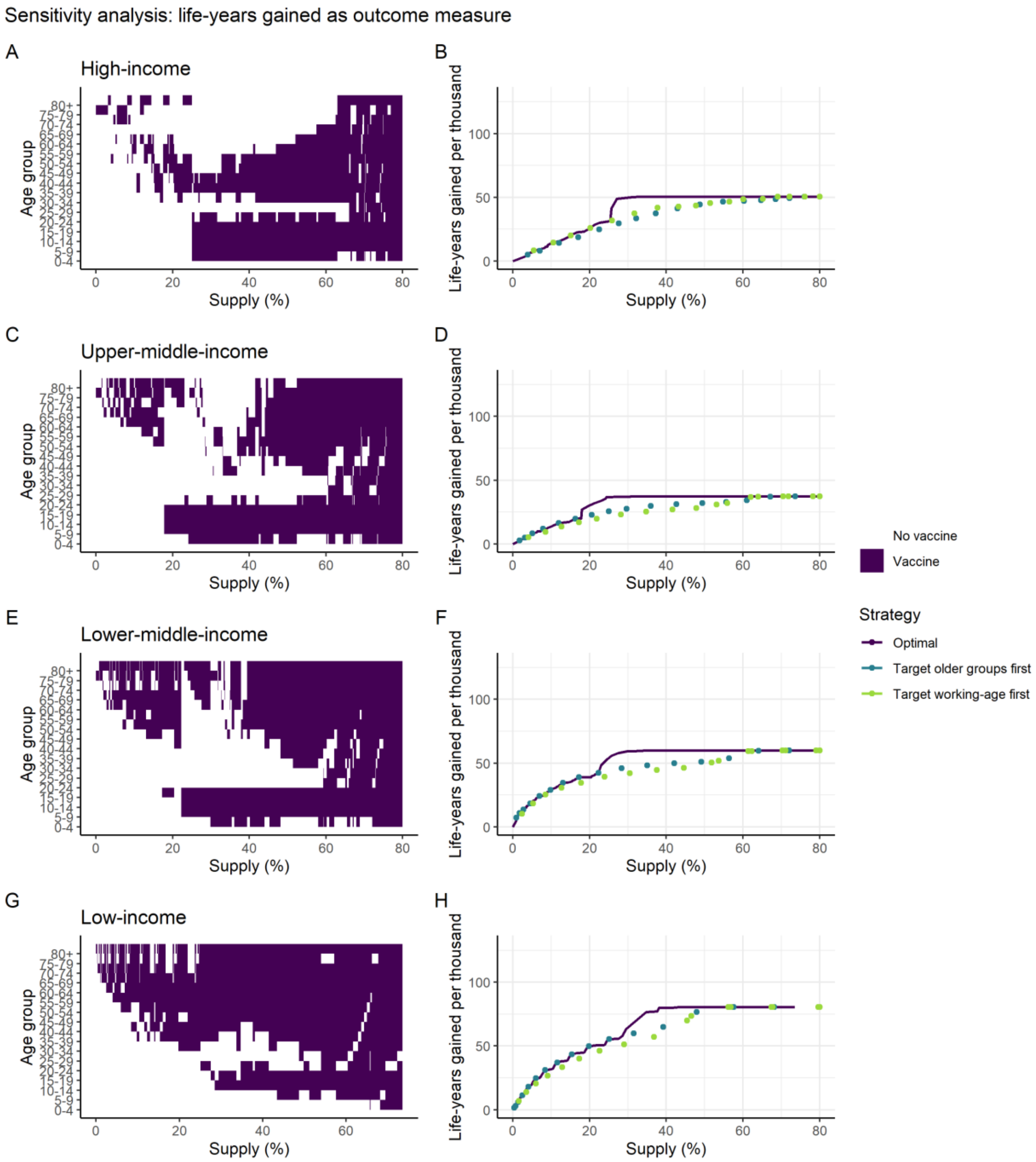


***Figure S19: Sensitivity analysis of targeting of vaccine introduction within each income setting; life-years gained as outcome measure.*** *These panels illustrate the most efficient allocation under different dose constraints, where the supply is defined as the proportion of the population able to access two doses. Panels A, C, E and G show the age groups allocated under each supply level, where the purple shaded regions indicate the age groups allocated the vaccine. Panels B, D, F and H show the efficiency frontiers expressed as life-years gained per thousand population as a function of vaccine supply. The optimal strategies from the left-hand panels are shown in purple. The turquoise shows the strategy that prioritises the older at-risk age: 80+ for the lowest coverage level, and sequentially including additional age groups (75–79, 70–74 and so on) as additional doses are available. The green strategy prioritises the working age population first (beginning with the 60–64 age group and sequentially adding younger groups), then vaccinates the elderly and children as doses become available.*

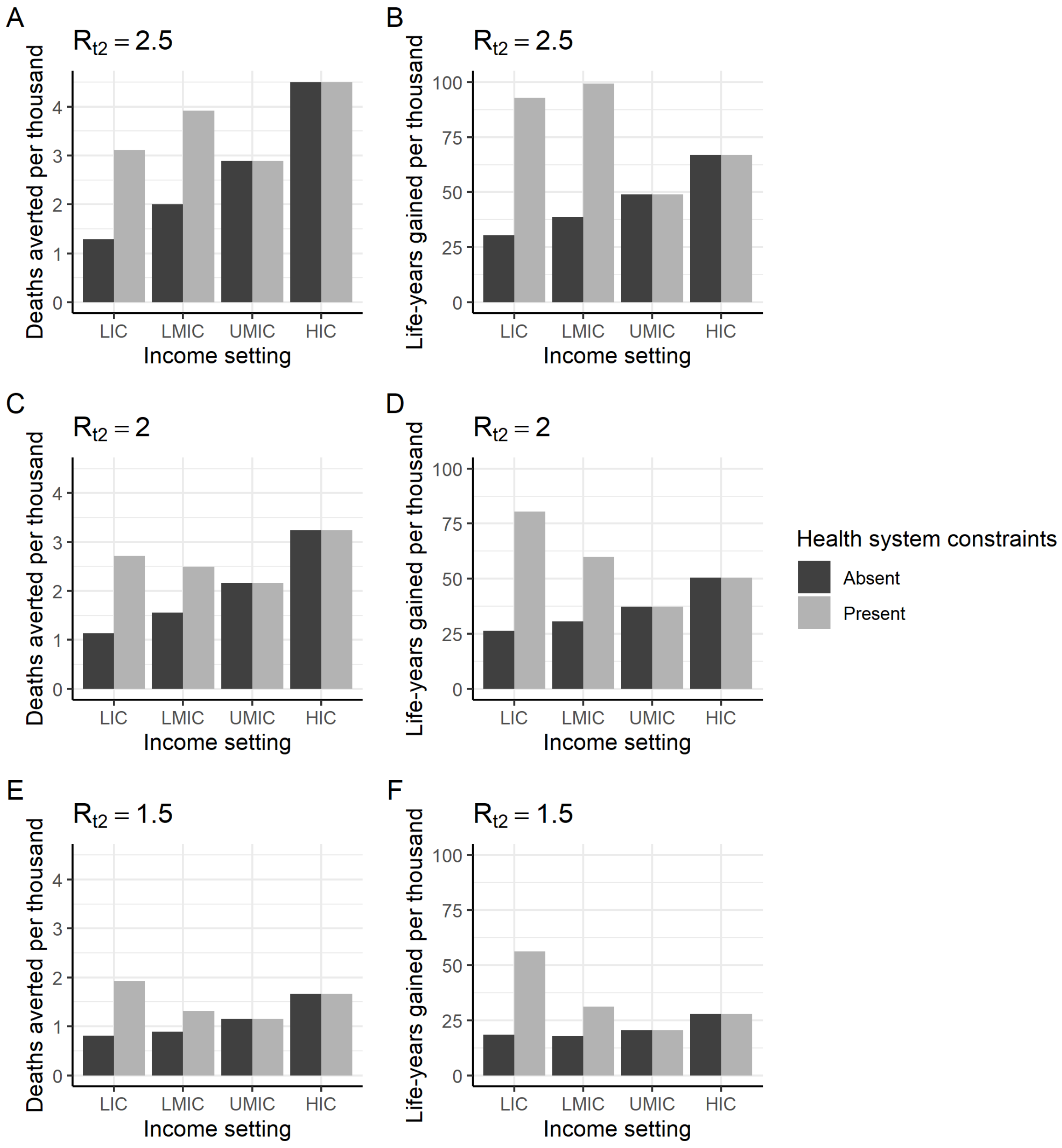


***Figure S20: Vaccine impact by income setting and level of NPIs at vaccine introduction.*** *Deaths averted (A, C, E) and life years gained (B, D, F) per thousand population in 2021 for each income setting (x-axis), where health systems are either unconstrained (dark grey) or constrained (light grey), and for* $R_{t2}$ *= 2.5 (upper row),* $R_{t2}$*=2 (default value, middle row) and* $R_{t2}$*=1.5 (lower row). Default vaccine parameters are in Table 1.*

***Table S4: Global allocation of vaccine doses for non-optimised scenarios***. *Here we assume that limited countries within each income setting are allocated doses at high (80%) coverage, rather than all countries being allocated doses at a lower level of coverage as in Table 2. The global vaccine supply is assumed to be constrained to 2 billion doses, with a two-dose schedule and 15% buffer and wastage (resulting in 0.85 billion vaccine courses available). FVP: fully vaccinated persons.*

| **Allocation strategy** | | **Income setting** | **Target age group** | **Deaths averted per million** | **Deaths averted per 100 FVP** | **Total deaths averted per million global population** | **Total deaths averted per 100 FVP** |
|---|---|---|---|---|---|---|---|
| Allocated to limited countries at 80% coverage | A: Countries receive doses in proportion to population | HIC | all | 450 | 0.4 | 348 | 0.31 |
| | | UMIC | all | 300 | 0.27 | | |
| | | LMIC | all | 346 | 0.31 | | |
| | | LIC | all | 378 | 0.34 | | |
| | B: Countries receive doses in proportion to population, with 65+ group prioritised and remaining doses allocated to 15-64 age groups | HIC | 15+ | 1293 | 1.16 | 1316 | 1.18 |
| | | UMIC | 15+ | 1268 | 1.14 | | |
| | | LMIC | 15+ | 1388 | 1.25 | | |
| | | LIC | 15+ | 1238 | 1.11 | | |
| | C: Countries receive doses in proportion to 65+ population, with 65+ group prioritised and remaining doses allocated to 15-64 age groups | HIC | 15+ | 2260 | 0.9 | 1394 | 1.25 |
| | | UMIC | 15+ | 1252 | 1.09 | | |
| | | LMIC | 15+ | 1279 | 1.91 | | |
| | | LIC | 15+ | 932 | 2.56 | | |
| | D: Allocated first to high-income countries | HIC | all | 2837 | 0.4 | 450 | 0.40 |
| | | UMIC | all | 0 | 0 | | |
| | | LMIC | all | 0 | 0 | | |
| | | LIC | all | 0 | 0 | | |
| | E: Allocated first to low-income and lower-middle-income countries | HIC | all | 0 | 0 | 352 | 0.32 |
| | | UMIC | all | 0 | 0 | | |
| | | LMIC | all | 740 | 0.31 | | |
| | | LIC | all | 808 | 0.34 | | |
| | F: Countries receive doses in proportion to population, plus 1.15 b doses to HIC and 1.1 b doses to MIC | HIC | all | 2081 | 0.4 | 785 | 0.33 |
| | | UMIC | all | 519 | 0.27 | | |
| | | LMIC | all | 598 | 0.31 | | |
| | | LIC | all | 378 | 0.34 | | |

***Table S5: Optimised global allocation of vaccine doses for different assumptions about vaccine characteristics, transmission, health system constraints, and optimisation outcome.***

| **Parameter assumption** | **Income setting** | **Deaths averted per million** | **Deaths averted per 100 fully vaccinated persons** | **Total deaths averted per million global population** | **Total deaths averted per 100 fully vaccinated persons** |
|---|---|---|---|---|---|
| Default | HIC | 3135 | 1.16 | 1672 | 1.43 |
| | UMIC | 1241 | 1.44 | | |
| | LMIC | 1581 | 1.62 | | |
| | LIC | 1533 | 1.81 | | |
| Lower vaccine efficacy (70%) | HIC | 3068 | 0.94 | 1497 | 1.28 |
| | UMIC | 1073 | 1.37 | | |
| | LMIC | 1365 | 1.61 | | |
| | LIC | 1335 | 1.58 | | |
| Reduced vaccine efficacy (scaled by 50%) in 65+ years population | HIC | 3135 | 1.16 | 1503 | 1.28 |
| | UMIC | 1235 | 1.07 | | |
| | LMIC | 1202 | 1.73 | | |
| | LIC | 1330 | 1.57 | | |
| Vaccine efficacious against disease only | HIC | 1773 | 0.97 | 1391 | 1.19 |
| | UMIC | 1068 | 1.36 | | |
| | LMIC | 1544 | 1.18 | | |
| | LIC | 1483 | 1.28 | | |
| NPIs maintained at higher level following vaccine introduction (such that $R_{t2}$=1.5) | HIC | 1551 | 1.37 | 1259 | 1.07 |
| | UMIC | 1131 | 0.99 | | |
| | LMIC | 1145 | 1.02 | | |
| | LIC | 1856 | 1.19 | | |
| NPIs maintained at lower level following vaccine introduction (such that $R_{t2}$=2.5) | HIC | 3734 | 1.28 | 2331 | 1.99 |
| | UMIC | 1573 | 2.01 | | |
| | LMIC | 2698 | 2.76 | | |
| | LIC | 1659 | 1.96 | | |
| Health system constraints absent | HIC | 3135 | 1.16 | 1417 | 1.21 |
| | UMIC | 1602 | 1.06 | | |
| | LMIC | 790 | 1.74 | | |
| | LIC | 585 | 1.46 | | |
| Reduced infectiousness in children younger than 10 years | HIC | 3381 | 1.16 | 1730 | 1.48 |
| | UMIC | 1238 | 1.58 | | |
| | LMIC | 1639 | 1.68 | | |
| | LIC | 1549 | 1.83 | | |
| Life-years gained as optimisation outcome measure | HIC | 3135 | 1.16 | 1532 | 1.31 |
| | UMIC | 593 | 3.52 | | |
| | LMIC | 1612 | 1.51 | | |
| | LIC | 2614 | 0.75 | | |

*Table S6: Sensitivity analysis for the fixed global vaccine allocation scenarios.*

| | | **Sensitivity Analysis**<br>Total deaths averted per million global population<br>(Total deaths averted per 100 fully vaccinated people) | | | | | | |
|---|---|---|---|---|---|---|---|---|
| | Default | Lower vaccine efficacy (70%) | Reduced vaccine efficacy (scaled by 50%) in 65+ years population | Vaccine efficacious against disease only | NPIs maintained at higher level following vaccine introduction (such that $R_{t2}$=1.5) | NPIs maintained at lower level following vaccine introduction (such that $R_{t2}$=2.5) | Health system constraints absent | Reduced infectiousness in children younger than 10 years |
| A: Countries receive doses in proportion to population | 650 (0.59) | 524 (0.48) | 609 (0.55) | 289 (0.26) | 562 (0.51) | 809 (0.74) | 484 (0.44) | 671 (0.61) |
| B: Countries receive doses in proportion to population, targeted first to 65+, then 15-64 age groups | 1324 (1.2) | 1182 (1.07) | 1082 (0.98) | 1118 (1.01) | 756 (0.68) | 1936 (1.75) | 1091 (0.99) | 1393 (1.26) |
| C: Countries receive doses in proportion to population in 65+ age group, targeted first to 65+, then 15-64 age groups | 1386 (1.25) | 1245 (1.12) | 1135 (1.02) | 1188 (1.07) | 767 (0.69) | 1995 (1.8) | 1193 (1.07) | 1461 (1.32) |
| D: Allocated first to high-income countries | 513 (0.46) | 513 (0.46) | 513 (0.46) | 330 (0.3) | 264 (0.24) | 713 (0.64) | 513 (0.46) | 559 (0.5) |
| E: Allocated first to low-income and lower-middle-income countries | 597 (0.53) | 484 (0.43) | 571 (0.51) | 319 (0.28) | 635 (0.57) | 908 (0.81) | 331 (0.29) | 610 (0.54) |
| F: Countries receive doses in proportion to population, plus additional 1.15 b doses to HIC and 1.1 b doses to MIC | 1308 (0.56) | 1103 (0.47) | 1266 (0.54) | 625 (0.27) | 1075 (0.46) | 1710 (0.73) | 1098 (0.47) | 1374 (0.59) |

36 Hall V, Foulkes S, Saei A, *et al.* Effectiveness of BNT162b2 mRNA Vaccine Against Infection and COVID-

19 Vaccine Coverage in Healthcare Workers in England, Multicentre Prospective Cohort Study (the SIREN Study). *SSRN Prepr* 2021. https://papers.ssrn.com/sol3/papers.cfm?abstract_id=3790399.

37 MRC Centre for Global Infectious Disease Analysis Imperial College London. Future scenarios of the healthcare burden of COVID-19 in Low- or Middle-income countries. 2021. https://mrc-ide.github.io/global-lmic-reports/ (accessed Jan 26, 2021).

38 Dong E, Du H, Gardner L. An interactive web-based dashboard to track COVID-19 in real time. *Lancet Infect Dis* 2020; **20**: 533–4.

39 Worldometer. Coronavirus updates. https://www.worldometers.info/ (accessed Jan 26, 2021).
